# Conversational Artificial Intelligence Agents-Enabled Dissection of RTK-RAS and MAPK Pathway Dependencies in Gemcitabine-Treated Pancreatic Ductal Adenocarcinoma (PDAC)

**DOI:** 10.64898/2026.03.01.26347364

**Authors:** Fernando C. Diaz, Brigette Waldrup, Francisco G. Carranza, Sophia Manjarrez, Enrique Velazquez-Villarreal

**Author notes:** These authors contributed equally to this work (Fernando C. Diaz, Enrique Velazquez-Villarreal).

## Abstract

Pancreatic ductal adenocarcinoma (PDAC) is an aggressive malignancy characterized by profound molecular heterogeneity and inconsistent responses to gemcitabine-based therapy. Although KRAS mutations are nearly ubiquitous, the broader RTK-RAS and MAPK signaling networks, and their association with therapeutic response, remain insufficiently characterized.

We performed an integrative clinical-genomic study of 184 PDAC tumors, stratified by age at diagnosis and gemcitabine exposure, systematically evaluating somatic alterations within curated RTK–RAS/MAPK gene panels. Conversational artificial intelligence agents (AI-HOPE-RTK-RAS and AI-HOPE-MAPK) were deployed to dynamically construct cohorts and conduct pathway-level analyses, with results subsequently confirmed using conventional statistical approaches.

Among late-onset PDAC cases, ERBB2 and RET mutations were significantly enriched in gemcitabine-treated tumors. In early-onset disease, CACNA2D family alterations were more common in untreated tumors, whereas FLNB and TP53 mutations were observed at higher frequencies in treated cases. Notably, late-onset patients who did not receive gemcitabine and lacked RTK-RAS or MAPK pathway alterations demonstrated significantly improved overall survival.

These findings identify age- and treatment-specific signaling dependencies extending beyond canonical KRAS alterations and reinforce a precision oncology framework in PDAC. Conversational AI enabled rapid, multidimensional integration of clinical and genomic data, facilitating the identification of clinically meaningful pathway architectures.

## 1. Introduction

Pancreatic ductal adenocarcinoma (PDAC) remains among the most lethal solid tumors, driven by late presentation, aggressive biology, and limited durability of systemic therapy responses (1,2). Despite incremental advances in multi-agent regimens, gemcitabine continues to serve as a cornerstone across metastatic, locally advanced, and perioperative settings, particularly for patients who are older, less fit, or have comorbidity profiles that limit intensive chemotherapy options (3,4). However, clinical benefit from gemcitabine is highly variable, and both intrinsic and rapidly acquired resistance emerge as dominant barriers to long-term disease control (3,5–8). This resistance is not purely tumor-cell autonomous; it is shaped by drug transport and metabolism, apoptosis evasion programs, stromal architecture, hypoxia, acidity, and immune contexture, features that collectively define the PDAC therapeutic bottleneck (5–8).

At the center of PDAC pathogenesis is aberrant RTK-RAS-MAPK signaling. Oncogenic KRAS mutations occur in the vast majority of PDACs, establishing a state of sustained pathway activation that promotes proliferation, metabolic adaptation, invasion, and therapy resistance (9–11).Yet KRAS is not a monolith: specific codon 12 variants exhibit distinct molecular features, immune phenotypes, and clinically relevant outcome differences, including differential survival across KRAS subtypes and treatment contexts (12–14). Moreover, KRAS allele state and transcriptional subtype can interact with prognosis and treatment response, underscoring that “KRAS-mutant PDAC” encompasses biologically distinct entities (15,16). This complexity matters for precision oncology because KRAS itself has historically been considered difficult to target, and even emerging mutation-specific inhibitors face rapid adaptive resistance through pathway reactivation, bypass signaling, and metabolic rewiring (11,17–19).

Gemcitabine response is also intertwined with RTK-RAS/MAPK biology and tumor-microenvironment feedback. Preclinical studies show that disrupting KRAS signaling can sensitize PDAC to gemcitabine, motivating combination strategies that simultaneously impair oncogenic signaling and cytotoxic response escape (20–22). More recent delivery platforms, including targeted lipid nanoparticles and microenvironment-aware drug release systems, have demonstrated that suppressing KRAS or downstream ERK activity can enhance gemcitabine efficacy in vivo, reinforcing the mechanistic coupling of MAPK signaling and chemotherapy response (23, 24–26). Conversely, gemcitabine itself can provoke compensatory programs that sustain aggressiveness, for example, epigenetic regulation of invasion-associated factors in gemcitabine-resistant tumor-initiating cells and metabolic reprogramming linked to KRAS-dependent signaling that promotes stem-like properties (27,28). Gemcitabine may also modulate antitumor immunity in context-dependent ways, including effects on CD8+ T-cell infiltration and potential antagonism of PD-1-based immunotherapy in KRAS-driven settings, highlighting the relevance of precision immuno-oncology framing even for a “chemotherapy backbone” drug (29,30).

These observations converge on a practical need: clinically actionable biomarkers that capture pathway dependencies beyond binary KRAS mutation status. While KRAS variants, transcriptional subtypes, and multi-omic subgrouping can stratify outcomes and reveal therapeutic vulnerabilities, translating such signals into workflow-ready precision oncology requires integrated clinical-genomic analyses that remain reproducible and scalable (12,14–16,31). Large clinicogenomic datasets now enable interrogation of how RTK-RAS and MAPK alterations distribute across clinically relevant strata (e.g., age of onset, treatment exposure), and how these alterations associate with survival outcomes under real-world care patterns (12,14,16,26,31,32,33,34). Yet traditional analytic pipelines, though rigorous, can be slow to iterate when investigators must repeatedly refine multi-parameter cohorts, test pathway-level hypotheses, and validate signals across stratifications that mirror real clinical decision-making.

To address this analytic bottleneck, we apply a conversational artificial intelligence framework AI-HOPE-RTK-RAS (35) and AI-HOPE-MAPK (36) to accelerate cohort construction and hypothesis-driven pathway analytics in gemcitabine-treated PDAC. Building on the biological premise that RTK-RAS and MAPK signaling operates as a dynamic network shaped by therapy pressure, stromal and immune context, and molecular subtype, we systematically characterize somatic alterations across these axes in a clinically annotated PDAC cohort stratified by age at diagnosis and gemcitabine exposure. We further evaluate the prognostic relevance of pathway disruption using survival analyses to identify patient subsets in which RTK-RAS/MAPK dependency may inform precision cancer medicine and future combination strategies, particularly those integrating targeted pathway inhibition with chemotherapy and, where appropriate, immunomodulatory approaches (3,4,6,7,8,17,29,37,38).

## 2. Materials and Methods

### 2.1 Study design and data provenance

We conducted a retrospective, clinicogenomic analysis of PDAC tumors with harmonized clinical, demographic, molecular, and treatment metadata. The analytic cohort comprised 184 unique PDAC cases with tumor-based next-generation sequencing (NGS) results and annotation to support age-at-diagnosis stratification and gemcitabine exposure classification. To avoid duplicate inclusion, when multiple tumor specimens or sequencing records were available for a single patient, a single representative sample was selected using a consistent rule-based approach prioritizing the most complete molecular profile and the sequencing record most proximal to the relevant treatment interval.

### 2.2 Clinical variables and subgroup definitions

Age at diagnosis was used to define early-onset PDAC (EOPDAC) and late-onset PDAC (LOPDAC) subgroups using a prespecified age threshold (applied uniformly across the cohort). Treatment exposure was abstracted from structured therapy fields and curated treatment descriptors. Patients were categorized as gemcitabine-treated if gemcitabine exposure was documented in the available treatment records; those without documented exposure were categorized as non-gemcitabine-treated. When treatment timing relative to sequencing was available, records were reviewed to ensure that gemcitabine exposure classification was clinically plausible with respect to the profiled tumor specimen.

### 2.3 RTK-RAS and MAPK pathway gene set curation and alteration definitions

Two pathway-focused gene panels were curated to represent (i) RTK-RAS signaling and (ii) MAPK signaling. The RTK-RAS panel included upstream receptor tyrosine kinases and immediate transducers (e.g., ERBB family members and RET), core RAS nodes, and key regulatory components that modulate pathway activation states. The MAPK panel encompassed canonical RAF-MEK-ERK components, pathway effectors, and regulatory feedback nodes, and additionally included selected genes with reported roles in signaling adaptation and therapy response. Somatic alterations were extracted from NGS variant calls and filtered to retain protein-altering events (missense, nonsense, frameshift indels, splice-site, and start-loss variants). Patient-level pathway alteration status was defined as the presence of ≥1 qualifying somatic alteration in any gene within the corresponding pathway panel. For gene-level analyses, alteration frequencies were computed within each clinically defined subgroup.

### 2.4 Primary endpoints and statistical analyses

The primary molecular endpoint was differential frequency of RTK-RAS and MAPK pathway alterations across age- and treatment-stratified subgroups (early-onset vs. late-onset; gemcitabine-treated vs. non-treated). Group-wise comparisons of alteration prevalence were evaluated using Fisher’s exact test (or chi-square testing when appropriate), with two-sided p-values reported and p<0.05 considered statistically significant; borderline signals were noted when p values approached the significance threshold. Overall survival (OS) was evaluated as the time from diagnosis (or from the earliest available survival anchor when diagnosis date was not available) to death or last follow-up. Survival distributions were estimated using Kaplan-Meier methods and compared using the log-rank test. Survival analyses were performed both for pathway-level alteration status and for clinically relevant subgroup interactions (e.g., late-onset PDAC non-gemcitabine-treated patients with vs. without RTK-RAS or MAPK pathway alterations).

### 2.5 Conversational AI-enabled analytic workflow and validation

To accelerate cohort construction, stratified querying, and pathway-level aggregation, we deployed a conversational artificial intelligence workflow using AI-HOPE and the pathway-specialized modules AI-HOPE-RTK-RAS (35) and AI-HOPE-MAPK (36). These agents were used to (i) generate reproducible, multi-parameter cohorts via natural language queries (e.g., age group, gemcitabine exposure and pathway alteration status), (ii) compute stratified mutation frequency summaries at pathway and gene levels, and (iii) prioritize candidate age- and treatment-associated alterations for confirmatory testing. All AI-derived cohort counts, gene-level frequencies, and survival groupings were independently cross-validated against conventional data extracts and statistical analyses to ensure methodological transparency, reproducibility, and concordance with standard bioinformatic workflows

## 3. Results

### 3.1 Cohort composition and baseline clinical characteristics

Baseline demographic, clinical, and molecular characteristics of PDAC cohort are summarized in Table 1. The final analytic dataset included 184 patients with histopathologically confirmed pancreatic ductal adenocarcinoma, all derived from primary tumor specimens.

**Table 1.** Summary of Baseline Demographic, Clinical, and Genomic Features of the Study Population.

| Clinical Feature | PAAD Cohort n (%) |
| --- | --- |
| Age Onset & Treatment |  |
| Early-Onset (< 50) Treated with Gemcitabine | 15 (8.2%) |
| Late-Onset ($\geq 50$) Treated with Gemcitabine | 91 (49.5%) |
| Early-Onset ($< 50$) Not Treated with Gemcitabine | 5 (2.7%) |
| Late-Onset ($\geq 50$) Not Treated with Gemcitabine | 73 (39.7%) |
| **Cancer Type** |  |
| Pancreatic Adenocarcinoma | 184 (100.0%) |
| **Sex** |  |
| Male | 101 (54.9%) |
| Female | 83 (45.1%) |
| **Sample Type** |  |
| Primary Tumor | 184 (100.0%) |
| **Stage at Diagnosis** |  |
| Stage I | 21 (11.4%) |
| Stage II | 151 (82.1%) |
| Stage III | 5 (2.7%) |
| Stage IV | 5 (2.7%) |
| NA | 2 (1.1%) |
| **Ethnicity** |  |
| Hispanic or Latino | 5 (2.7%) |
| Not Hispanic or Latino | 136 (73.9%) |
| Unknown | 43 (23.4%) |

When stratified by age at diagnosis and gemcitabine exposure, late-onset PDAC (≥50 years) treated with gemcitabine constituted the largest subgroup (49.5%), followed by late-onset PDAC cases not treated with gemcitabine (39.7%). Early-onset PDAC (<50 years) represented a smaller proportion of the cohort overall, accounting for 8.2% of gemcitabine-treated and 2.7% of non-gemcitabine-treated cases. These distributions reflect the predominance of late-onset disease and the widespread use of gemcitabine-based therapy in this clinical context.

Sex distribution was balanced, with males representing 54.9% and females 45.1% of cases. All tumors were profiled from primary lesions, minimizing variability related to metastatic sampling.

At diagnosis, most patients presented with stage II disease (82.1%), while stage I tumors comprised 11.4% of cases. Advanced-stage presentations (stage III and IV) were less frequent (2.7% each), and staging information was unavailable in 1.1% of cases. Ethnicity annotations indicated that the majority of patients were categorized as not Hispanic or Latino (73.9%), with 2.7% identified as Hispanic or Latino and 23.4% classified as unknown. Together, these baseline features provide the clinical framework for subsequent age- and treatment-stratified analyses of RTK-RAS and MAPK pathway alterations.

### 3.2 Distribution of RTK-RAS and MAPK Pathway Alterations Across Age and Gemcitabine Context

To evaluate whether pathway-level alteration frequencies differed by age at diagnosis or gemcitabine exposure, we compared RTK-RAS and MAPK pathway status across stratified PDAC subgroups (Table 2a-d). In contrast to the gene-specific enrichments described later, pathway-level alteration prevalence was broadly consistent across age and treatment categories.

**Table 2.** Distribution of RTK-RAS and MAPK Pathway Alterations Across Age- and Gemcitabine-Stratified PDAC Subgroups. This table presents the prevalence of RTK-RAS and MAPK pathway alterations across PDAC cohorts stratified by age at diagnosis (early-onset vs. late-onset) and gemcitabine exposure. Results are organized into four analytical panels: (a) RTK-RAS pathway alterations comparing gemcitabine-treated versus non-treated tumors within early-onset disease; (b) RTK-RAS pathway alterations comparing early- versus late-onset disease within each treatment category; (c) MAPK pathway alterations comparing gemcitabine-treated versus non-treated tumors within early-onset disease; and (d) MAPK pathway alterations comparing early- versus late-onset disease within each treatment stratum. This structure enables pathway-specific, age-dependent, and treatment-contextual interpretation of molecular differences.

| Pathway Alterations | Early-Onset<br>Treated<br>with<br>Gemcitabine<br>n (%) | Early-Onset<br>Not<br>Treated<br>with<br>Gemcitabine<br>n (%) | p-value | Late-Onset<br>Treated<br>with<br>Gemcitabine<br>n (%) | Late-Onset<br>Not<br>Treated<br>with<br>Gemcitabine<br>n (%) | p-value |
| --- | --- | --- | --- | --- | --- | --- |
| RTK/RAS Alterations<br>Present | 11 (73.3%) | 4 (80.0%) | 1 | 59 (64.8%) | 49 (67.1%) | 0.8875 |
| RTK/RAS Alterations<br>Absent | 4 (26.7%) | 1 (20.0%) |  | 32 (35.2%) | 24 (32.9%) |  |

b)
| Pathway Alterations | Early-Onset<br>Treated<br>with<br>Gemcitabine<br>n (%) | Late-Onset<br>Treated<br>with<br>Gemcitabine<br>n (%) | p-value | Early-Onset<br>Not<br>Treated<br>with<br>Gemcitabine<br>n (%) | Late-Onset<br>Not<br>Treated<br>with<br>Gemcitabine<br>n (%) | p-value |
| --- | --- | --- | --- | --- | --- | --- |
| RTK/RAS Alterations<br>Present | 11 (73.3%) | 59 (64.8%) | 0.7693 | 4 (80.0%) | 49 (67.1%) | 1 |
| RTK/RAS Alterations<br>Absent | 4 (26.7%) | 32 (35.2%) |  | 1 (20.0%) | 24 (32.9%) |  |

c)
| Pathway Alterations | Early-Onset<br>Treated<br>with<br>Gemcitabine<br>n (%) | Early-Onset<br>Not<br>Treated<br>with<br>Gemcitabine<br>n (%) | p-value | Late-Onset<br>Treated<br>with<br>Gemcitabine<br>n (%) | Late-Onset<br>Not<br>Treated<br>with<br>Gemcitabine<br>n (%) | p-value |
| --- | --- | --- | --- | --- | --- | --- |
| MAPK Alterations<br>Present | 13 (86.7%) | 4 (80.0%) | 1 | 76 (83.5%) | 55 (75.3%) | 0.2706 |
| MAPK Alterations<br>Absent | 2 (13.3%) | 1 (20.0%) |  | 15 (16.5%) | 18 (24.7%) |  |

d)
| Pathway Alterations | Early-Onset Treated with Gemcitabine n (%) | Late-Onset Treated with Gemcitabine n (%) | p-value | Early-Onset Not Treated with Gemcitabine n (%) | Late-Onset Not Treated with Gemcitabine n (%) | p-value |
| --- | --- | --- | --- | --- | --- | --- |
| MAPK Alterations Present | 13 (86.7%) | 76 (83.5%) | 1 | 4 (80.0%) | 55 (75.3%) | 1 |
| MAPK Alterations Absent | 2 (13.3%) | 15 (16.5%) |  | 1 (20.0%) | 18 (24.7%) |  |

#### 3.2.1 RTK-RAS pathway alterations by age and treatment

Within EOPDAC, RTK-RAS pathway alterations were present in 73.3% of gemcitabine-treated tumors and 80.0% of non-treated tumors (p=1), indicating no significant treatment-associated difference. A similar pattern was observed in late-onset PDAC, where alterations were detected in 64.8% of gemcitabine-treated and 67.1% of non-treated tumors (p=0.8875) (Table 2a).

Direct age-based comparisons within treatment strata likewise showed no statistically significant differences. Among gemcitabine-treated patients, RTK-RAS alterations occurred in 73.3% of early-onset versus 64.8% of late-onset PDAC tumors (p=0.7693). In the non-treated setting, alteration frequencies were 80.0% in early-onset PDAC and 67.1% in late-onset PDAC cases (p=1) (Table 2b). These findings suggest that, at the pathway level, RTK-RAS disruption is common across PDAC and does not vary substantially by age or gemcitabine exposure.

#### 3.2.2 MAPK pathway alterations by age and treatment

MAPK pathway alterations were highly prevalent across all subgroups. In early-onset PDAC, 86.7% of gemcitabine-treated and 80.0% of non-treated tumors harbored MAPK alterations (p=1). In LOPDAC disease, prevalence was 83.5% in treated and 75.3% in non-treated cases (p=0.2706) (Table 2c).

Age-based comparisons within treatment categories similarly demonstrated comparable frequencies. Among gemcitabine-treated tumors, MAPK alterations were present in 86.7% of early-onset PDAC and 83.5% of late-onset PDAC cases (p=1). In non-treated tumors, frequencies were 80.0% and 75.3%, respectively (p=1) (Table 2d).

These results indicate that both RTK-RAS and MAPK pathway alterations are widespread features of PDAC, largely independent of age at onset or gemcitabine exposure. This relative uniformity at the pathway level contrasts with the age- and treatment-dependent gene-specific differences observed in subsequent analyses, underscoring the importance of dissecting pathway architecture beyond aggregate alteration status.

### 3.3 Gene-level architecture of RTK-RAS and MAPK pathway alterations across age and gemcitabine exposure

Because pathway-level alteration rates were broadly similar across strata (Section 3.2), we next resolved RTK-RAS and MAPK signaling dysregulation at single-gene resolution to identify age-dependent dependencies and treatment-contextual enrichment patterns (Tables S1-S8). Across the cohort, KRAS represented the dominant RTK-RAS node in both early- and late-onset PDAC and in both gemcitabine-exposed and non-exposed tumors, whereas most canonical receptor and kinase alterations occurred at low frequency. Notably, several non-canonical but potentially actionable alterations emerged in treatment- and age-restricted contexts, revealing molecular substructures that were not apparent from aggregate pathway status.

#### 3.3.1 RTK-RAS gene-level landscape highlights gemcitabine-associated RTK enrichment in late-onset PDAC

In early-onset PDAC, the RTK-RAS landscape was largely KRAS-centric. KRAS mutations were present in 73.3% of gemcitabine-treated early-onset tumors and 80.0% of non-treated tumors (Table S1), with no meaningful difference by treatment status. Beyond KRAS, RTK alterations were rare in early-onset disease; for example, NTRK1 and CBL mutations were each observed in 6.7% of gemcitabine-treated tumors and absent in non-treated early-onset tumors (Table S1), consistent with sporadic, low-prevalence events.

In contrast, late-onset PDAC demonstrated treatment-contextual enrichment of select RTKs within gemcitabine-exposed tumors. ERBB2 mutations were detected in 6.6% of gemcitabine-treated late-onset tumors and 0% of non-treated late-onset tumors (p=0.034) (Table S2). Similarly, RET mutations occurred in 7.7% of gemcitabine-treated late-onset tumors and 0% of non-treated late-onset tumors (p=0.0175) (Table S2). Additional RTK/RAS-axis genes (e.g., EGFR, ERBB3, FGFR3, MET, ROS1, ALK) appeared at low frequency among treated late-onset tumors but did not reach statistical significance versus non-treated disease (Table S2). Importantly, KRAS mutation rates were comparable between treated and non-treated late-onset tumors (65.9% vs. 63.0%, p=0.8224) (Table S2), indicating that the ERBB2/RET signals were not driven by differences in KRAS prevalence.

Age-based comparisons within the gemcitabine-treated setting did not show significant differences for ERBB2 or RET between early- and late-onset tumors (Table S3), reflecting limited power in the early-onset treated subset; however, the late-onset within-group contrast against non-treated tumors supported a treatment-associated enrichment pattern for these RTKs (Table S2). Together, these results suggest that gemcitabine exposure in late-onset PDAC coincides with a measurable shift toward RTK-level diversity (ERBB2, RET) superimposed on a persistent KRAS-driven backbone. Comparison of early-onset versus late-onset PDAC patients not treated with gemcitabine did not reveal any statistically significant differences (Table S4).

#### 3.3.2 MAPK gene-level landscape reveals age- and treatment-dependent non-canonical dependencies

Within the MAPK signaling axis, early-onset PDAC again showed a restricted set of recurrent events in gemcitabine-treated tumors, but with notable age- and treatment-sensitive features. In gemcitabine-treated early-onset disease, TP53 mutations were present in 86.7% of tumors, compared with 40.0% in non-treated early-onset tumors (Table S5), representing a strong trend that did not meet statistical significance in this small subgroup (p=0.0726). Moreover, cytoskeletal MAPK-linked modulators demonstrated early-onset specificity under gemcitabine exposure: FLNB mutations occurred in 13.3% of gemcitabine-treated early-onset tumors and 0% of treated late-onset tumors (p=0.0189) (Table S7), consistent with an early-onset, treatment-contextual dependency at this node. TP53 was also significantly more frequent in gemcitabine-treated early-onset versus treated late-onset tumors (86.7% vs. 57.1%, p=0.0431) (Table S7), reinforcing age-specific genomic architecture among treated patients.

A distinct pattern emerged in the non-gemcitabine setting, where early-onset tumors exhibited striking enrichment of MAPK-associated calcium channel genes. CACNA2D1, CACNA2D3, and CACNA2D4 mutations were each present in 40.0% of non-treated early-onset tumors versus 0% of gemcitabine-treated early-onset tumors, showing borderline treatment-associated differences within early-onset disease (p=0.0526 for each) (Table S5). When early- versus late-onset disease was compared within the non-treated stratum, these same genes remained strongly age-enriched: CACNA2D1/3/4 were each present in 40.0% of early-onset non-treated tumors versus 1.4% of late-onset non-treated tumors (p=0.0097 for each) (Table S8). These findings indicate that, in the absence of gemcitabine exposure, early-onset PDAC is characterized by a distinct, non-canonical MAPK-linked signature involving CACNA2D-family alterations.

In late-onset PDAC, MAPK-associated alterations were distributed across multiple low-frequency nodes, with a subset showing treatment-contextual differences. Among treated late-onset tumors, CACNA1C (9.9% vs. 0%, p=0.0047) and CACNA1E (7.7% vs. 0%, p=0.0175) were significantly more frequent compared with non-treated late-onset tumors (Table S6). Additionally, TGFBR2 mutations were enriched in treated late-onset tumors (8.8% vs. 1.4%, p=0.0439) (Table S6), suggesting broader signaling cross-talk that may co-occur with MAPK pathway perturbation in the treated setting.

#### 3.3.3 Integrated interpretation

Collectively, gene-level dissection clarified that the apparent stability of pathway-level alteration prevalence (Section 3.2) masks substantial age- and gemcitabine-dependent re-wiring at specific nodes. Late-onset gemcitabine-treated PDAC was distinguished by enrichment of RTK alterations (ERBB2, RET) within the RTK-RAS axis, while early-onset disease, particularly under gemcitabine exposure, showed higher rates of TP53 and FLNB alterations. Conversely, early-onset non-treated tumors displayed a pronounced CACNA2D-family alteration signature, indicating a potentially distinct MAPK-linked dependency structure in the absence of gemcitabine.

### 3.4 Overall Survival According to RTK-RAS Pathway Alteration Status

We next evaluated the prognostic significance of RTK-RAS pathway alterations across age- and gemcitabine-stratified PDAC subgroups using Kaplan-Meier analyses (Figure 1a-d).

**Figure 1.**
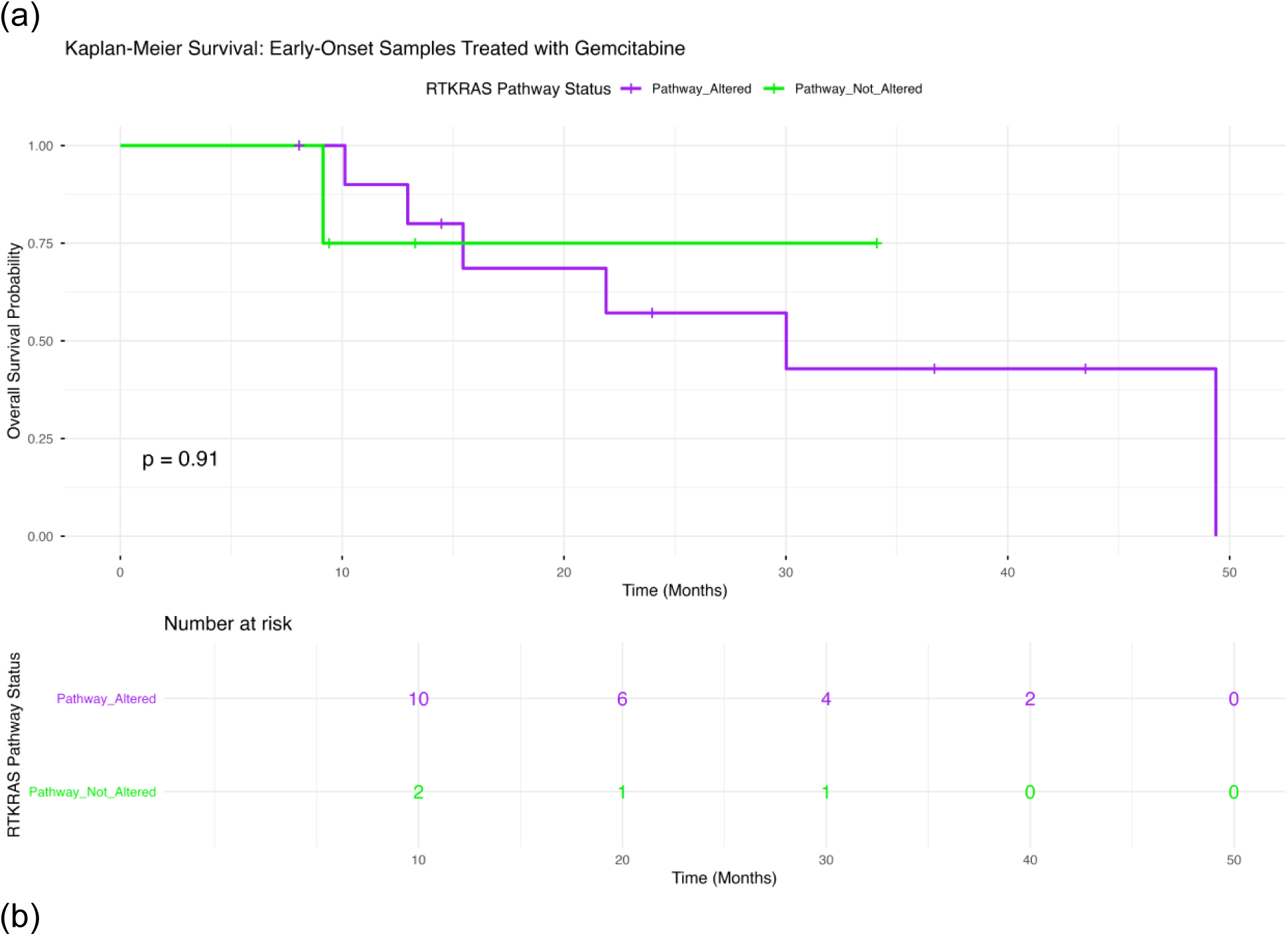
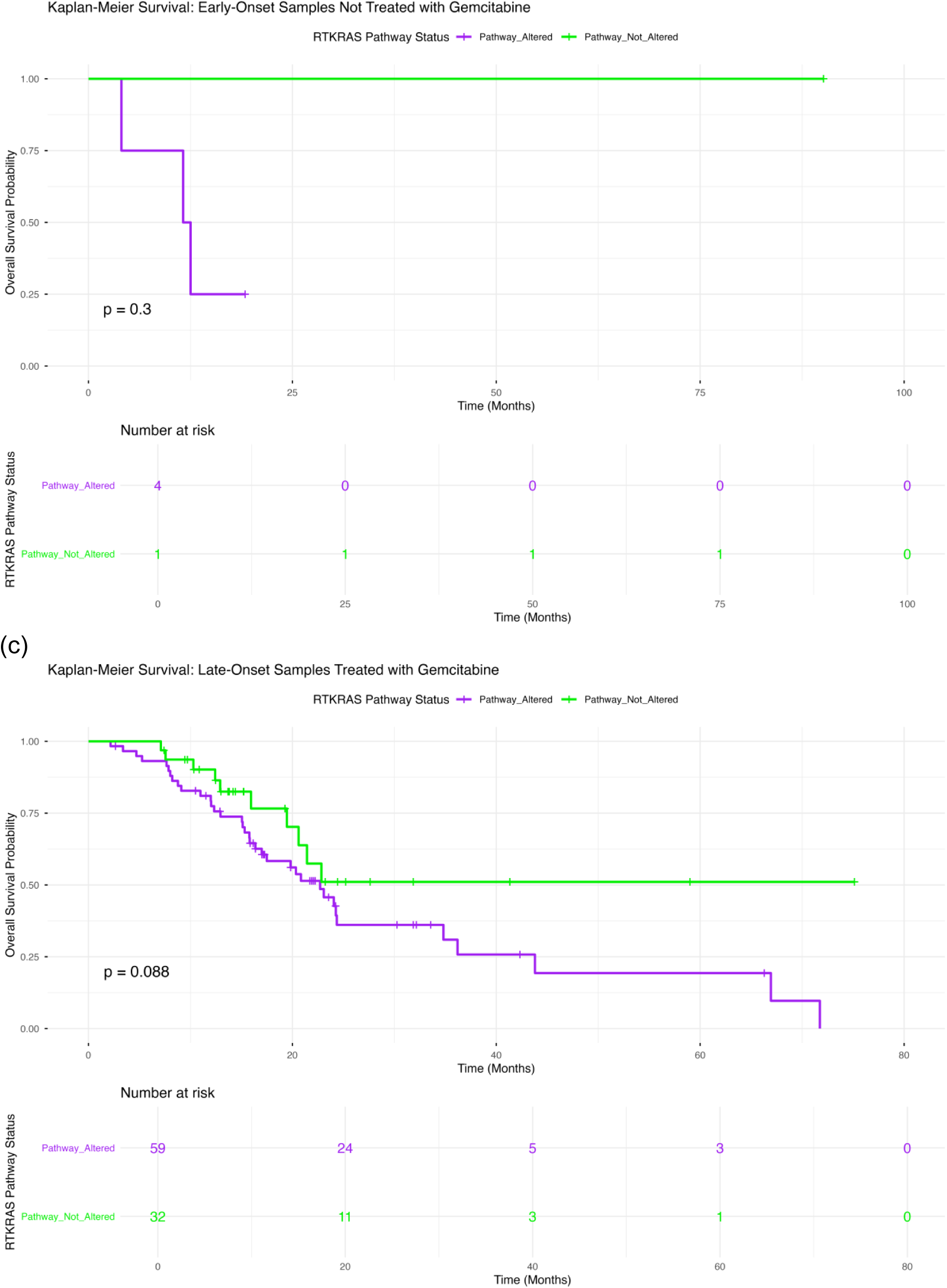
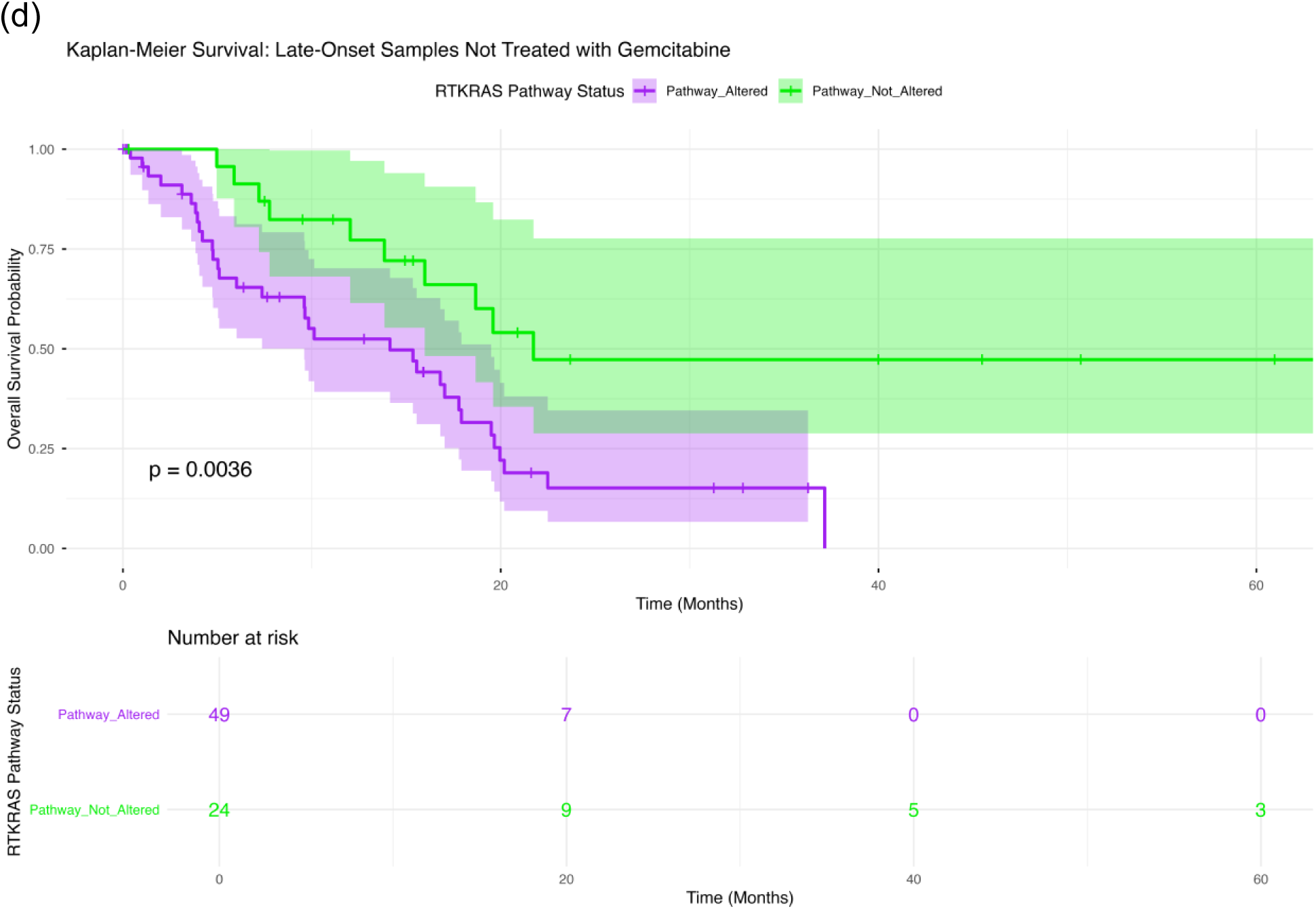
Overall survival by RTK-RAS pathway alteration status across age- and gemcitabine-stratified PDAC subgroups. Kaplan-Meier curves compare overall survival between patients whose tumors harbored RTK-RAS pathway alterations and those without detectable RTK-RAS alterations, stratified by age at diagnosis and gemcitabine exposure. Panels depict (a) early-onset PDAC (<50 years) treated with gemcitabine, (b) early-onset PDAC not treated with gemcitabine, (c) late-onset PDAC (≥50 years) treated with gemcitabine, and (d) late-onset PDAC not treated with gemcitabine. Shaded bands indicate 95% confidence intervals, and tables beneath each panel show numbers at risk over time.

#### 3.4.1 Early-Onset PDAC Treated with Gemcitabine

Among early-onset patients receiving gemcitabine (Figure 1a), overall survival did not differ meaningfully according to RTK-RAS pathway status (p = 0.91). Survival trajectories for tumors with and without RTK-RAS alterations were largely overlapping throughout follow-up. Confidence intervals were wide, reflecting the relatively small sample size in this subgroup, and no consistent survival advantage was observed in either group.

#### 3.4.2 Early-Onset PDAC Not Treated with Gemcitabine

In early-onset patients not exposed to gemcitabine (Figure 1b), survival curves showed modest separation; however, the difference was not statistically significant (p = 0.30). The RTK-RAS-unaltered group demonstrated sustained survival across follow-up, whereas the altered group exhibited earlier decline. Interpretation is limited by small numbers at risk, particularly in the non-altered cohort.

#### 3.4.3 Late-Onset PDAC Treated with Gemcitabine

Among late-onset patients treated with gemcitabine (Figure 1c), a trend toward improved survival was observed in tumors lacking RTK-RAS alterations compared with altered tumors; however, this did not reach statistical significance (p = 0.088). While early survival probabilities were similar, divergence became more apparent at later time points, suggesting a potential interaction between pathway status and treatment context that warrants further evaluation.

#### 3.4.4 Late-Onset PDAC Not Treated with Gemcitabine

In contrast, late-onset patients not treated with gemcitabine (Figure 1d) demonstrated a statistically significant survival difference by RTK-RAS pathway status (p = 0.0036). Patients whose tumors lacked RTK-RAS alterations exhibited substantially improved overall survival compared with those harboring pathway mutations. The survival curves separated early and remained distinct over time, with non-altered tumors maintaining higher survival probabilities across the follow-up period.

These findings indicate that RTK-RAS pathway alteration status has context-dependent prognostic relevance in PDAC. While no significant survival impact was observed in early-onset disease or among gemcitabine-treated patients, the absence of RTK-RAS alterations identifies a favorable-prognosis subgroup within late-onset, non-gemcitabine-treated PDAC, underscoring the importance of integrating pathway status with age and treatment exposure in precision outcome modeling.

### 3.5 Overall Survival According to MAPK Pathway Alteration Status

We next examined the prognostic impact of MAPK pathway alterations across age-and gemcitabine-defined PDAC subgroups (Figure 2a-d), assessing whether pathway-level dysregulation stratifies survival outcomes in a treatment-contextual manner.

**Figure 2.**
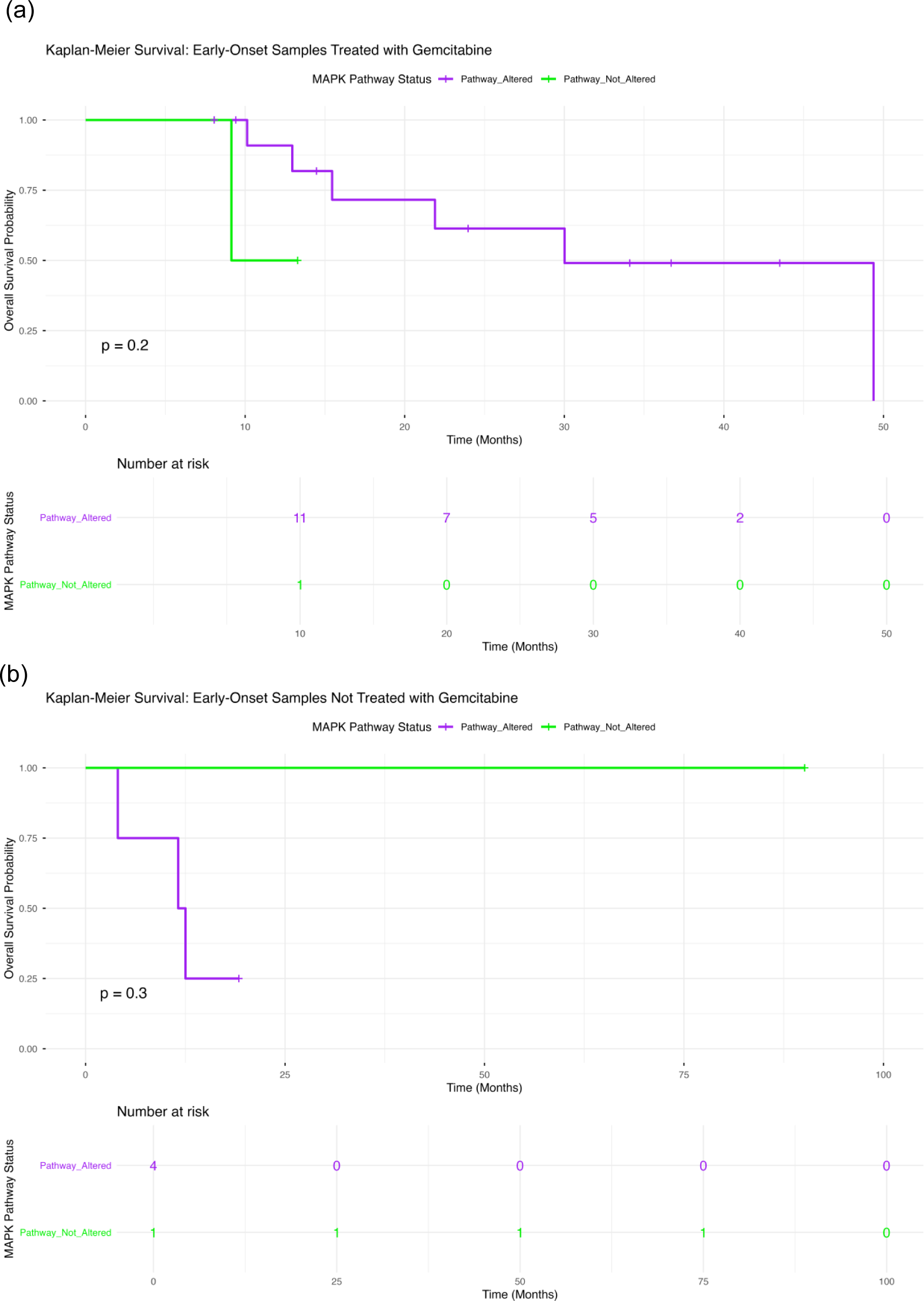
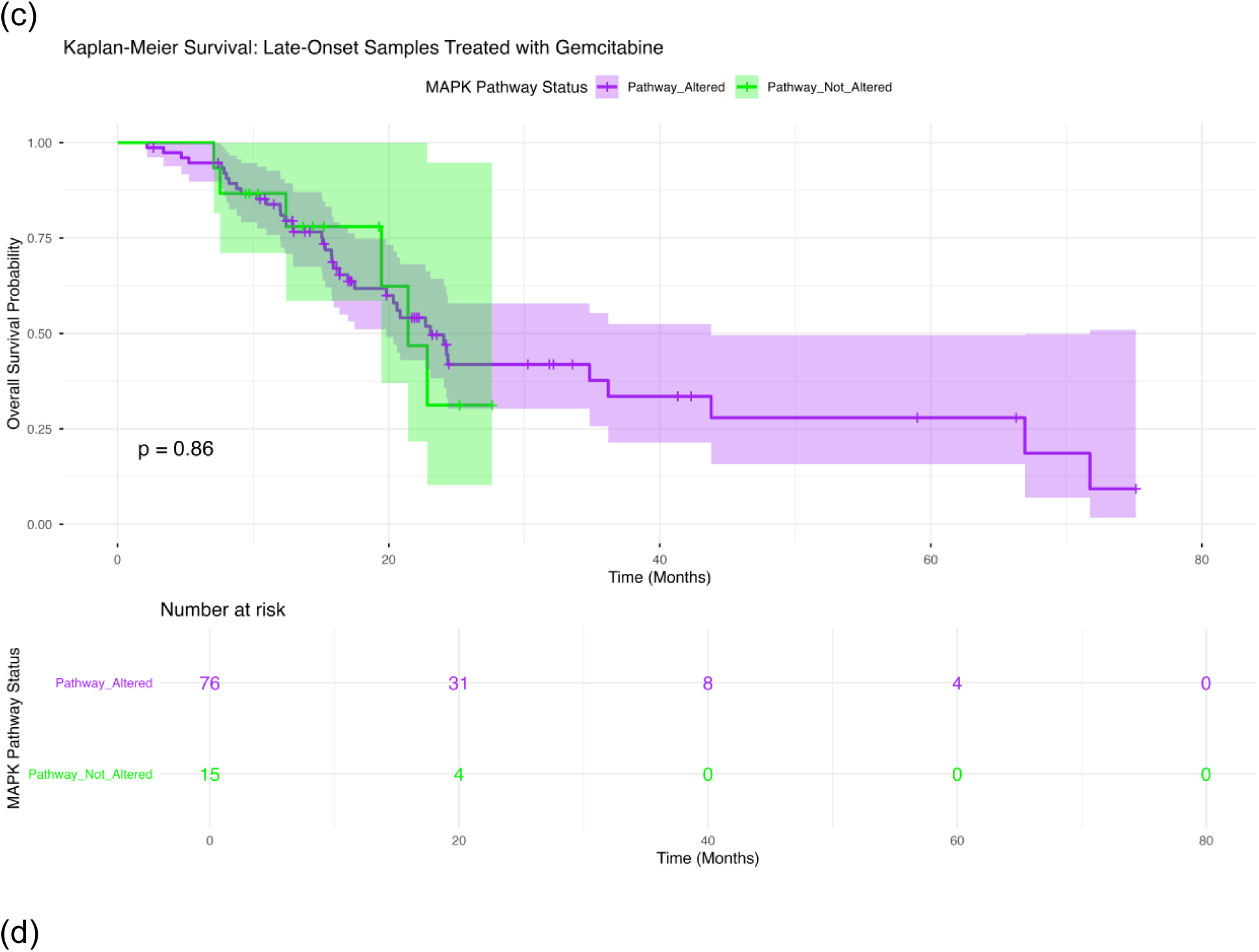
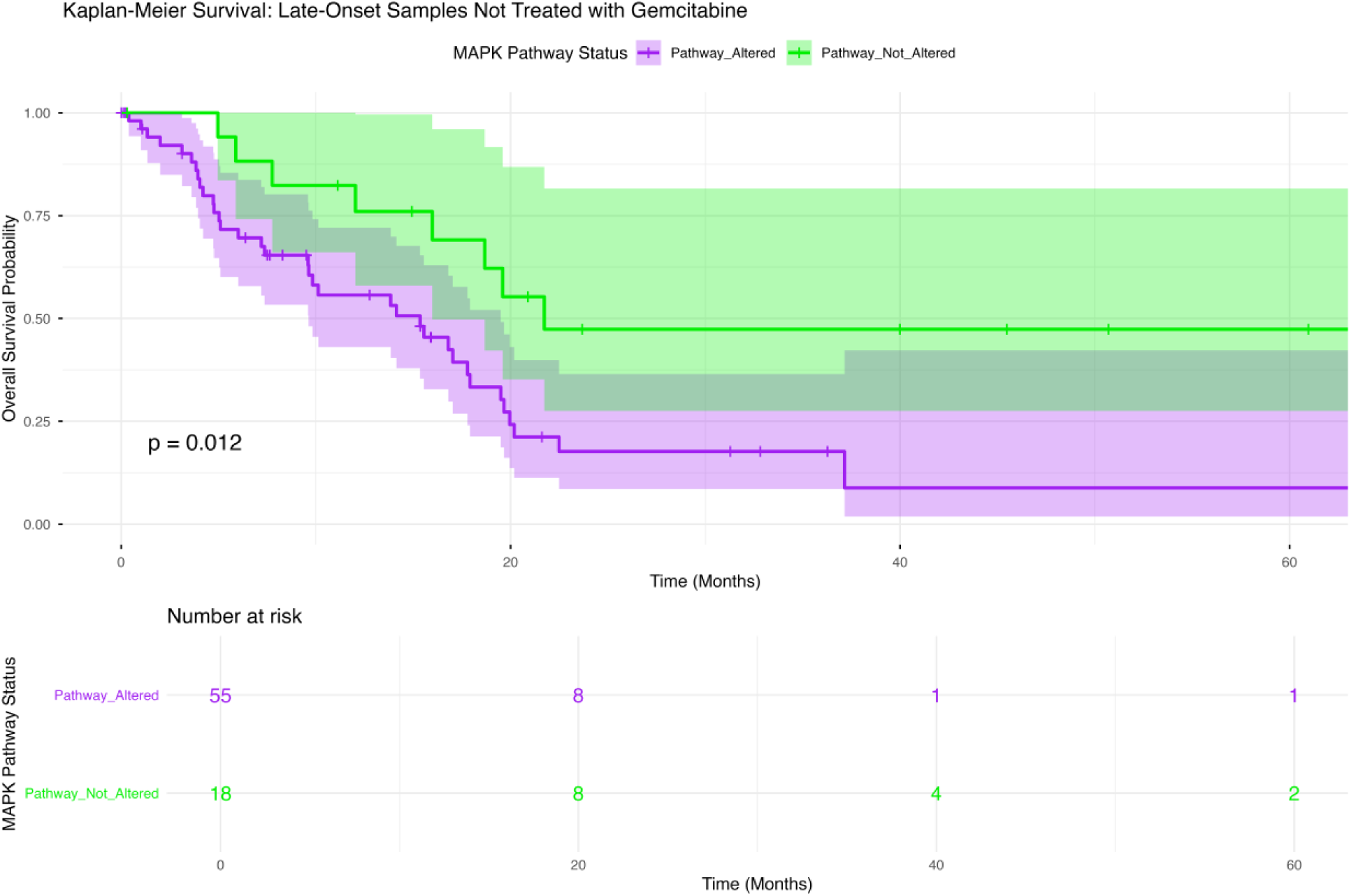
Overall survival according to MAPK pathway alteration status across age-and gemcitabine-stratified PDAC subgroups. Kaplan-Meier survival curves illustrate overall survival differences between patients with tumors harboring MAPK pathway alterations and those without detectable MAPK alterations, stratified by age at diagnosis and gemcitabine exposure. Panels include: (a) early-onset PDAC (<50 years) treated with gemcitabine; (b) early-onset PDAC not treated with gemcitabine; (c) late-onset PDAC (≥50 years) treated with gemcitabine; and (d) late-onset PDAC not treated with gemcitabine. Within each subgroup, survival distributions for MAPK-altered versus MAPK-wild-type tumors are compared. Shaded areas represent 95% confidence intervals, and accompanying risk tables display the number of patients at risk over time, facilitating interpretation of temporal survival patterns within each clinically defined stratum.

#### 3.5.1 Early-Onset PDAC Treated with Gemcitabine

In early-onset patients receiving gemcitabine (Figure 2a), overall survival did not significantly differ by MAPK pathway status (p = 0.20). Although tumors harboring MAPK alterations demonstrated a gradual decline in survival over time, the non-altered group as small and showed wide confidence intervals. The survival curves overlapped substantially during early follow-up, and no durable separation was observed.

#### 3.5.2 Early-Onset PDAC Not Treated with Gemcitabine

Among early-onset patients who did not receive gemcitabine (Figure 2b), survival comparisons were likewise not statistically significant (p = 0.30). The MAPK-wild-type group maintained prolonged survival, while the altered cohort showed earlier attrition; however, the small number of non-treated early-onset cases limited statistical power and precision of estimates.

#### 3.5.3 Late-Onset PDAC Treated with Gemcitabine

In late-onset patients treated with gemcitabine (Figure 2c), survival trajectories for MAPK-altered and non-altered tumors were nearly superimposed (p = 0.86). Both groups exhibited comparable early and intermediate survival probabilities, and confidence intervals overlapped extensively throughout follow-up. These findings suggest that MAPK pathway status does not independently stratify prognosis among gemcitabine-treated late-onset PDAC.

#### 3.5.4 Late-Onset PDAC Not Treated with Gemcitabine

In contrast, a significant survival difference emerged in late-onset patients not exposed to gemcitabine (Figure 2d). Tumors lacking MAPK pathway alterations were associated with improved overall survival compared with MAPK-altered tumors (p = 0.012). The survival curves separated early and remained distinct over time, with the MAPK-wild-type subgroup maintaining higher survival probabilities throughout follow-up.

These analyses indicate that MAPK pathway alterations confer context-dependent prognostic relevance in PDAC. While no significant survival associations were observed in early-onset disease or in gemcitabine-treated subgroups, the absence of MAPK pathway alterations identifies a favorable-prognosis subset among late-onset, non-gemcitabine-treated patients, reinforcing the importance of integrating pathway status with age and therapeutic exposure in precision outcome modeling.

### 3.6 Conversational Artificial Intelligence-Driven Exploratory Analyses

#### 3.6.1 AI-HOPE-RTK-RAS: Dynamic Cohort Construction and Pathway-Centric Profiling

To complement pathway-specific statistical modeling, we deployed the AI-HOPE-RTK-RAS agent to perform structured, natural language-guided exploratory analyses within integrated PDAC clinical-genomic datasets. This framework enabled real-time cohort definition, mutation frequency comparisons, and multidimensional association testing under strictly user-defined eligibility criteria.

In an initial proof-of-concept demonstration (Figure S1), the agent constructed treatment- and age-matched cohorts and executed survival comparisons based on structured filters. The system dynamically parsed inclusion rules, generated case and control groups, and computed Kaplan-Meier estimates with associated log-rank statistics. This workflow confirmed the feasibility of automated survival interrogation under tightly controlled clinical contexts, establishing the foundation for subsequent RTK-RAS-focused exploration.

We next interrogated treatment-contextual molecular stability using AI-driven 2×2 contingency analyses (Figure S2). The AI agent isolated gemcitabine-defined strata and compared predefined genomic attributes across cohorts. Odds ratio testing demonstrated no significant association between treatment exposure and the selected genomic stability metric, illustrating the platform’s ability to rapidly validate null hypotheses in clinically constrained subsets.

Similarly, targeted mutation frequency analysis (Figure S3) assessed whether a candidate signaling gene exhibited treatment-associated enrichment. The conversational interface constructed gemcitabine-stratified groups and performed Fisher’s exact testing, revealing no statistically significant difference in mutation prevalence between exposure categories. These findings underscore the importance of treatment-contextual validation before attributing biological significance to pathway-level differences.

We then expanded the AI interrogation to a broader, pathway-centric association analysis (Figure S4), comparing RTK-RAS pathway-altered tumors (n = 123) with pathway-non-altered tumors (n = 61). Automated categorical and continuous statistical testing identified strong associations with canonical KRAS mutation status and MAPK pathway involvement, as anticipated. Notably, RTK-RAS-altered tumors demonstrated enrichment for TP53 alterations, SMAD4 and CDKN2A events, and TGFβ pathway involvement. Continuous genomic instability measures, including nonsynonymous tumor mutation burden, mutation count, fraction of genome altered, aneuploidy score, and multiple hypoxia signatures, were significantly elevated in the RTK-RAS-altered cohort. Clinical outcome variables were also associated with pathway status. Collectively, these AI-derived signals reveal a coordinated genomic instability and microenvironmental phenotype linked to RTK-RAS dysregulation in PDAC.

#### 3.6.2 AI-HOPE-MAPK: Treatment-Contextual MAPK Interrogation

We subsequently deployed the AI-HOPE-MAPK agent to interrogate MAPK pathway dependencies within gemcitabine-defined contexts (Figures S5-S9).

First, the agent constructed late-onset PDAC cohorts stratified by gemcitabine exposure (Figure S5). Kaplan-Meier analysis demonstrated a statistically significant survival difference between treated and untreated groups (log-rank p = 0.0114), confirming that the conversational AI workflow can reproduce clinically meaningful treatment-outcome relationships in PDAC. This served as a validation step prior to deeper pathway-specific interrogation.

We then evaluated whether specific MAPK-adjacent genomic events differed by treatment exposure. Analysis of TGFBR2 mutation frequency (Figure S6) revealed a numerically higher prevalence among gemcitabine-treated patients (6.59%) compared with untreated patients (1.37%); however, this difference did not reach statistical significance (p = 0.209). This illustrates how AI-guided testing can distinguish true enrichment from stochastic variation in relatively small cohorts.

Age-stratified mutation comparisons within gemcitabine-treated PDAC further demonstrated the agent’s contextual flexibility. FLNB mutation prevalence (Figure S7) was modestly higher in early-onset treated tumors than in late-onset treated tumors, yet this difference was not statistically significant. In contrast, TP53 mutation analysis (Figure S8) showed a higher numerical frequency in early-onset treated tumors (80.0%) compared with late-onset treated tumors (56.0%), though again without statistical significance. These analyses emphasize that while numerical trends may suggest biological divergence, rigorous statistical testing within AI-defined cohorts is essential to avoid overinterpretation.

Finally, we performed a comprehensive association analysis comparing gemcitabine-treated MAPK-altered tumors (n = 89) with gemcitabine-treated MAPK-non-altered tumors (n = 17) (Figure S9). The AI-HOPE-MAPK agent integrated Chi-square and Mann-Whitney U testing to identify significant categorical and continuous attributes associated with MAPK alteration status. MAPK-altered tumors exhibited strong co-association with RTK-RAS pathway involvement, KRAS mutations, and TP53 pathway alterations. Measures of genomic instability, including tumor mutation burden, total mutation count, tumor break load, fraction genome altered, and aneuploidy, were significantly enriched. Hypoxia signatures (Winter, Buffa, and Ragnum scores) were also elevated in MAPK-altered tumors, alongside associations with TGFβ pathway and SMAD4 alterations.

Together, these conversational AI-driven analyses reveal that MAPK pathway alterations in gemcitabine-treated PDAC are embedded within a broader genomic instability and stress-response landscape. Importantly, the AI-HOPE-RTK-RAS and AI-HOPE-MAPK agents enabled rapid, reproducible generation of treatment-contextual cohorts, survival modeling, mutation enrichment testing, and multidimensional association discovery. This integrated workflow demonstrates the practical utility of conversational AI as a scalable analytical layer for pathway-dependent precision oncology in pancreatic ductal adenocarcinoma.

## 4. Discussion

Pancreatic ductal adenocarcinoma (PDAC) remains a therapeutically refractory malignancy in which canonical oncogenic signaling, most prominently KRAS-driven RTK-RAS/MAPK activation, coexists with marked inter-patient heterogeneity and treatment-contextual adaptation (1,2,39–45). In this study, we used a conversational artificial intelligence framework, AI-HOPE-RTK-RAS (35) and AI-HOPE-MAPK (36), to dissect pathway-level and gene-level architecture of RTK-RAS and MAPK alterations in a clinically annotated PDAC cohort stratified by age at onset and gemcitabine exposure. Three overarching insights emerge: (i) pathway alteration prevalence is uniformly high across strata, (ii) gene-level “rewiring” reveals age- and treatment-dependent dependencies that are obscured by aggregate pathway status, and (iii) prognostic associations of pathway alterations are context-dependent, with the strongest survival separation observed in late-onset PDAC not treated with gemcitabine.

### 4.1 Pathway prevalence is stable, but pathway architecture is not

At the pathway level, RTK-RAS and MAPK alterations were common across all age and gemcitabine strata (Section 3.2), consistent with PDAC’s KRAS-centric biology. Importantly, this apparent uniformity could have been misinterpreted as biological equivalence across contexts. However, gene-level resolution (Section 3.3) demonstrated that pathway “membership” is not synonymous with pathway “dependency.” In other words, while most tumors are classified as RTK-RAS/MAPK altered, the specific nodes contributing to pathway dysregulation differ by age and gemcitabine exposure, indicating that PDAC signaling architecture is modular and context-sensitive.

This distinction has practical implications for precision oncology. Pathway-level classifiers can support broad mechanistic framing, but they may not adequately guide actionable targeting or biomarker strategies. Our findings argue for pathway dissection at node-level granularity, especially when the clinical question is treatment-specific (gemcitabine exposure) or population-specific (early- vs. late-onset PDAC).

### 4.2 Late-onset gemcitabine-treated PDAC shows RTK diversification on a KRAS backbone

Within the RTK-RAS axis, KRAS remained dominant across groups, yet late-onset gemcitabine-treated tumors showed enrichment of select receptor tyrosine kinases, particularly ERBB2 and RET (Section 3.3.1). The observation that KRAS prevalence was similar in treated and untreated late-onset tumors argues against a trivial explanation that RTK enrichment simply reflects differential KRAS rates. Instead, these data suggest that gemcitabine exposure in late-onset PDAC may coincide with (or select for) a broader RTK repertoire, potentially reflecting adaptive signaling, clonal selection, or treatment-associated tumor evolutionary pressures.

From a translational standpoint, the emergence of ERBB2 and RET alterations in a treatment-defined context is notable because these are, in principle, more directly targetable than many other PDAC lesions. Although the current cohort does not establish causality, the data support a testable hypothesis: gemcitabine-treated late-onset PDAC may contain an RTK-enriched subset where receptor-level targeting (or RTK-MAPK co-targeting strategies) could be most rationally explored. Future studies should evaluate whether these RTK events represent truncal drivers, subclonal adaptations, or therapy-associated expansions and whether they correlate with response or resistance trajectories.

### 4.3 Early-onset PDAC exhibits distinct MAPK-linked non-canonical signatures

In MAPK-centered analyses, early-onset PDAC demonstrated patterns that diverge from the late-onset landscape. Under gemcitabine exposure, early-onset tumors showed higher TP53 mutation frequency than late-onset treated disease and enrichment of FLNB alterations (Section 3.3.2). FLNB’s emergence as an early-onset, gemcitabine-contextual feature suggests that cytoskeletal and mechanotransduction-related biology may intersect with MAPK signaling in age-specific ways. While FLNB is not a canonical MAPK node, its association in this context raises the possibility that early-onset PDAC may exhibit distinct invasion, stress-adaptation, or microenvironment-interaction programs that become clinically relevant under chemotherapy pressure.

Conversely, in the non-gemcitabine early-onset subset, CACNA2D-family alterations (CACNA2D1/3/4) were strikingly enriched relative to late-onset non-treated tumors (Section 3.3.2). These genes, linked to calcium channel regulation, suggest an alternative, non-canonical signaling configuration in early-onset PDAC that could influence cellular excitability, stress signaling, and downstream kinase cascades that interface with MAPK outputs. While sample size in early-onset non-treated tumors is limited, the magnitude and consistency of CACNA2D-family enrichment across comparisons indicates a biologically intriguing axis that warrants confirmation in larger EO cohorts and functional follow-up. Collectively, these age-dependent signals reinforce the concept that “early-onset PDAC” is not simply PDAC diagnosed earlier, but may represent a subset with distinct molecular wiring.

### 4.4 Prognostic relevance of RTK-RAS and MAPK alterations is context-dependent

Survival analyses further underscored the importance of age and treatment context (Sections 3.4-3.5). Across both RTK-RAS and MAPK pathways, the most robust survival separation was observed in late-onset patients not treated with gemcitabine, where pathway non-altered tumors had significantly improved overall survival (RTK-RAS p=0.0036; MAPK p=0.012). In contrast, no significant pathway-level survival effects were observed in early-onset disease or in gemcitabine-treated strata (with only a trend for RTK-RAS in late-onset treated patients).

Several interpretations are plausible. First, in gemcitabine-treated subgroups, treatment effects and clinical confounding (e.g., selection for therapy based on performance status, stage nuances, or comorbidity) may attenuate the apparent prognostic signal of pathway status. Second, given the near-ubiquity of pathway alterations in PDAC, the “non-altered” groups may represent biologically distinct tumors with alternative oncogenic drivers and potentially different natural histories, an effect that becomes most visible when not masked by therapy. Third, the late-onset non-treated cohort may capture a clinically heterogeneous group where absence of RTK-RAS/MAPK alterations correlates with less aggressive biology or differences in genomic instability, consistent with the association patterns observed in the AI-driven attribute analyses (Figures S4 and S9).

### 4.5 Conversational AI as an analytic layer for reproducible, context-aware precision oncology

A major contribution of this work is methodological: we demonstrate that conversational AI agents can be used as a practical analytic interface for cohort construction, stratified survival modeling, mutation enrichment testing, and association analyses under explicit user-defined rules (Section 3.6). Importantly, the AI-HOPE workflow operationalizes a common translational bottleneck: the need to iteratively refine cohorts and hypotheses across multiple clinical contexts (age, treatment and pathway status) without sacrificing reproducibility.

The supplemental analyses illustrate how the system can rapidly validate signals (e.g., survival differences by gemcitabine exposure in late-onset PDAC), flag non-significant trends that should not be overinterpreted (e.g., TGFBR2, FLNB, TP53 age-stratified frequency comparisons), and map pathway alterations to a broader phenotype of genomic instability and hypoxia signatures (Figures S4, S9). This combination of speed, transparency of inclusion criteria, and statistical execution supports the role of conversational AI as a scalable “analysis co-pilot” for precision oncology research, particularly valuable in exploratory studies spanning many stratified subgroups.

### 4.6 Limitations and future directions

This study has limitations inherent to retrospective, aggregated clinical-genomic datasets. First, early-onset PDAC subgroups were small, especially the non-gemcitabine early-onset group, limiting power and inflating uncertainty in subgroup survival curves and gene-level comparisons. Second, gemcitabine exposure is treated as a binary variable, but real-world therapy includes complex sequencing, combination regimens, dosing intensity, and lines of therapy; these factors can confound outcomes and may influence apparent enrichment patterns. Third, stage distribution was heavily weighted toward stage II, which may reflect cohort ascertainment and can constrain generalizability to metastatic PDAC. Fourth, pathway classification does not capture variant-level functional effects, clonality, or allelic dosage, which are critical to defining true dependencies.

Future work should prioritize: (i) validation in larger, independent PDAC cohorts with richer treatment metadata; (ii) integration of clonality and evolutionary timing for RTK events (e.g., ERBB2, RET) in gemcitabine-treated late-onset PDAC; (iii) functional investigation of early-onset signatures (CACNA2D-family alterations and FLNB) in MAPK-linked stress adaptation and chemotherapy response; and (iv) multivariable survival modeling incorporating stage, performance status proxies, and co-mutation burden to separate prognostic from predictive effects. Incorporating transcriptomic or proteomic readouts (e.g., pathway activity scores) would also help determine whether gene-level alterations translate into measurable signaling states and therapeutic vulnerabilities.

Another important limitation is the constrained demographic diversity and insufficient inclusion of underrepresented populations in existing aggregated PDAC datasets. In our cohort, Hispanic/Latino annotation was sparse and a substantial fraction of cases had unknown ethnicity, limiting the ability to evaluate ancestry-associated biology, treatment response heterogeneity, and inequities in actionable alteration patterns. Future studies should deliberately expand inclusion of Hispanic/Latino and African American/Black patients (and improve the completeness of race/ethnicity and social context annotations) to ensure that RTK–RAS/MAPK dependency maps and outcome models are generalizable and do not reinforce existing disparities (1,2,32,33). In parallel, deeper molecular phenotyping is needed to resolve PDAC heterogeneity beyond DNA alterations alone. Integrating spatial biology (spatial transcriptomics and spatial proteomics) (46) with multi-omics layers (bulk/single-cell RNA-seq, proteomics/phosphoproteomics, epigenomics, and immune/stromal profiling) (47,48) will be essential to define cell-type–specific pathway activation states, microenvironmental interactions, and therapy-adaptive programs that may explain the context-dependent signals observed here and uncover more precise, equity-aware therapeutic vulnerabilities.

## 5. Conclusions

In a clinically stratified PDAC cohort, RTK-RAS and MAPK pathway alterations were highly prevalent and largely invariant by age at onset and gemcitabine exposure at the aggregate pathway level. However, gene-level dissection revealed substantial context-specific pathway architecture: late-onset gemcitabine-treated PDAC showed enrichment of actionable RTK alterations (notably ERBB2 and RET) superimposed on persistent KRAS dominance, while early-onset PDAC exhibited distinct MAPK-linked non-canonical signatures, including FLNB enrichment in gemcitabine-treated disease and CACNA2D-family enrichment in non-treated disease. Survival analyses demonstrated that the prognostic relevance of pathway alterations is context-dependent, with the strongest survival advantage observed in late-onset patients not treated with gemcitabine whose tumors lacked RTK-RAS or MAPK alterations.

Methodologically, AI-HOPE-RTK-RAS and AI-HOPE-MAPK enabled rapid, reproducible, and context-aware cohort construction and statistical interrogation, highlighting conversational AI as a scalable analytic layer for pathway-centric precision oncology research. These findings support moving beyond pathway-level alteration status toward node-level, treatment-contextual dependency mapping to better identify actionable substructures within PDAC signaling networks and to accelerate hypothesis generation for rational therapeutic stratification.

## Data Availability

All data used in the present study is publicly available at https://www.cbioportal.org/ and https://genie.cbioportal.org. The datasets used in our study were aggregated/summary data, and no individual-level data were used. Additional data can be provided upon reasonable request to the authors.

## Supplementary Materials

**Table S1.**
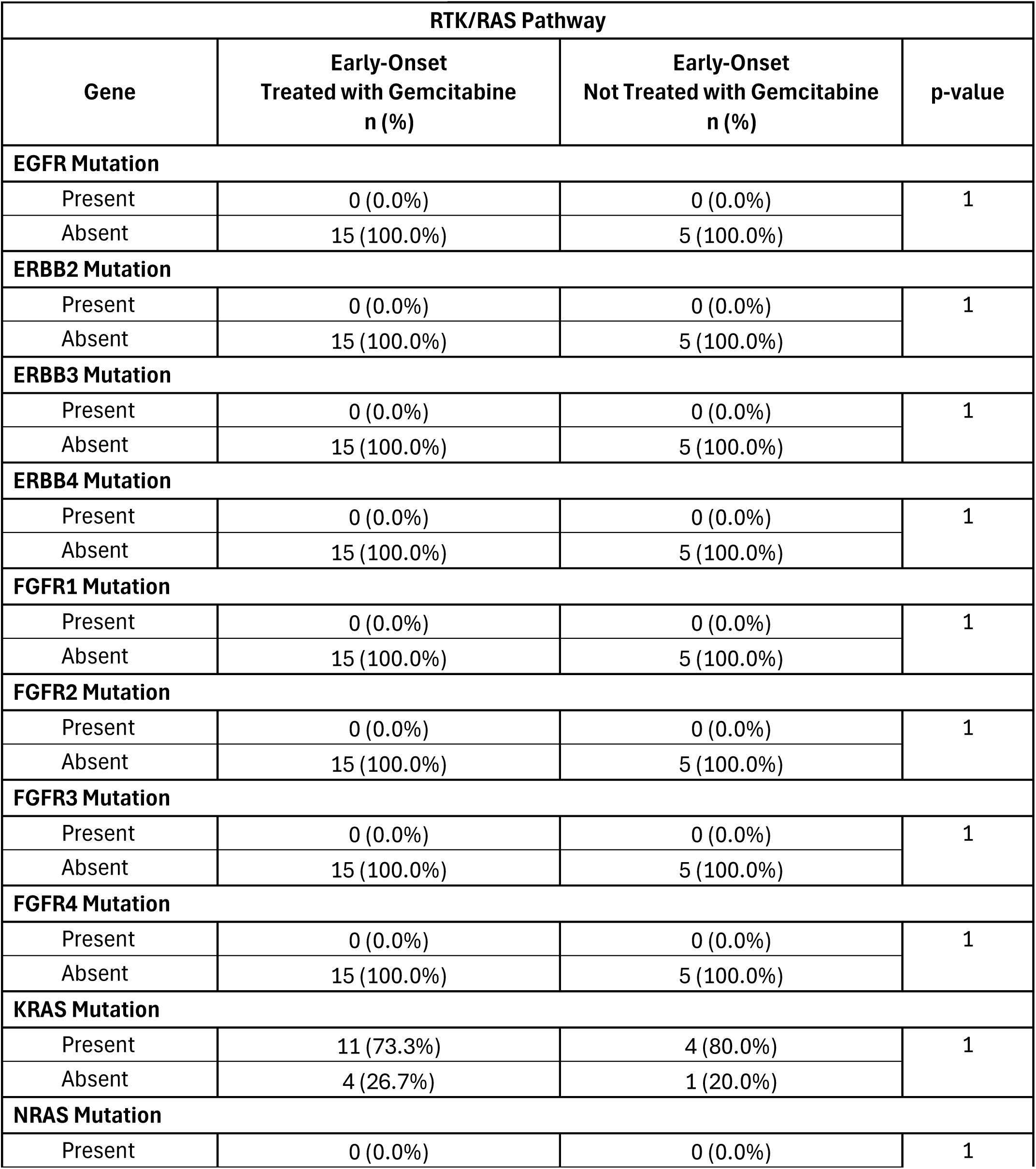
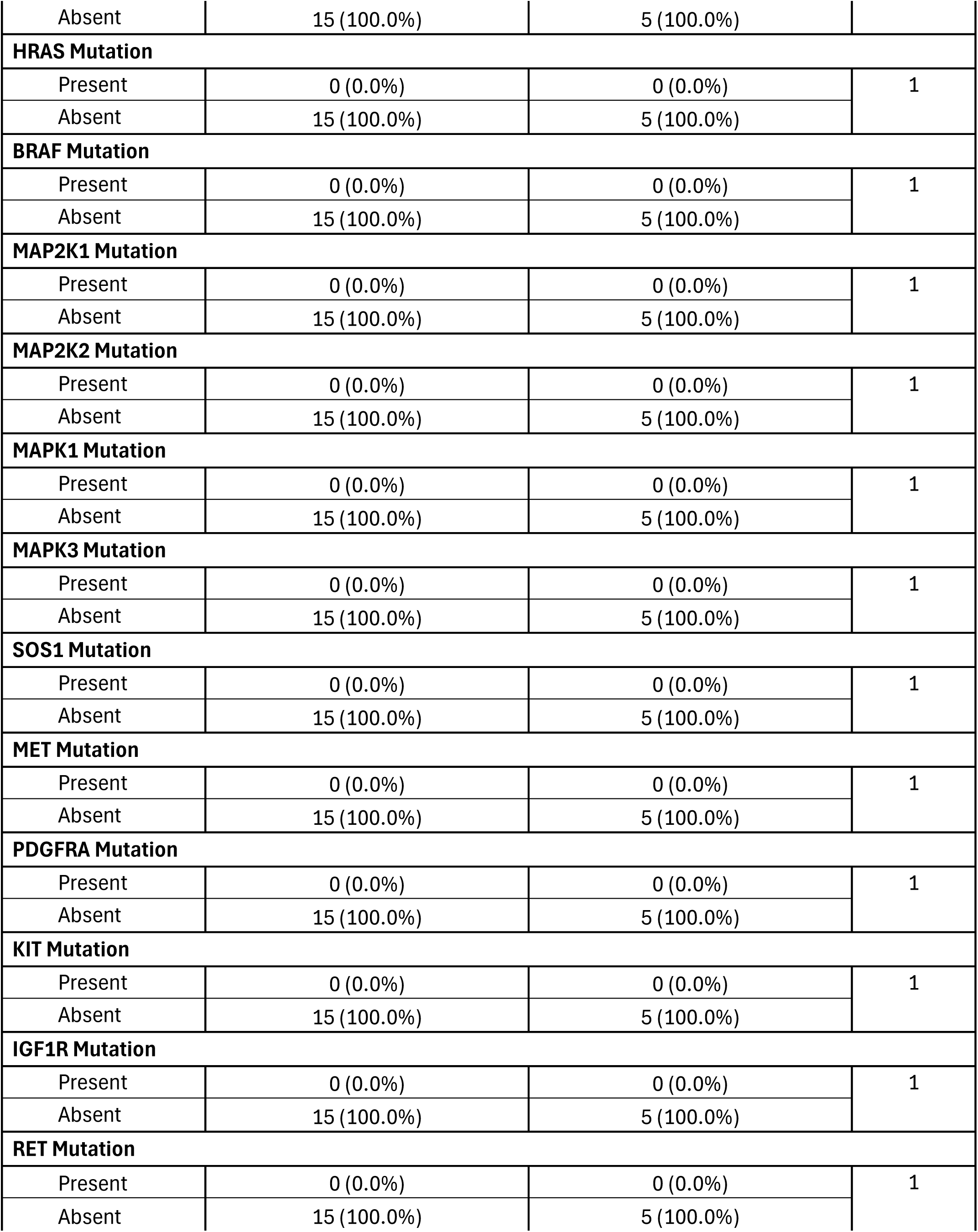
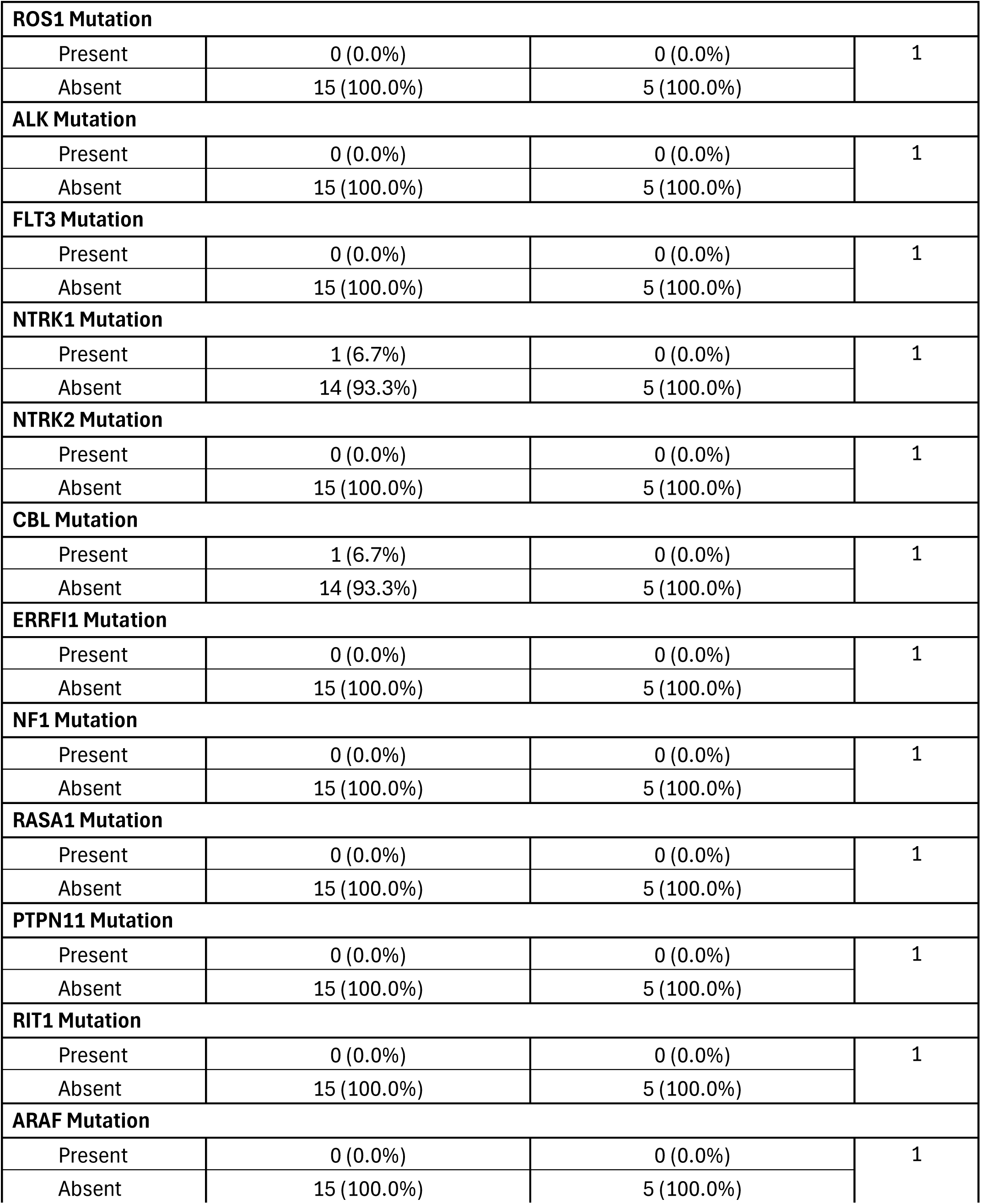
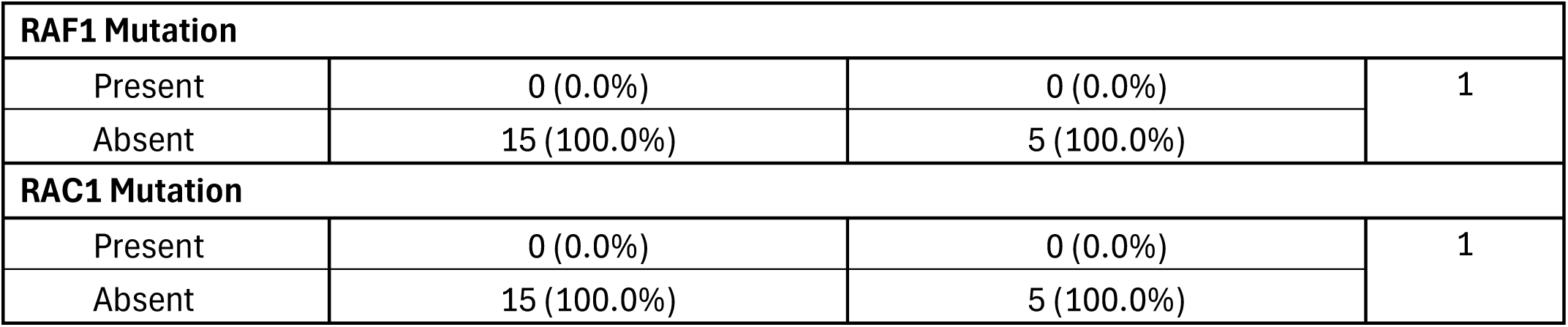
Comparison of Early-Onset PDAC Patients Treated with Gemcitabine Versus Those Not Treated with Gemcitabine.

**Table S2.** Comparison of Late-Onset PDAC Patients Treated with Gemcitabine Versus Those Not Treated with Gemcitabine.

| RTK/RAS Pathway |  |  |  |
| --- | --- | --- | --- |
| Gene | Late-Onset<br>Treated with Gemcitabine<br>n (%) | Late-Onset<br>Not Treated with Gemcitabine<br>n (%) | p-value |
| EGFR Mutation |  |  |  |
| Present | 2 (2.2%) | 0 (0.0%) | 0.503 |
| Absent | 89 (97.8%) | 73 (100.0%) |  |
| ERBB2 Mutation |  |  |  |
| Present | 6 (6.6%) | 0 (0.0%) | 0.034 |
| Absent | 85 (93.4%) | 73 (100.0%) |  |
| ERBB3 Mutation |  |  |  |
| Present | 2 (2.2%) | 0 (0.0%) | 0.503 |
| Absent | 89 (97.8%) | 73 (100.0%) |  |
| ERBB4 Mutation |  |  |  |
| Present | 5 (5.5%) | 1 (1.4%) | 0.2273 |
| Absent | 86 (94.5%) | 72 (98.6%) |  |
| FGFR1 Mutation |  |  |  |
| Present | 0 (0.0%) | 0 (0.0%) | 1 |
| Absent | 91 (100.0%) | 73 (100.0%) |  |
| FGFR2 Mutation |  |  |  |
| Present | 0 (0.0%) | 0 (0.0%) | 1 |
| Absent | 91 (100.0%) | 73 (100.0%) |  |
| FGFR3 Mutation |  |  |  |
| Present | 2 (2.2%) | 0 (0.0%) | 0.503 |
| Absent | 89 (97.8%) | 73 (100.0%) |  |
| FGFR4 Mutation |  |  |  |
| Present | 1 (1.1%) | 0 (0.0%) | 1 |
| Absent | 90 (98.9%) | 73 (100.0%) |  |
| KRAS Mutation |  |  |  |
| Present | 60 (65.9%) | 46 (63.0%) | 0.8224 |
| Absent | 31 (34.1%) | 27 (37.0%) |  |
| NRAS Mutation |  |  |  |
| Present | 0 (0.0%) | 0 (0.0%) | 1 |
| Absent | 91 (100.0%) | 73 (100.0%) |  |
| HRAS Mutation |  |  |  |
| Present | 1 (1.1%) | 0 (0.0%) | 1 |
| Absent | 90 (98.9%) | 73 (100.0%) |  |
| BRAF Mutation |  |  |  |
| Present | 0 (0.0%) | 2 (2.7%) | 0.1966 |
| Absent | 91 (100.0%) | 71 (97.3%) |  |
| MAP2K1 Mutation |  |  |  |
| Present | 0 (0.0%) | 0 (0.0%) | 1 |
| Absent | 91 (100.0%) | 73 (100.0%) |  |
| MAP2K2 Mutation |  |  |  |
| Present | 0 (0.0%) | 0 (0.0%) | 1 |
| Absent | 91 (100.0%) | 73 (100.0%) |  |
| MAPK1 Mutation |  |  |  |
| Present | 0 (0.0%) | 0 (0.0%) | 1 |
| Absent | 91 (100.0%) | 73 (100.0%) |  |
| MAPK3 Mutation |  |  |  |
| Present | 0 (0.0%) | 0 (0.0%) | 1 |
| Absent | 91 (100.0%) | 73 (100.0%) |  |
| SOS1 Mutation |  |  |  |
| Present | 2 (2.2%) | 0 (0.0%) | 0.503 |
| Absent | 89 (97.8%) | 73 (100.0%) |  |
| MET Mutation |  |  |  |
| Present | 4 (4.4%) | 0 (0.0%) | 0.1295 |
| Absent | 87 (95.6%) | 73 (100.0%) |  |
| PDGFRA Mutation |  |  |  |
| Present | 2 (2.2%) | 0 (0.0%) | 0.503 |
| Absent | 89 (97.8%) | 73 (100.0%) |  |
| KIT Mutation |  |  |  |
| Present | 3 (3.3%) | 0 (0.0%) | 0.2545 |
| Absent | 88 (96.7%) | 73 (100.0%) |  |
| IGF1R Mutation |  |  |  |
| Present | 3 (3.3%) | 0 (0.0%) | 0.2545 |
| Absent | 88 (96.7%) | 73 (100.0%) |  |
| RET Mutation |  |  |  |
| Present | 7 (7.7%) | 0 (0.0%) | 0.0175 |
| Absent | 84 (92.3%) | 73 (100.0%) |  |
| ROS1 Mutation |  |  |  |
| Present | 4 (4.4%) | 0 (0.0%) | 0.1295 |
| Absent | 87 (95.6%) | 73 (100.0%) |  |
| ALK Mutation |  |  |  |
| Present | 2 (2.2%) | 0 (0.0%) | 0.503 |
| Absent | 89 (97.8%) | 73 (100.0%) |  |
| FLT3 Mutation |  |  |  |
| Present | 2 (2.2%) | 1 (1.4%) | 1 |
| Absent | 89 (97.8%) | 72 (98.6%) |  |
| NTRK1 Mutation |  |  |  |
| Present | 1 (1.1%) | 1 (1.4%) | 1 |
| Absent | 90 (98.9%) | 72 (98.6%) |  |
| NTRK2 Mutation |  |  |  |
| Present | 2 (2.2%) | 0 (0.0%) | 0.503 |
| Absent | 89 (97.8%) | 73 (100.0%) |  |
| CBL Mutation |  |  |  |
| Present | 0 (0.0%) | 0 (0.0%) | 1 |
| Absent | 91 (100.0%) | 73 (100.0%) |  |
| ERRFI1 Mutation |  |  |  |
| Present | 0 (0.0%) | 0 (0.0%) | 1 |
| Absent | 91 (100.0%) | 73 (100.0%) |  |
| NF1 Mutation |  |  |  |
| Present | 2 (2.2%) | 0 (0.0%) | 0.503 |
| Absent | 89 (97.8%) | 73 (100.0%) |  |
| RASA1 Mutation |  |  |  |
| Present | 1 (1.1%) | 1 (1.4%) | 1 |
| Absent | 90 (98.9%) | 72 (98.6%) |  |
| PTPN11 Mutation |  |  |  |
| Present | 0 (0.0%) | 0 (0.0%) | 1 |
| Absent | 91 (100.0%) | 73 (100.0%) |  |
| RIT1 Mutation |  |  |  |
| Present | 0 (0.0%) | 0 (0.0%) | 1 |
| Absent | 91 (100.0%) | 73 (100.0%) |  |
| ARAF Mutation |  |  |  |
| Present | 3 (3.3%) | 0 (0.0%) | 0.2545 |
| Absent | 88 (96.7%) | 73 (100.0%) |  |
| RAF1 Mutation |  |  |  |
| Present | 0 (0.0%) | 0 (0.0%) | 1 |
| Absent | 91 (100.0%) | 73 (100.0%) |  |
| RAC1 Mutation |  |  |  |
| Present | 0 (0.0%) | 0 (0.0%) | 1 |
| Absent | 91 (100.0%) | 73 (100.0%) |  |

**Table S3.**
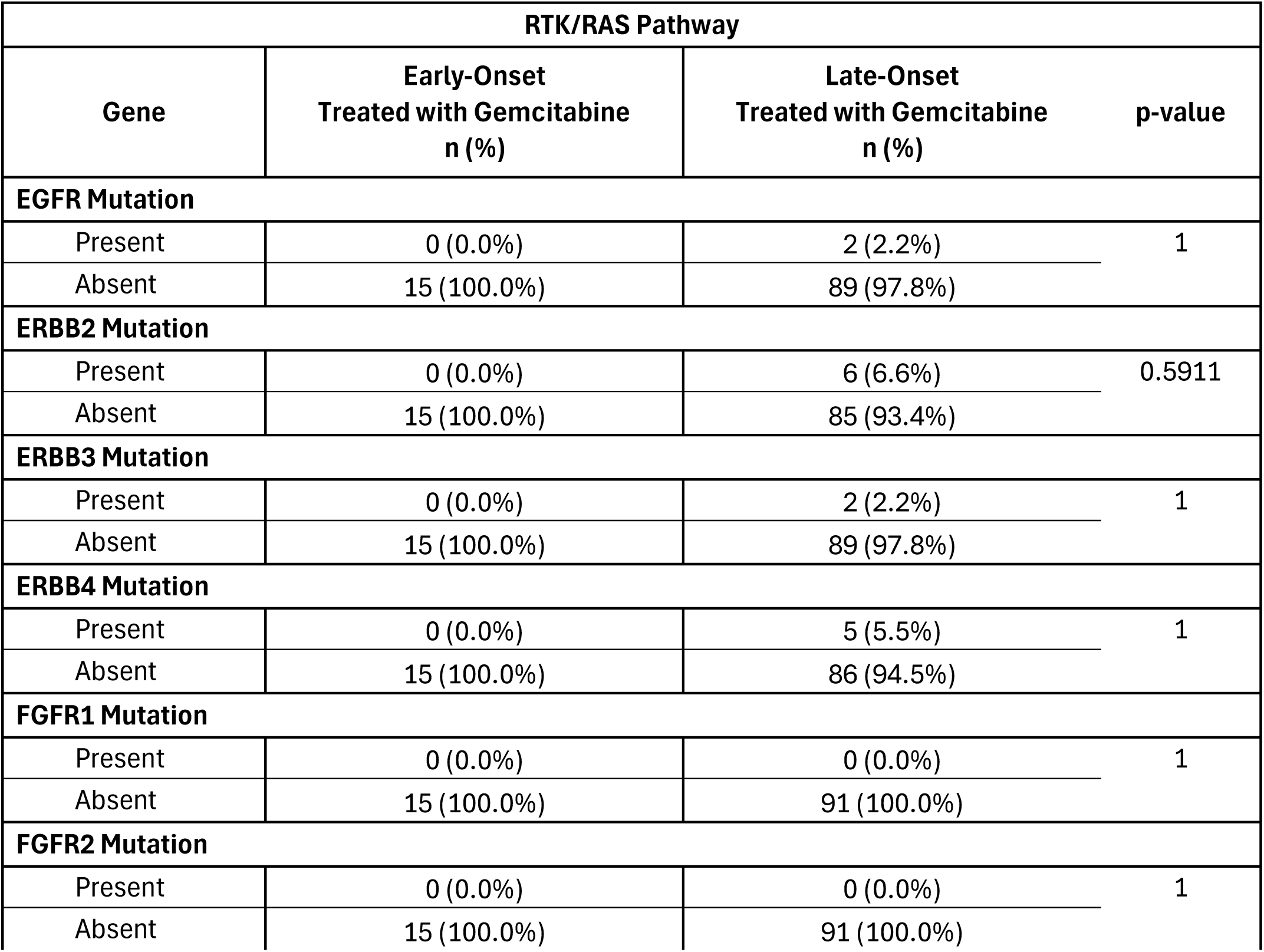
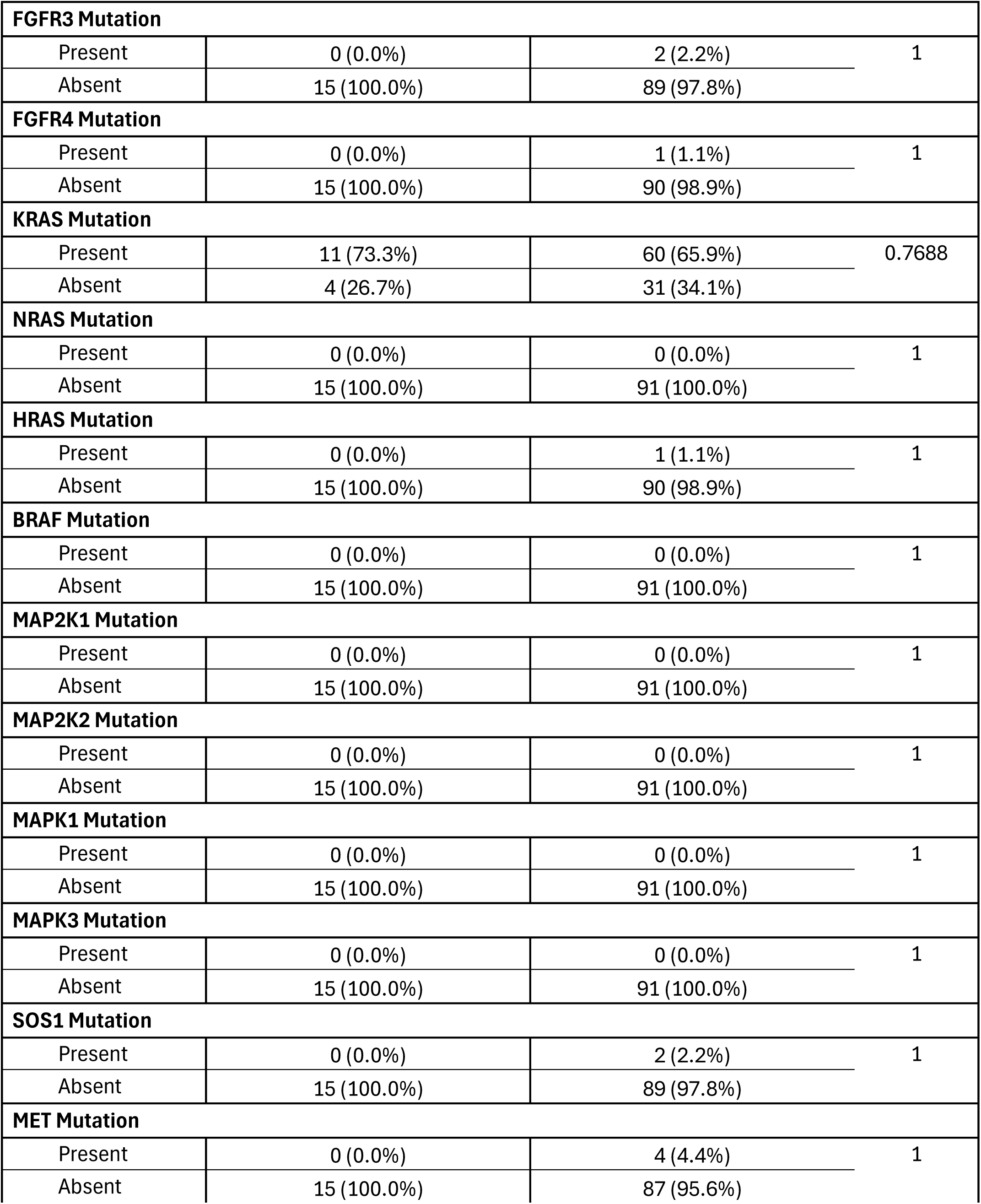
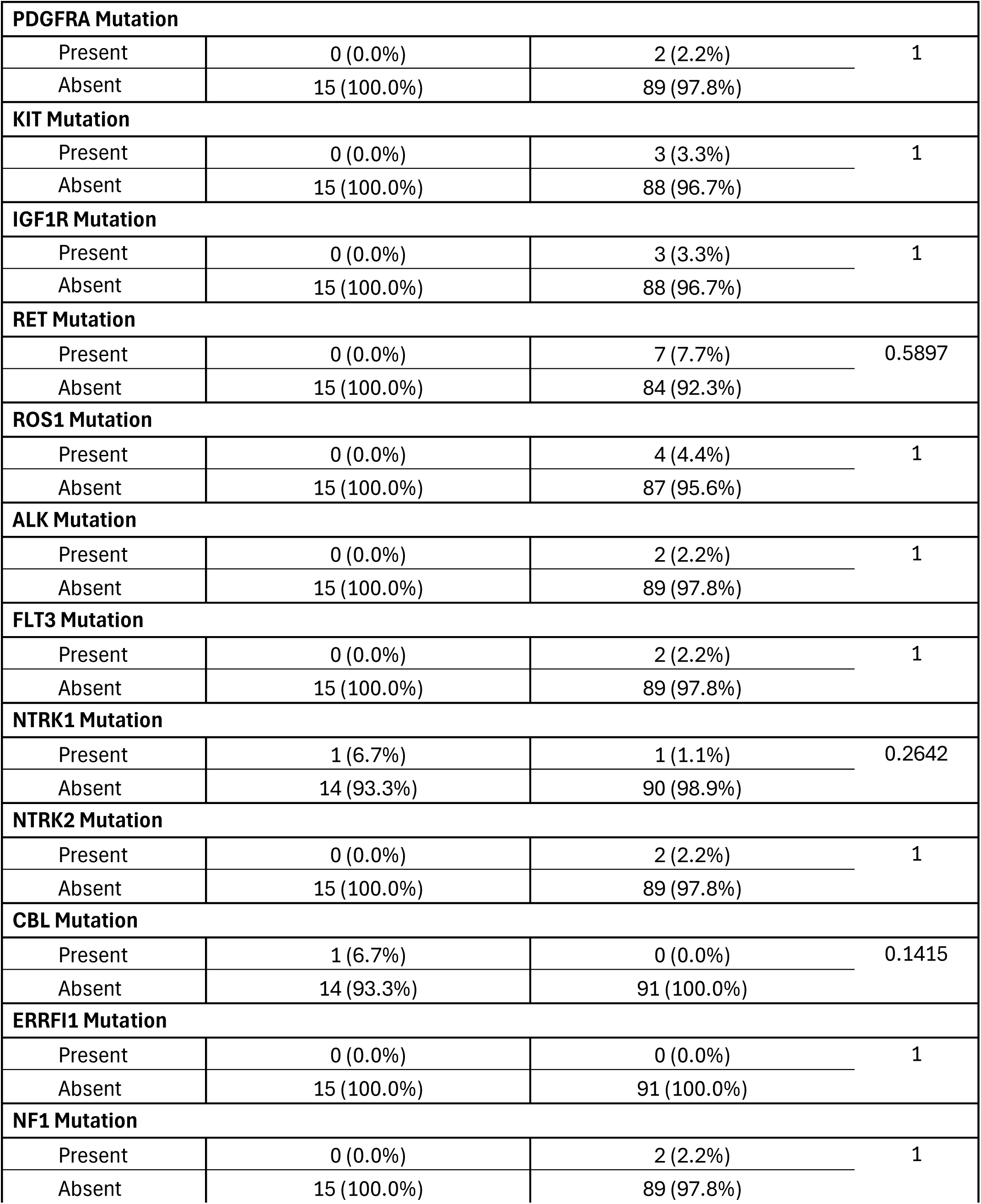
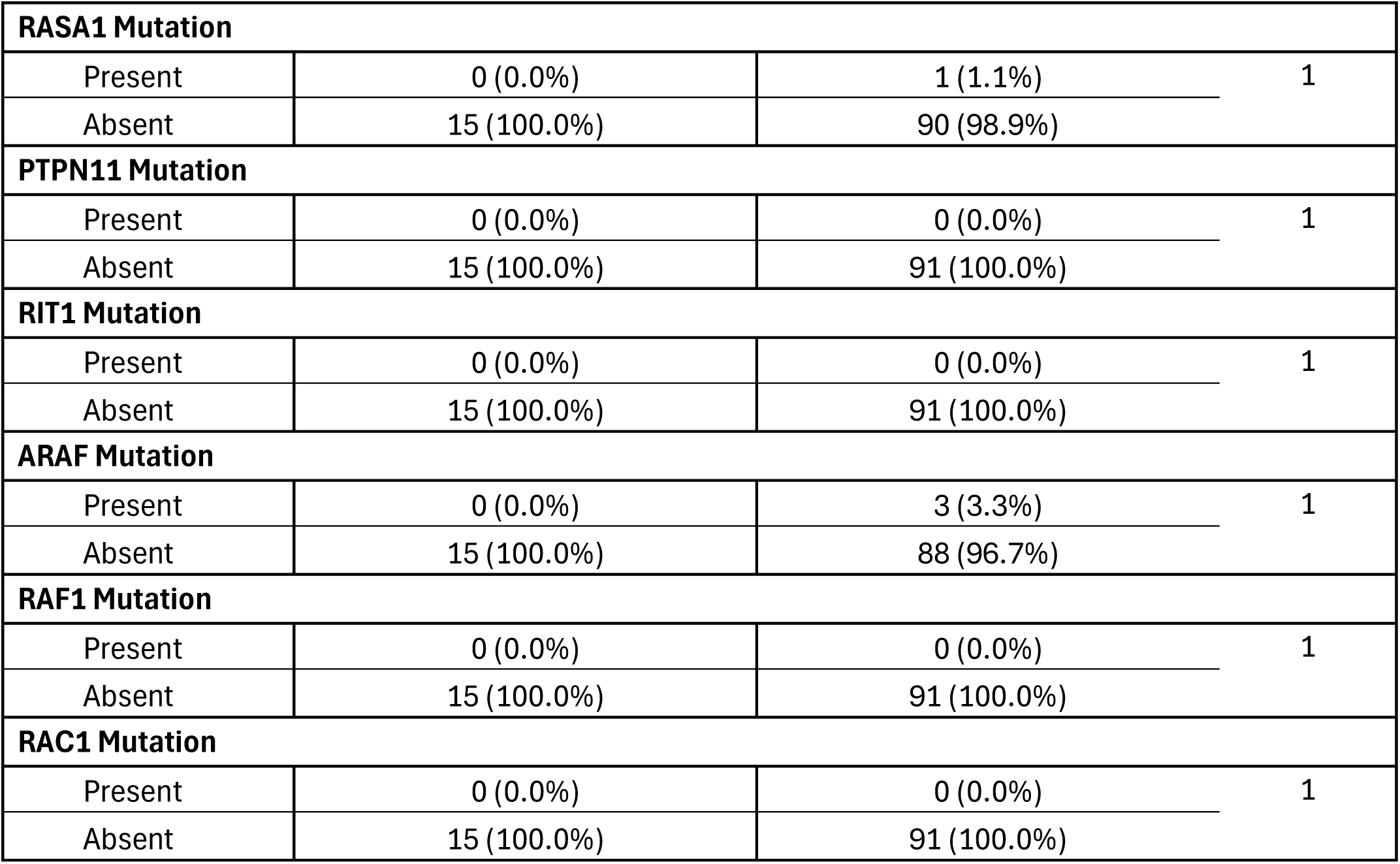
Comparison of Early-Onset PDAC Patients Versus Late-Onset PDAC Patients Treated with Gemcitabine.

**Table S4.** Comparison of Early-Onset PDAC Patients Versus Late-Onset PDAC Patients Not Treated with Gemcitabine.

| RTK/RAS Pathway |  |  |  |
| --- | --- | --- | --- |
| Gene | Early-Onset<br>Not Treated with Gemcitabine<br>n (%) | Late-Onset<br>Not Treated with Gemcitabine<br>n (%) | p-value |
| EGFR Mutation |  |  |  |
| Present | 0 (0.0%) | 0 (0.0%) | 1 |
| Absent | 5 (100.0%) | 73 (100.0%) |  |
| ERBB2 Mutation |  |  |  |
| Present | 0 (0.0%) | 0 (0.0%) | 1 |
| Absent | 5 (100.0%) | 73 (100.0%) |  |
| ERBB3 Mutation |  |  |  |
| Present | 0 (0.0%) | 0 (0.0%) | 1 |
| Absent | 5 (100.0%) | 73 (100.0%) |  |
| ERBB4 Mutation |  |  |  |
| Present | 0 (0.0%) | 1 (1.4%) | 1 |
| Absent | 5 (100.0%) | 72 (98.6%) |  |
| FGFR1 Mutation |  |  |  |
| Present | 0 (0.0%) | 0 (0.0%) | 1 |
| Absent | 5 (100.0%) | 73 (100.0%) |  |
| FGFR2 Mutation |  |  |  |
| Present | 0 (0.0%) | 0 (0.0%) | 1 |
| Absent | 5 (100.0%) | 73 (100.0%) |  |
| FGFR3 Mutation |  |  |  |
| Present | 0 (0.0%) | 0 (0.0%) | 1 |
| Absent | 5 (100.0%) | 73 (100.0%) |  |
| FGFR4 Mutation |  |  |  |
| Present | 0 (0.0%) | 0 (0.0%) | 1 |
| Absent | 5 (100.0%) | 73 (100.0%) |  |
| KRAS Mutation |  |  |  |
| Present | 4 (80.0%) | 46 (63.0%) | 0.6491 |
| Absent | 1 (20.0%) | 27 (37.0%) |  |
| NRAS Mutation |  |  |  |
| Present | 0 (0.0%) | 0 (0.0%) | 1 |
| Absent | 5 (100.0%) | 73 (100.0%) |  |
| HRAS Mutation |  |  |  |
| Present | 0 (0.0%) | 0 (0.0%) | 1 |
| Absent | 5 (100.0%) | 73 (100.0%) |  |
| BRAF Mutation |  |  |  |
| Present | 0 (0.0%) | 2 (2.7%) | 1 |
| Absent | 5 (100.0%) | 71 (97.3%) |  |
| MAP2K1 Mutation |  |  |  |
| Present | 0 (0.0%) | 0 (0.0%) | 1 |
| Absent | 5 (100.0%) | 73 (100.0%) |  |
| MAP2K2 Mutation |  |  |  |
| Present | 0 (0.0%) | 0 (0.0%) | 1 |
| Absent | 5 (100.0%) | 73 (100.0%) |  |
| MAPK1 Mutation |  |  |  |
| Present | 0 (0.0%) | 0 (0.0%) | 1 |
| Absent | 5 (100.0%) | 73 (100.0%) |  |
| MAPK3 Mutation |  |  |  |
| Present | 0 (0.0%) | 0 (0.0%) | 1 |
| Absent | 5 (100.0%) | 73 (100.0%) |  |
| SOS1 Mutation |  |  |  |
| Present | 0 (0.0%) | 0 (0.0%) | 1 |
| Absent | 5 (100.0%) | 73 (100.0%) |  |
| MET Mutation |  |  |  |
| Present | 0 (0.0%) | 0 (0.0%) | 1 |
| Absent | 5 (100.0%) | 73 (100.0%) |  |
| PDGFRA Mutation |  |  |  |
| Present | 0 (0.0%) | 0 (0.0%) | 1 |
| Absent | 5 (100.0%) | 73 (100.0%) |  |
| KIT Mutation |  |  |  |
| Present | 0 (0.0%) | 0 (0.0%) | 1 |
| Absent | 5 (100.0%) | 73 (100.0%) |  |
| IGF1R Mutation |  |  |  |
| Present | 0 (0.0%) | 0 (0.0%) | 1 |
| Absent | 5 (100.0%) | 73 (100.0%) |  |
| RET Mutation |  |  |  |
| Present | 0 (0.0%) | 0 (0.0%) | 1 |
| Absent | 5 (100.0%) | 73 (100.0%) |  |
| ROS1 Mutation |  |  |  |
| Present | 0 (0.0%) | 0 (0.0%) | 1 |
| Absent | 5 (100.0%) | 73 (100.0%) |  |
| ALK Mutation |  |  |  |
| Present | 0 (0.0%) | 0 (0.0%) | 1 |
| Absent | 5 (100.0%) | 73 (100.0%) |  |
| FLT3 Mutation |  |  |  |
| Present | 0 (0.0%) | 1 (1.4%) | 1 |
| Absent | 5 (100.0%) | 72 (98.6%) |  |
| NTRK1 Mutation |  |  |  |
| Present | 0 (0.0%) | 1 (1.4%) | 1 |
| Absent | 5 (100.0%) | 72 (98.6%) |  |
| NTRK2 Mutation |  |  |  |
| Present | 0 (0.0%) | 0 (0.0%) | 1 |
| Absent | 5 (100.0%) | 73 (100.0%) |  |
| CBL Mutation |  |  |  |
| Present | 0 (0.0%) | 0 (0.0%) | 1 |
| Absent | 5 (100.0%) | 73 (100.0%) |  |
| ERRFI1 Mutation |  |  |  |
| Present | 0 (0.0%) | 0 (0.0%) | 1 |
| Absent | 5 (100.0%) | 73 (100.0%) |  |
| NF1 Mutation |  |  |  |
| Present | 0 (0.0%) | 0 (0.0%) | 1 |
| Absent | 5 (100.0%) | 73 (100.0%) |  |
| RASA1 Mutation |  |  |  |
| Present | 0 (0.0%) | 1 (1.4%) | 1 |
| Absent | 5 (100.0%) | 72 (98.6%) |  |
| PTPN11 Mutation |  |  |  |
| Present | 0 (0.0%) | 0 (0.0%) | 1 |
| Absent | 5 (100.0%) | 73 (100.0%) |  |
| RIT1 Mutation |  |  |  |
| Present | 0 (0.0%) | 0 (0.0%) | 1 |
| Absent | 5 (100.0%) | 73 (100.0%) |  |
| ARAF Mutation |  |  |  |
| Present | 0 (0.0%) | 0 (0.0%) | 1 |
| Absent | 5 (100.0%) | 73 (100.0%) |  |
| RAF1 Mutation |  |  |  |
| Present | 0 (0.0%) | 0 (0.0%) | 1 |
| Absent | 5 (100.0%) | 73 (100.0%) |  |
| RAC1 Mutation |  |  |  |
| Present | 0 (0.0%) | 0 (0.0%) | 1 |
| Absent | 5 (100.0%) | 73 (100.0%) |  |

**Table S5.**
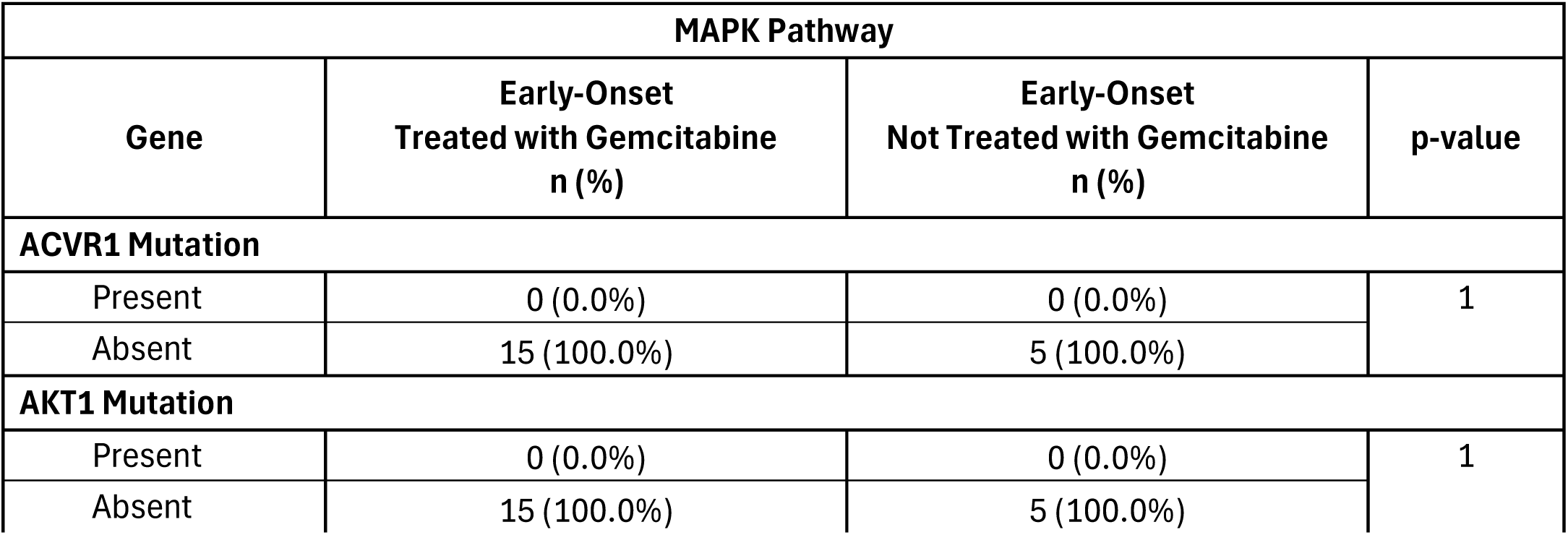
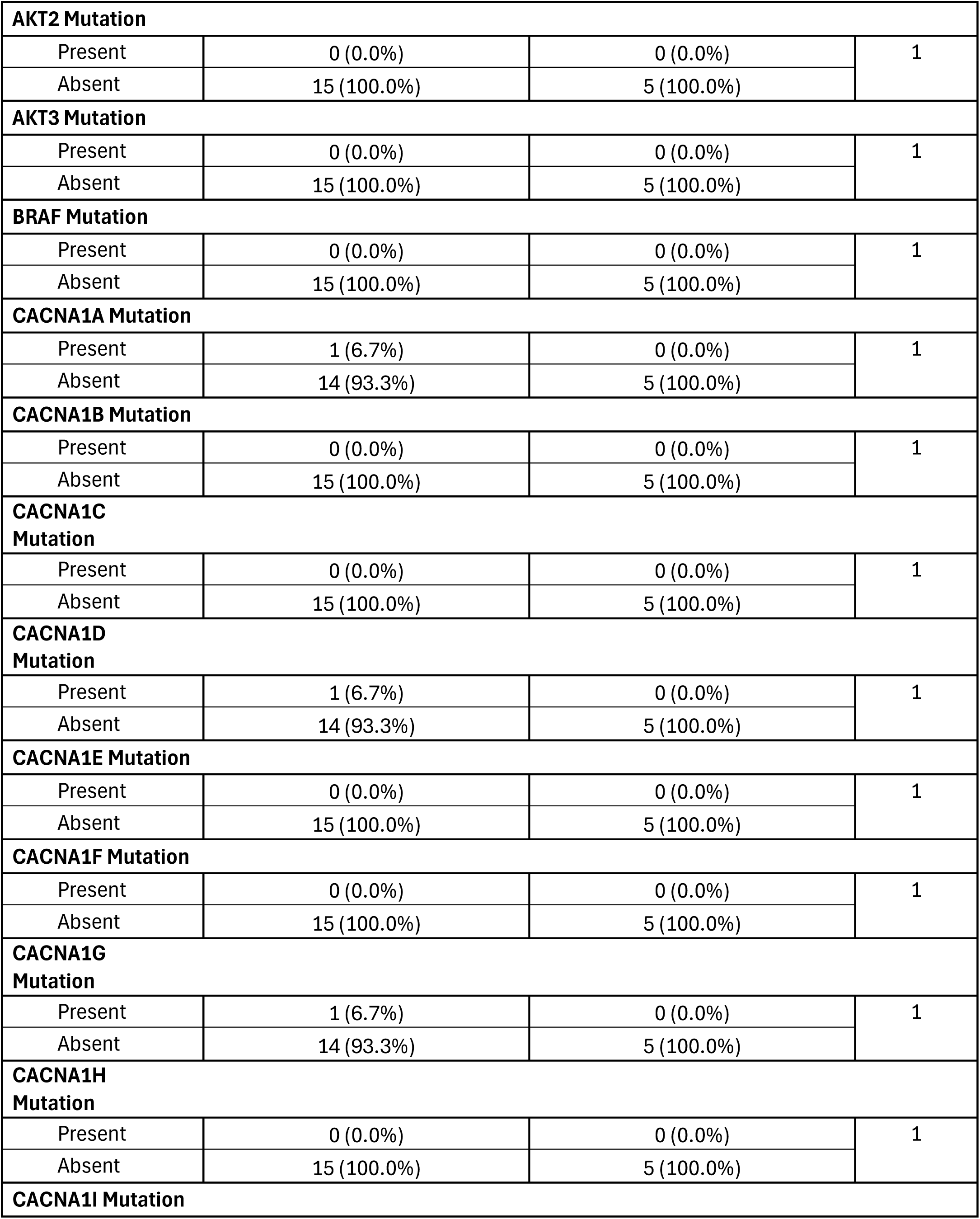
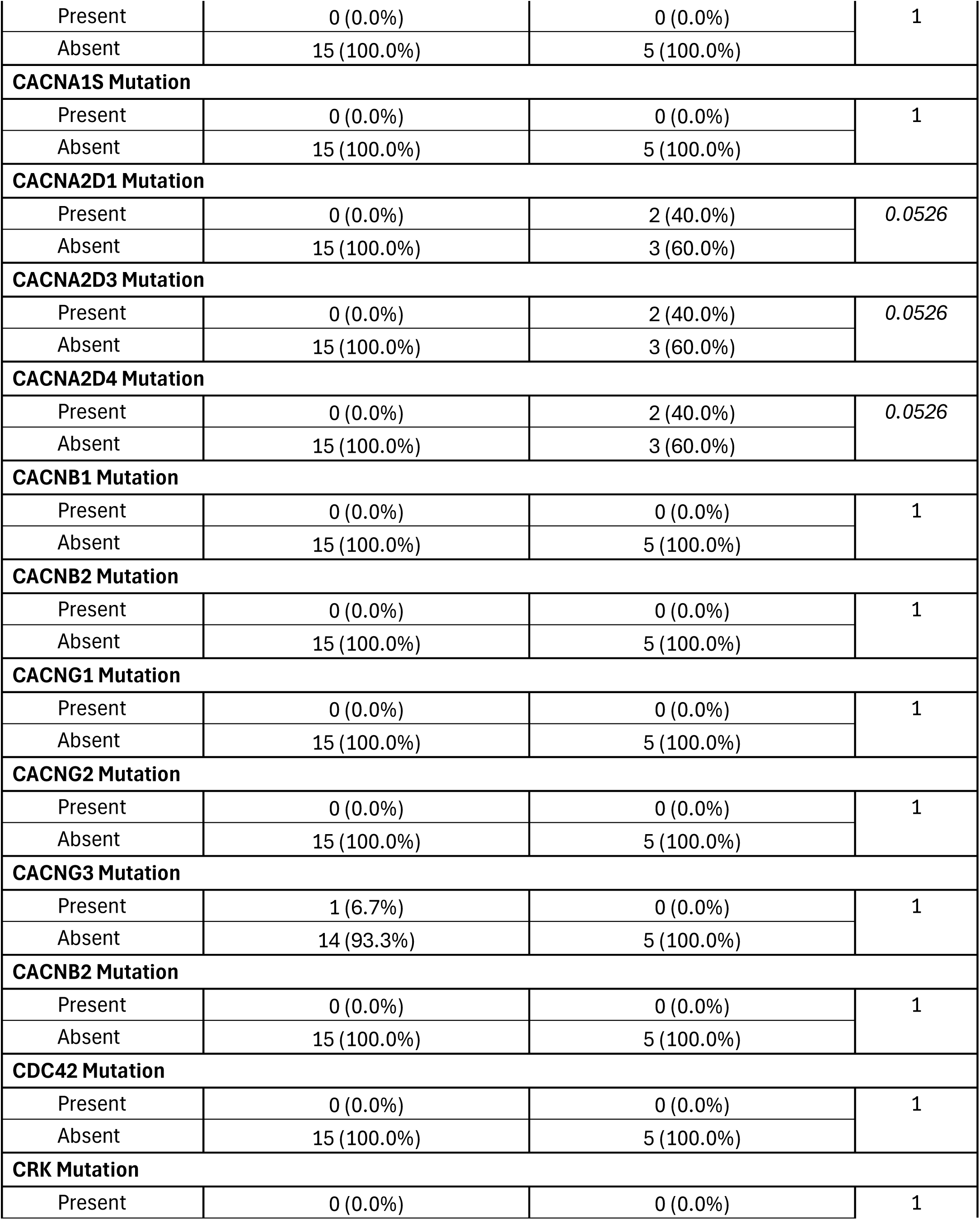
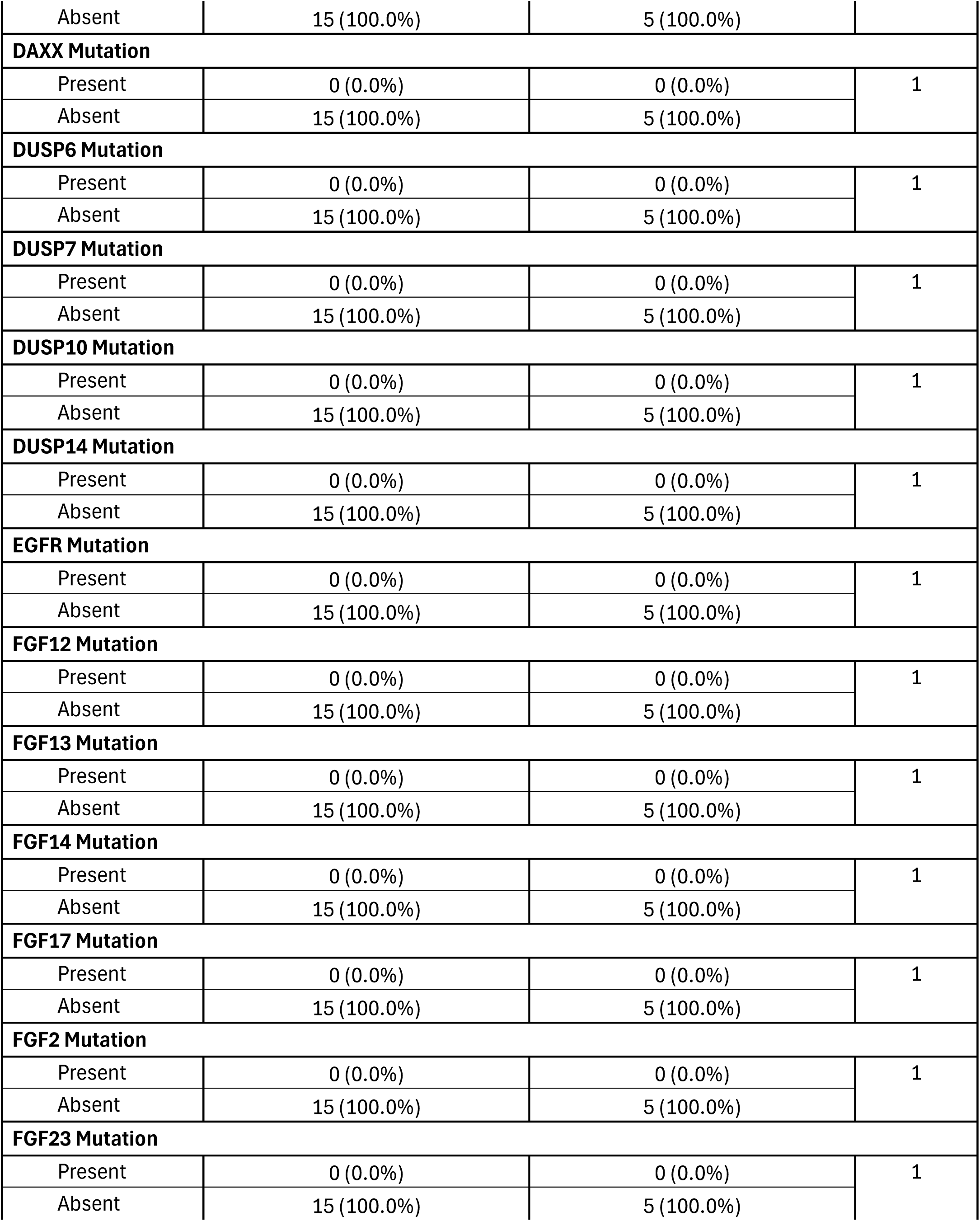
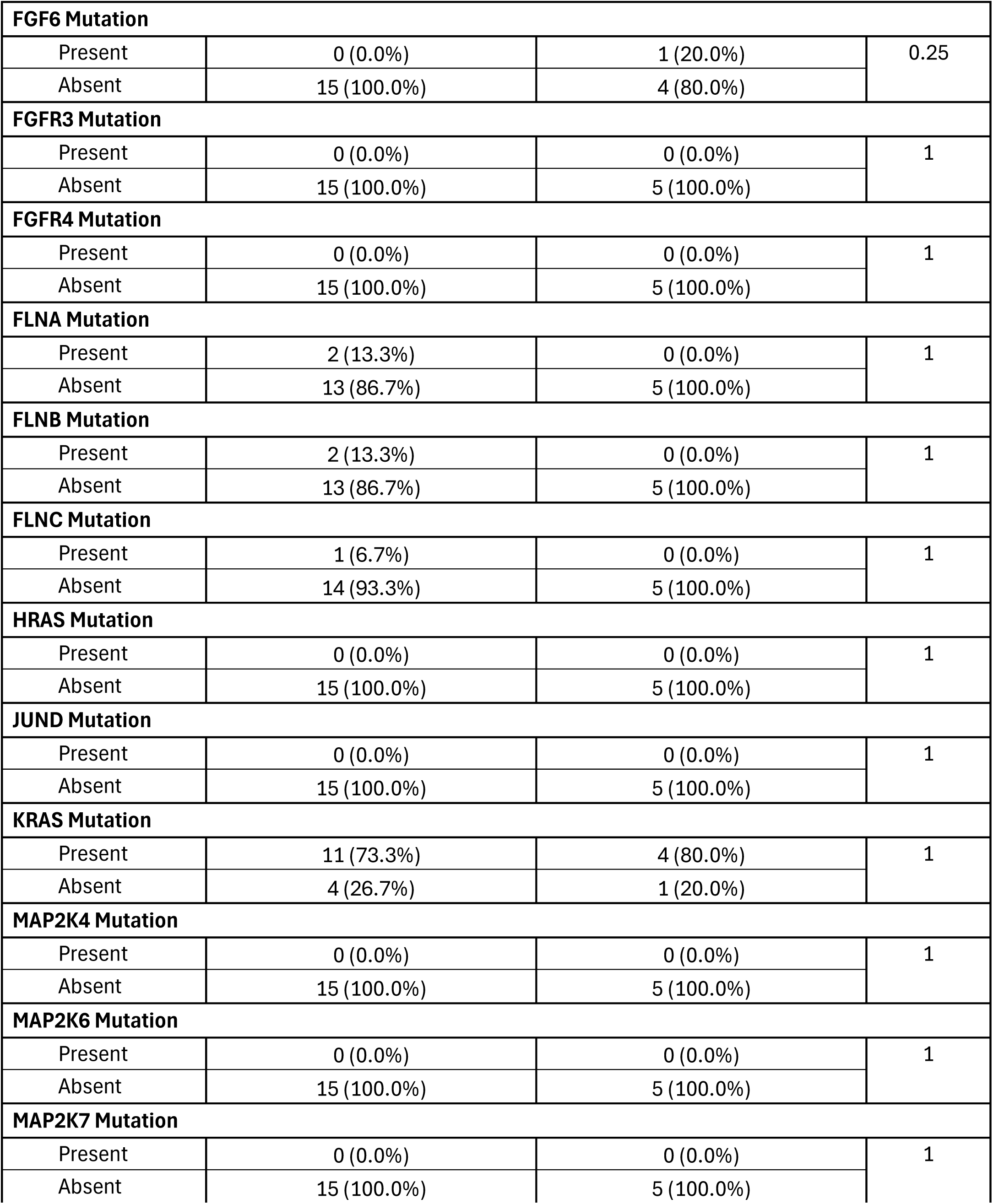
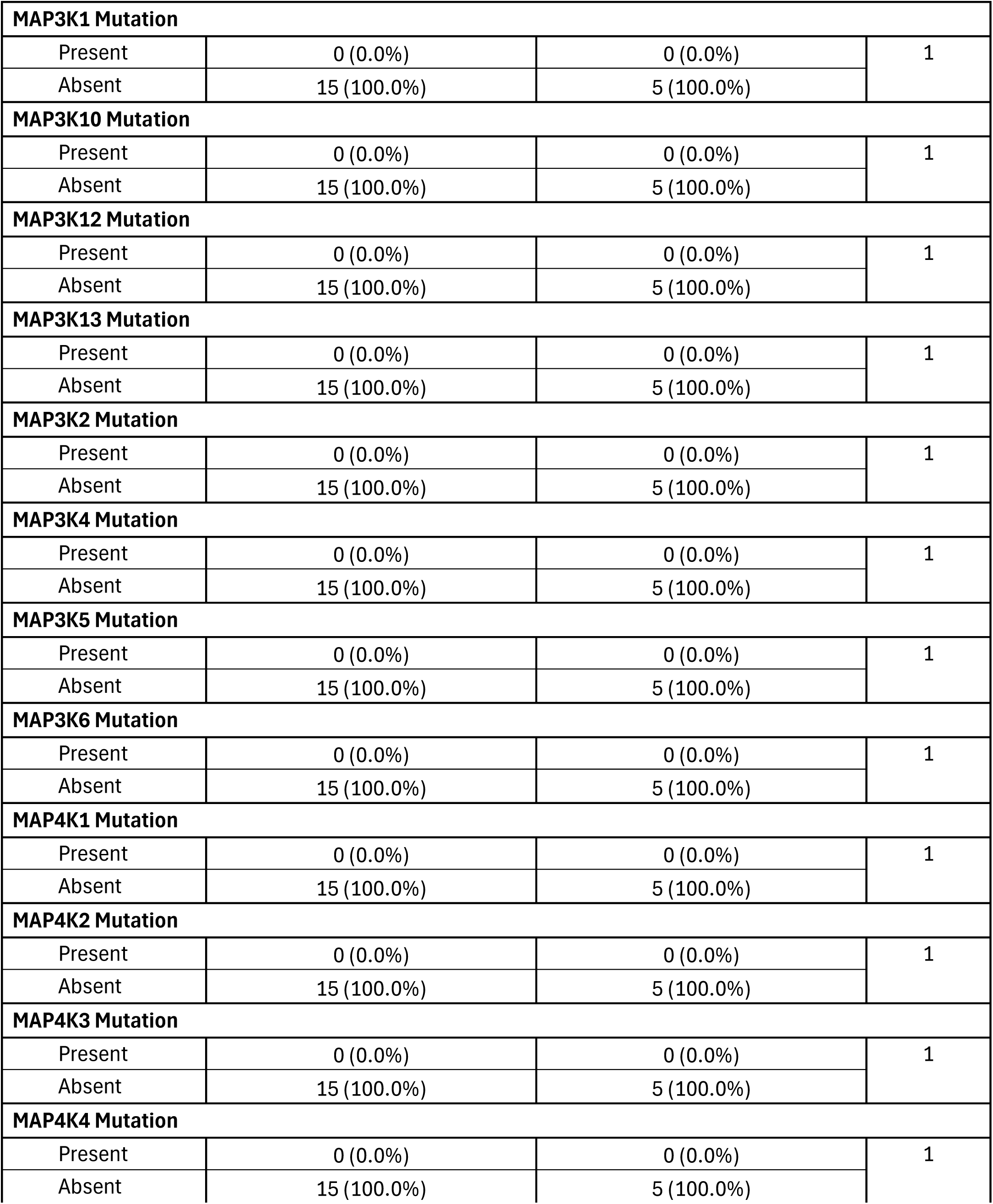
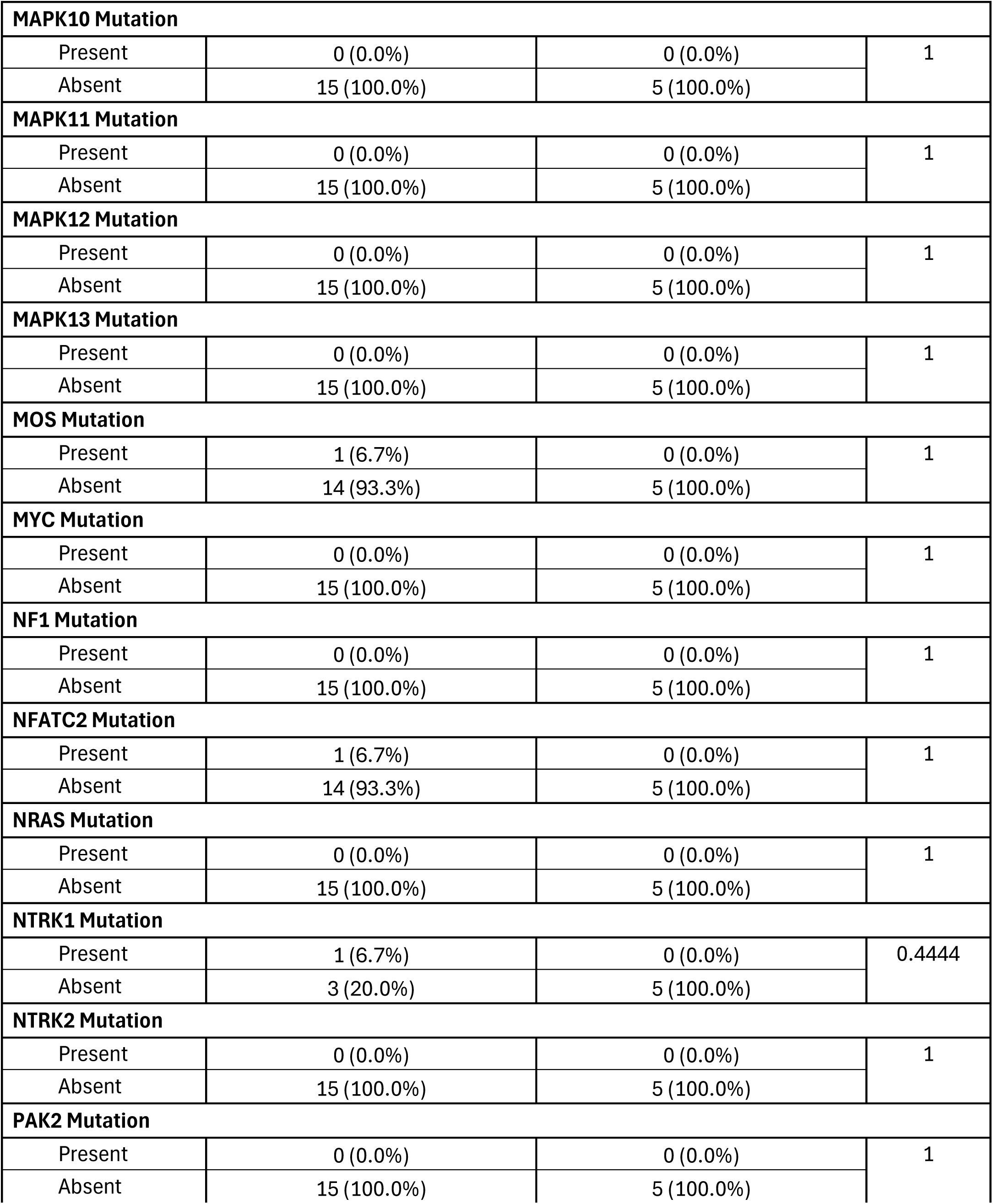
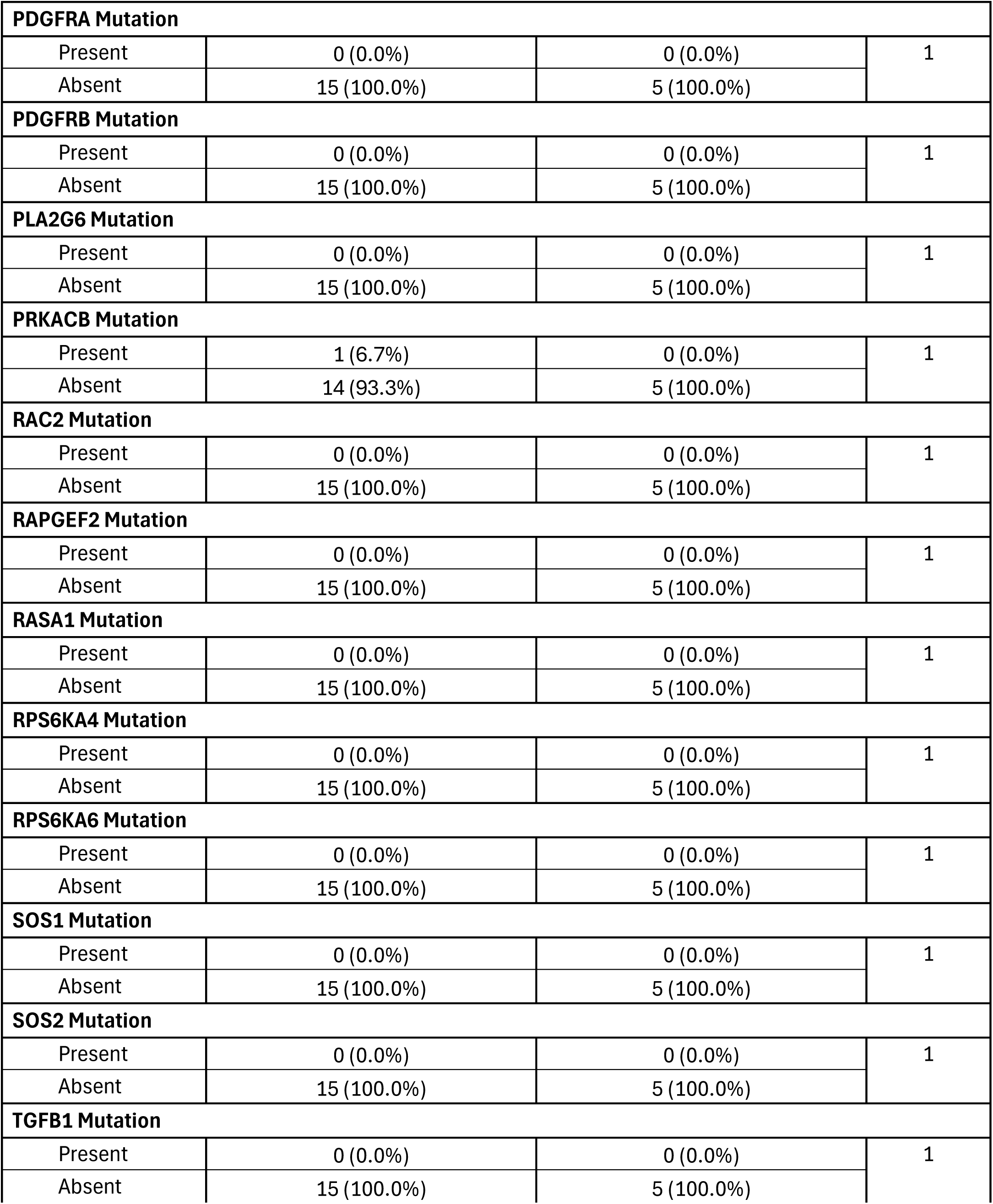
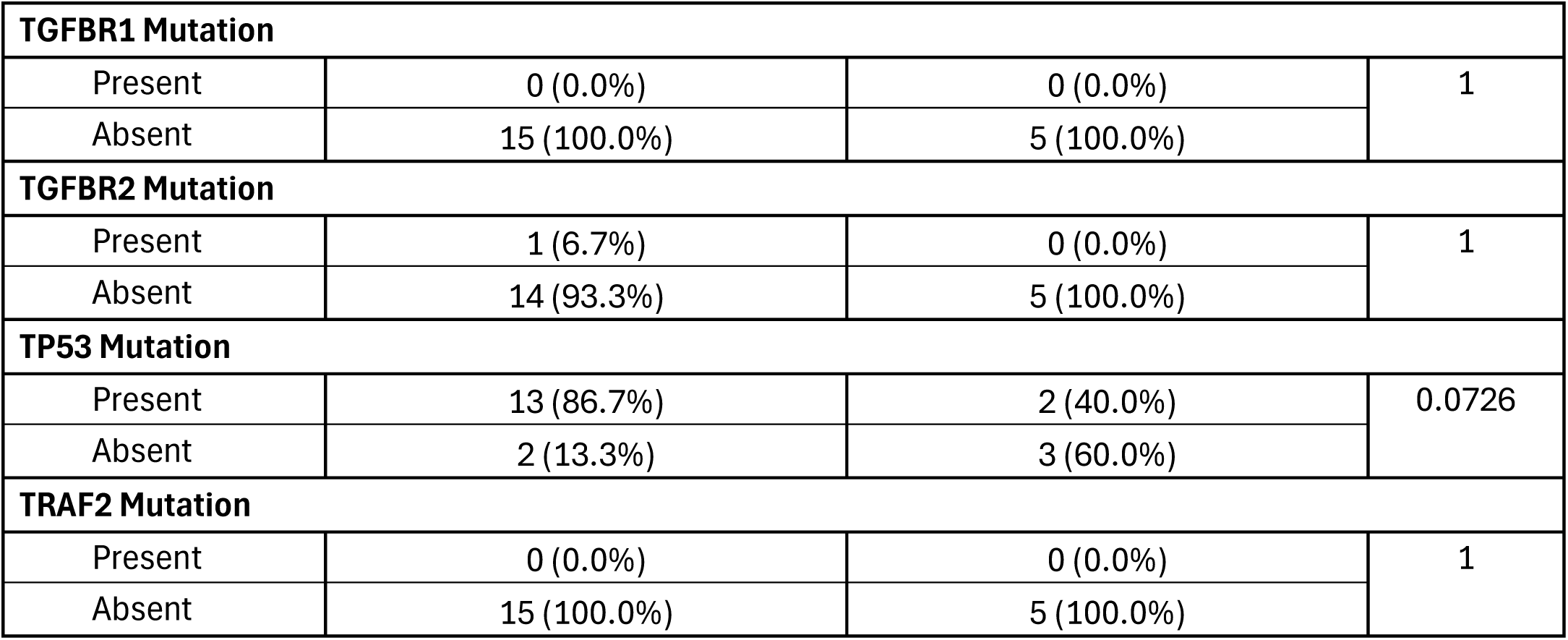
Comparison of Early-Onset PDAC Patients Treated with Gemcitabine Versus Those Not Treated with Gemcitabine.

**Table S6.**
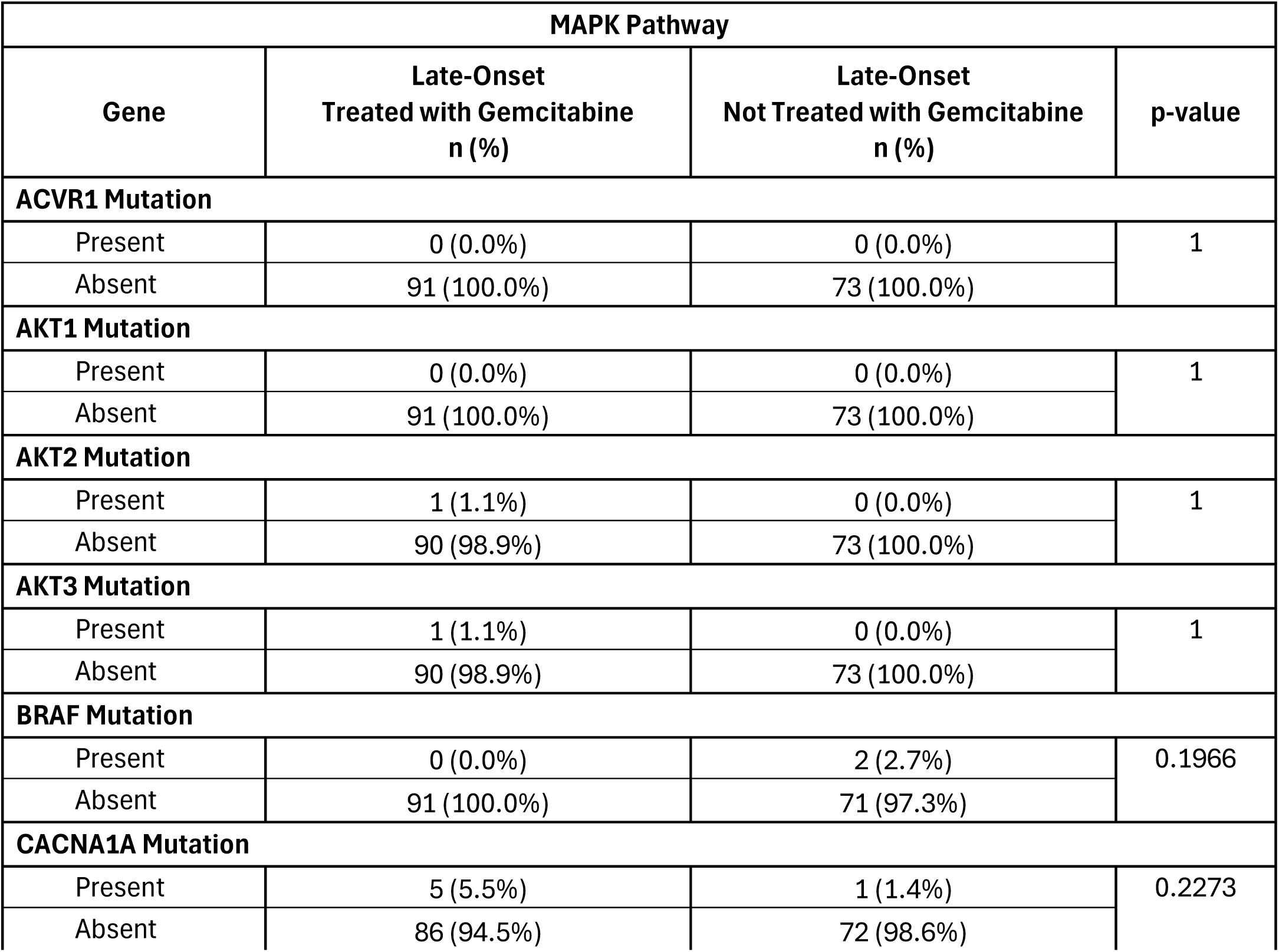
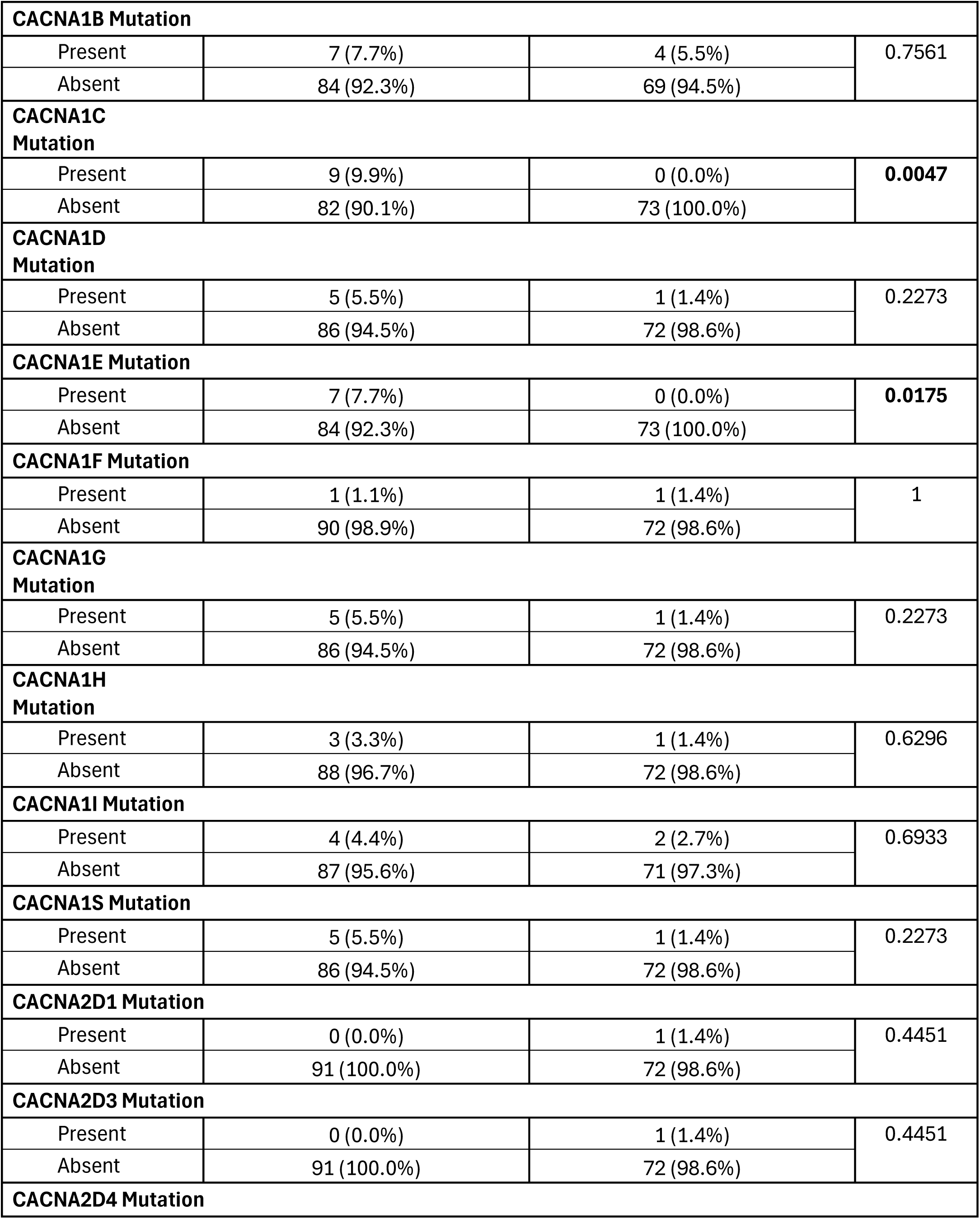
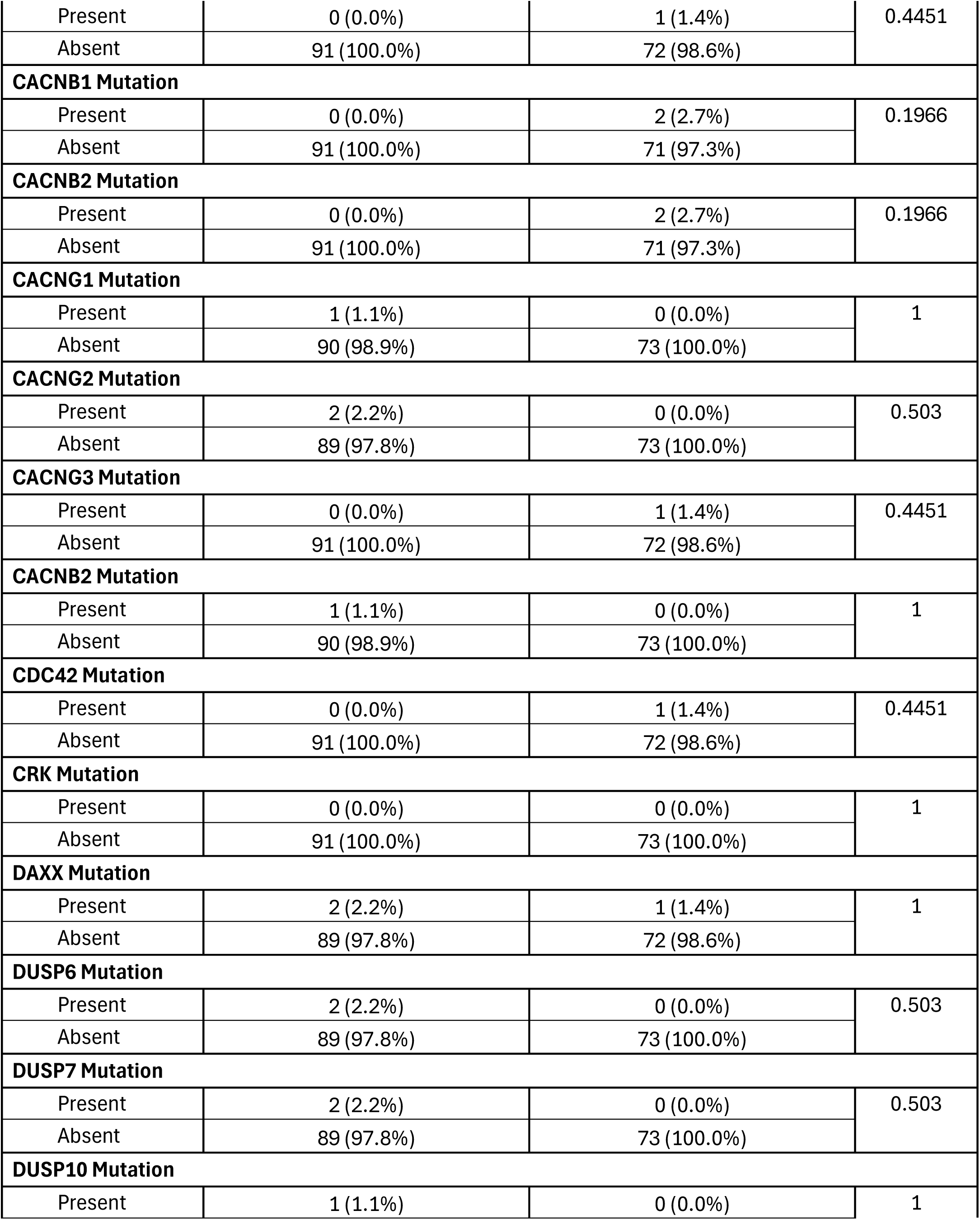
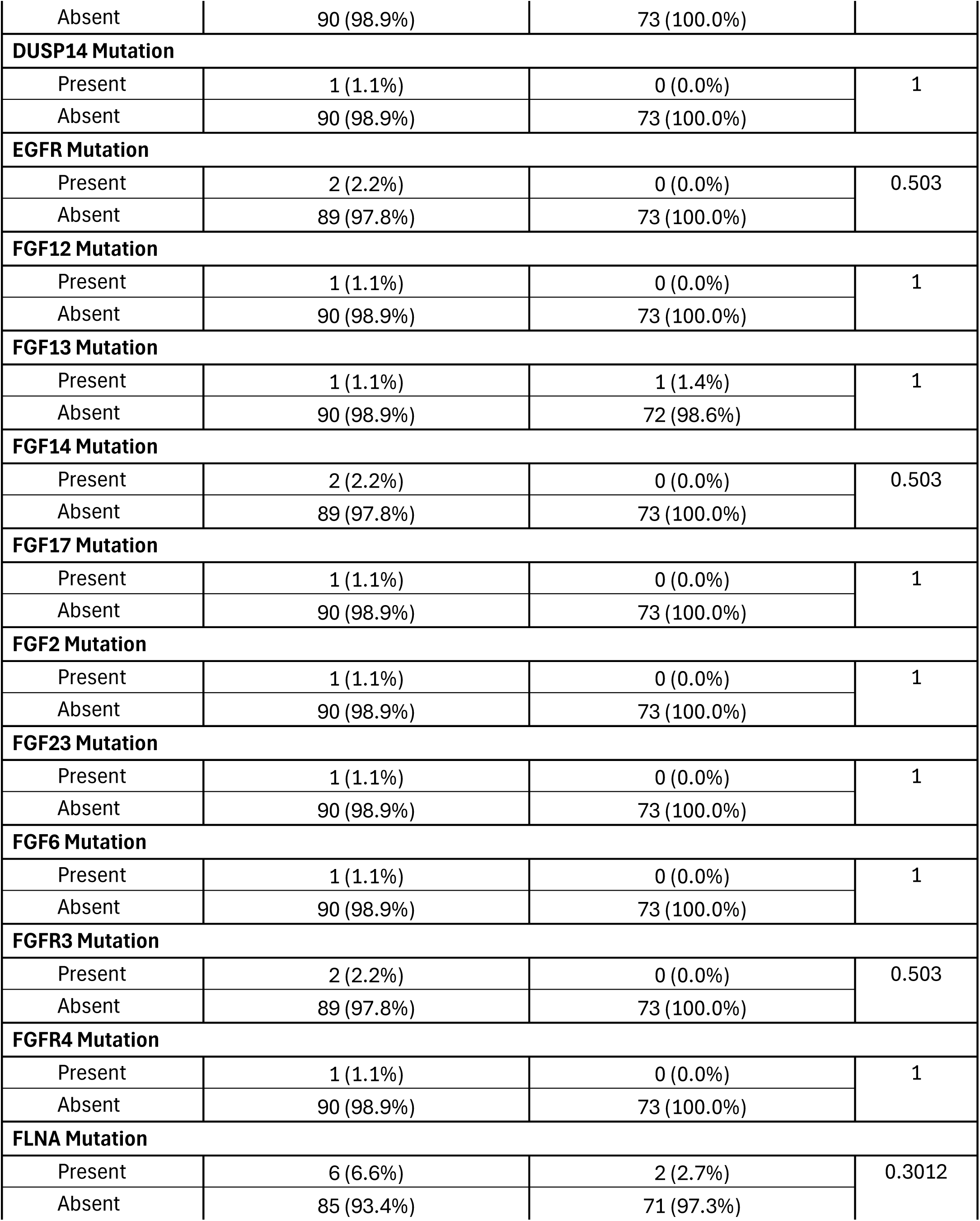
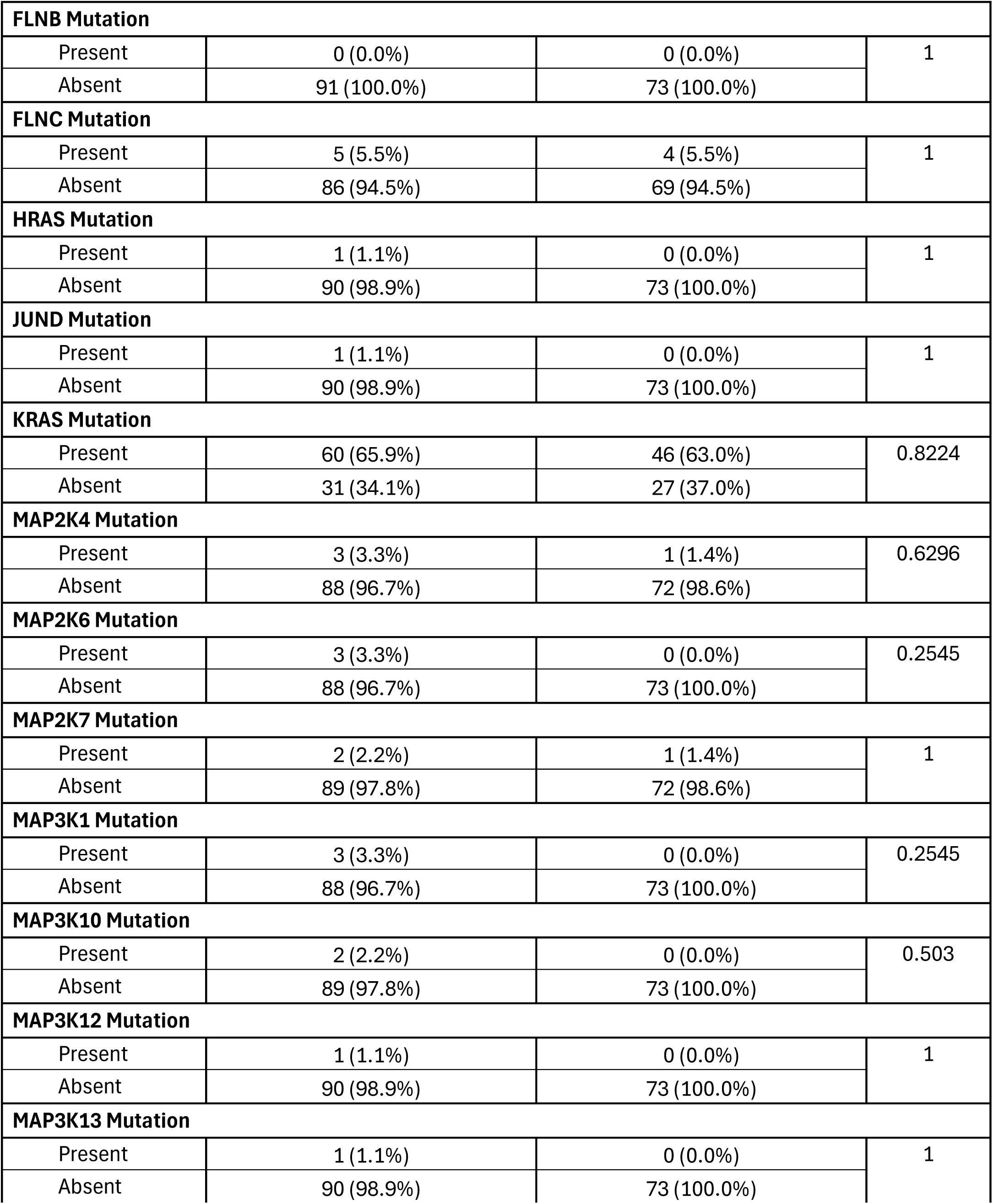
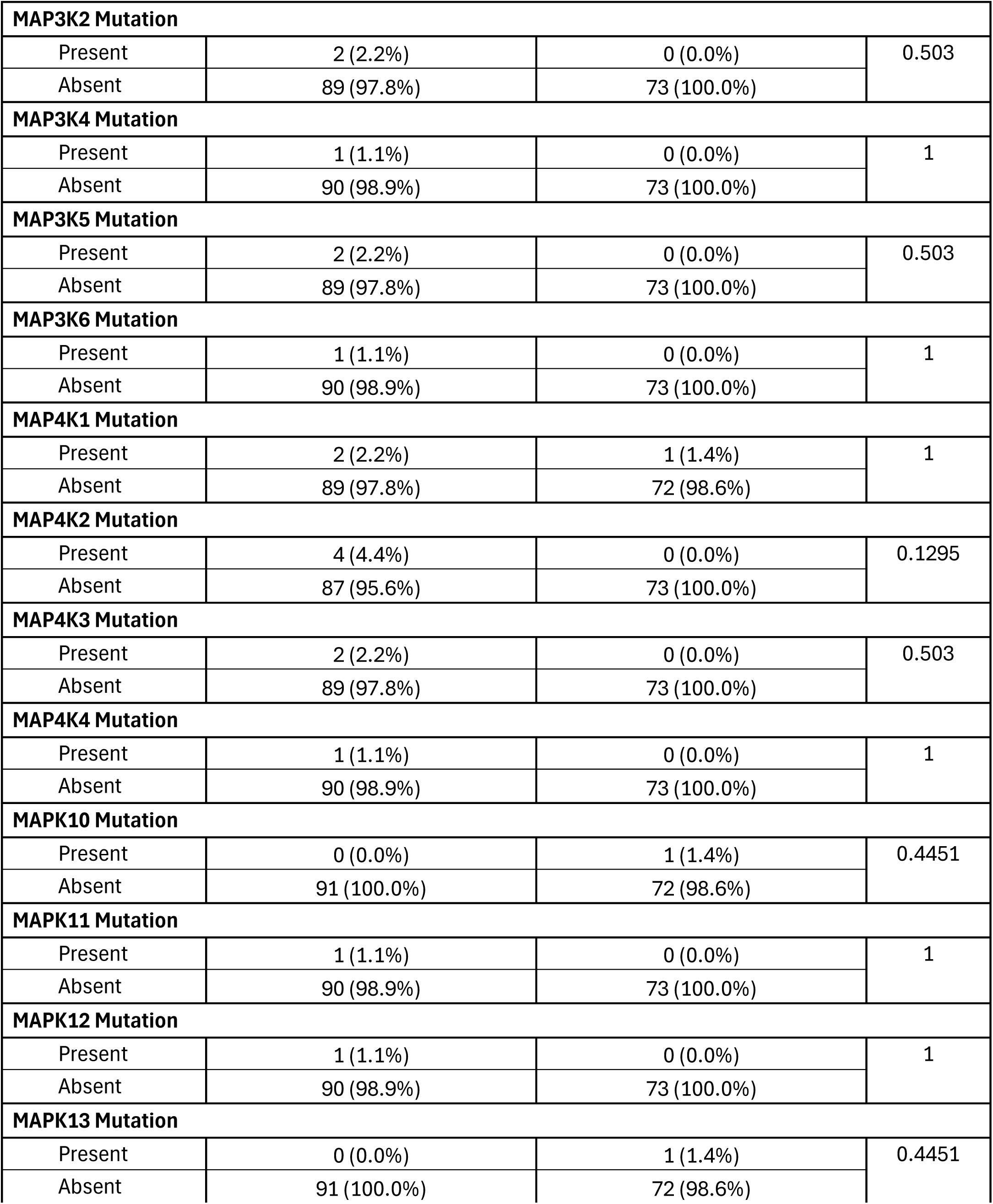
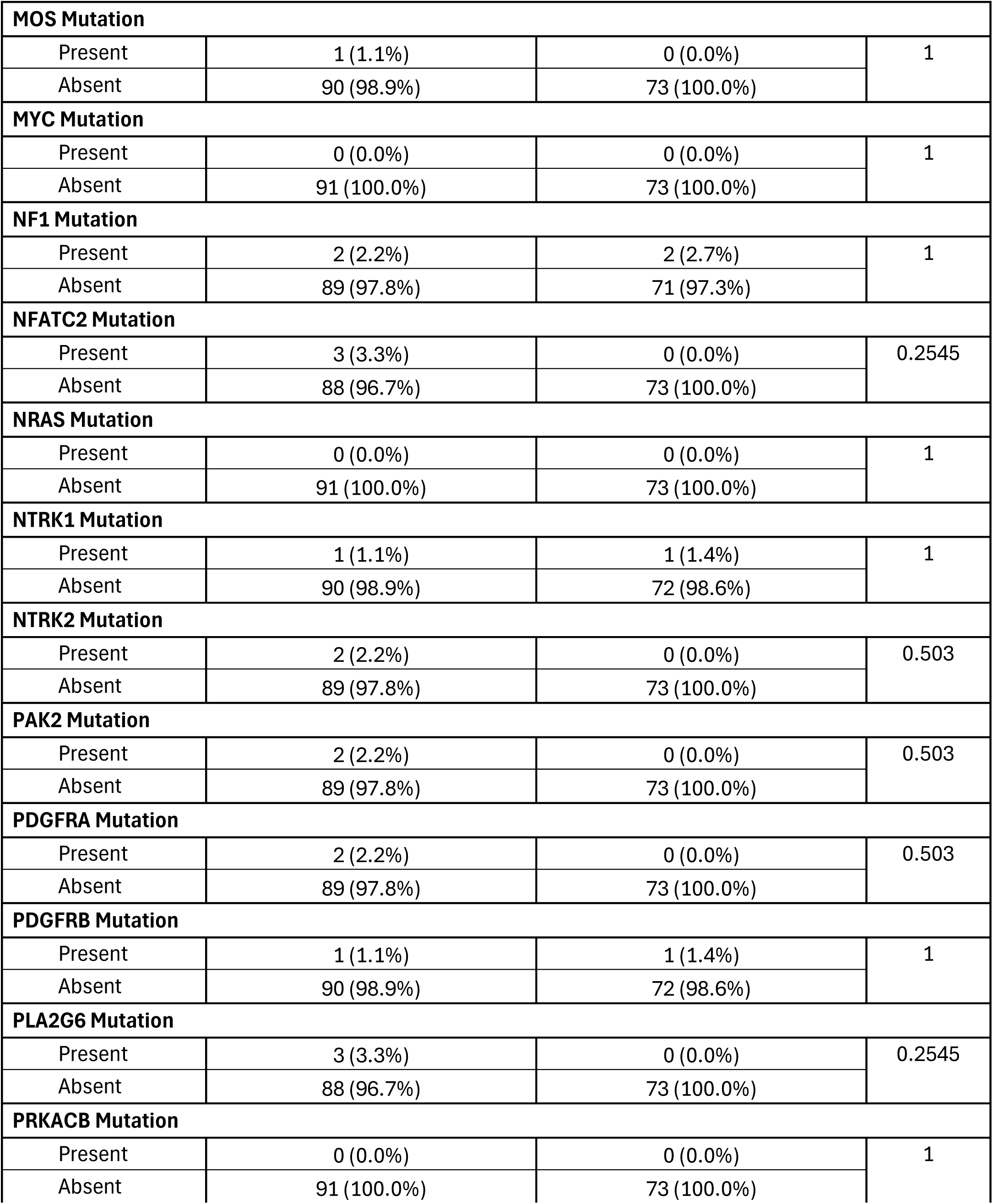
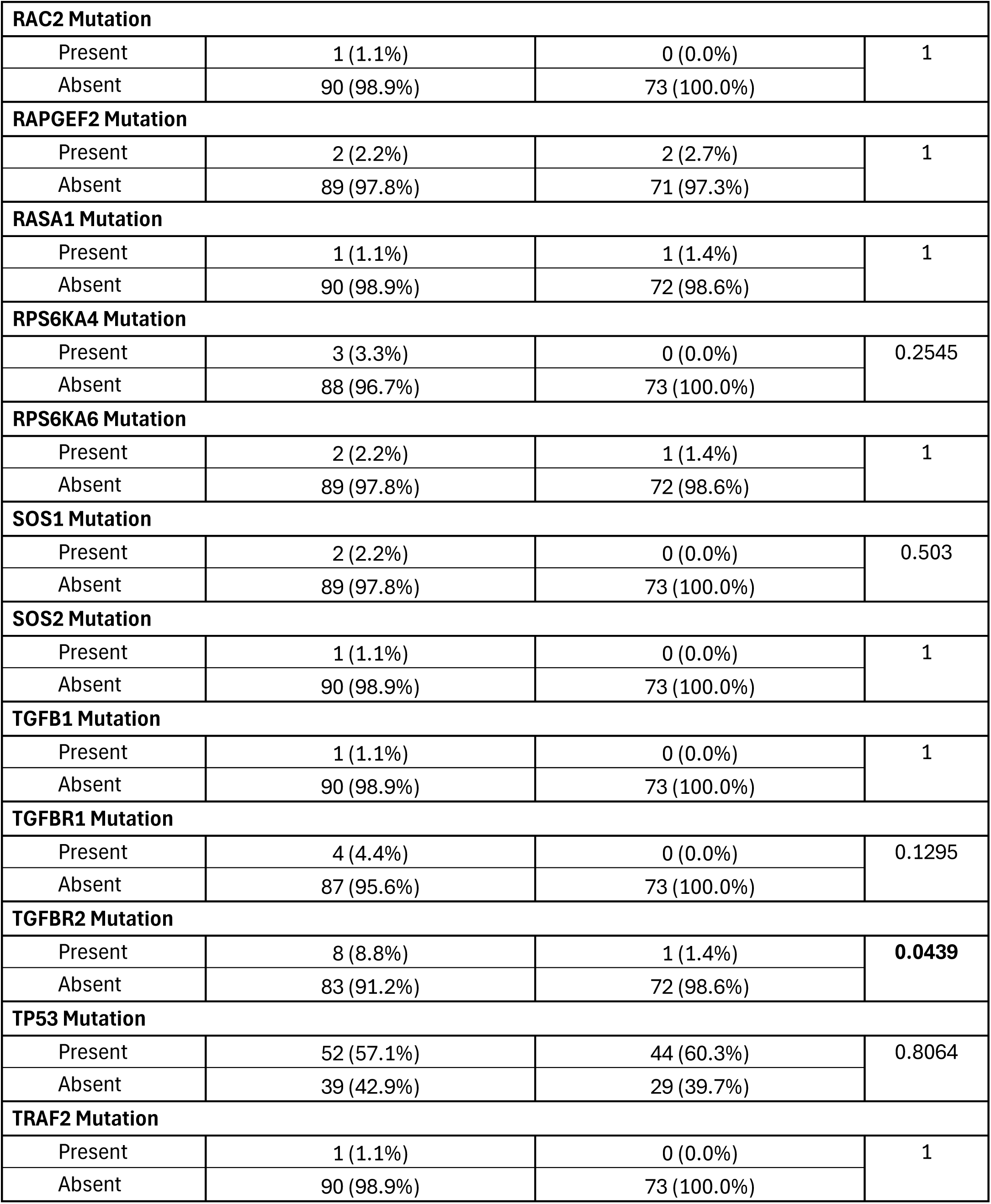
Comparison of Late-Onset PDAC Patients Treated with Gemcitabine Versus Those Not Treated with Gemcitabine.

**Table S7.**
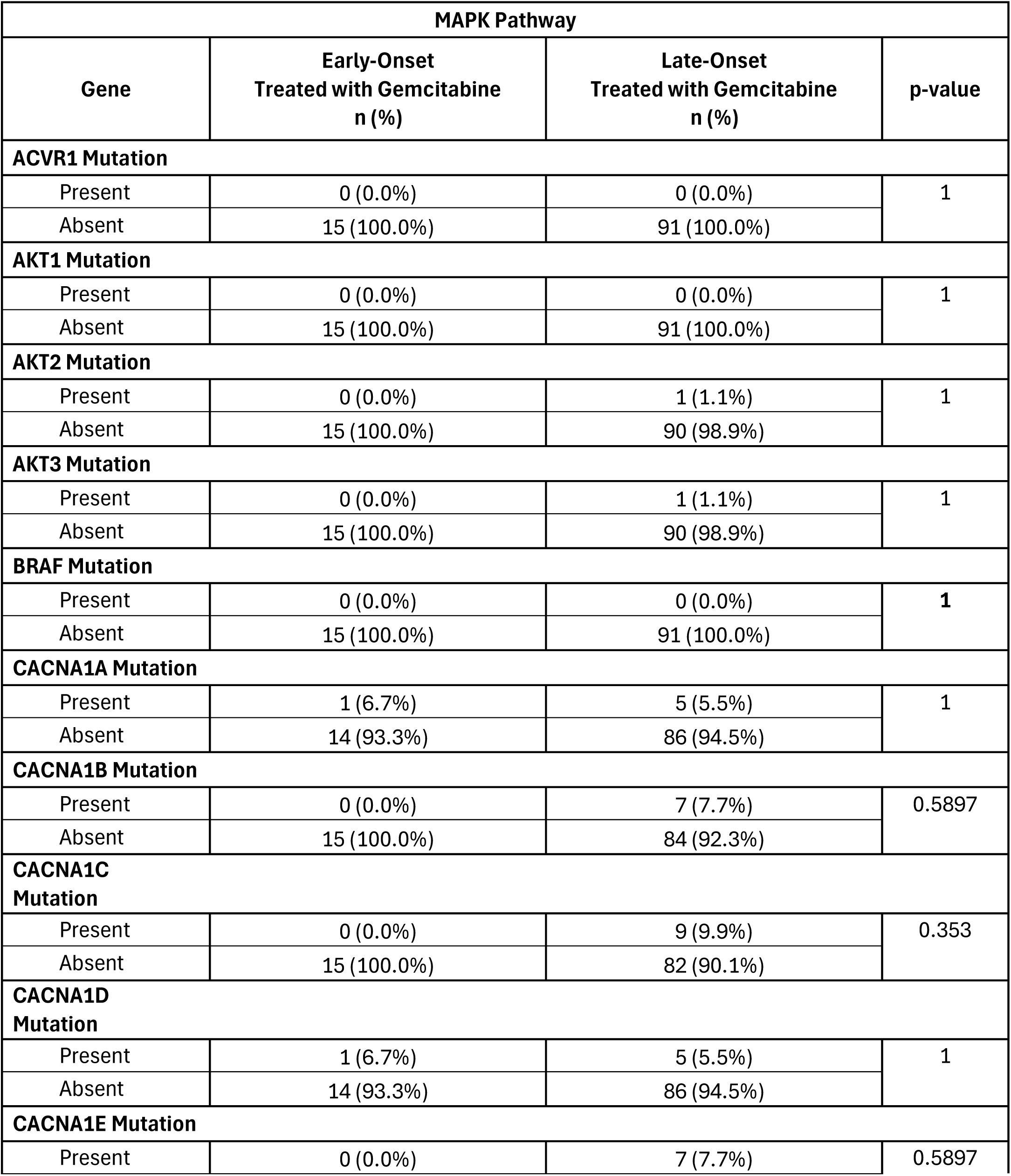
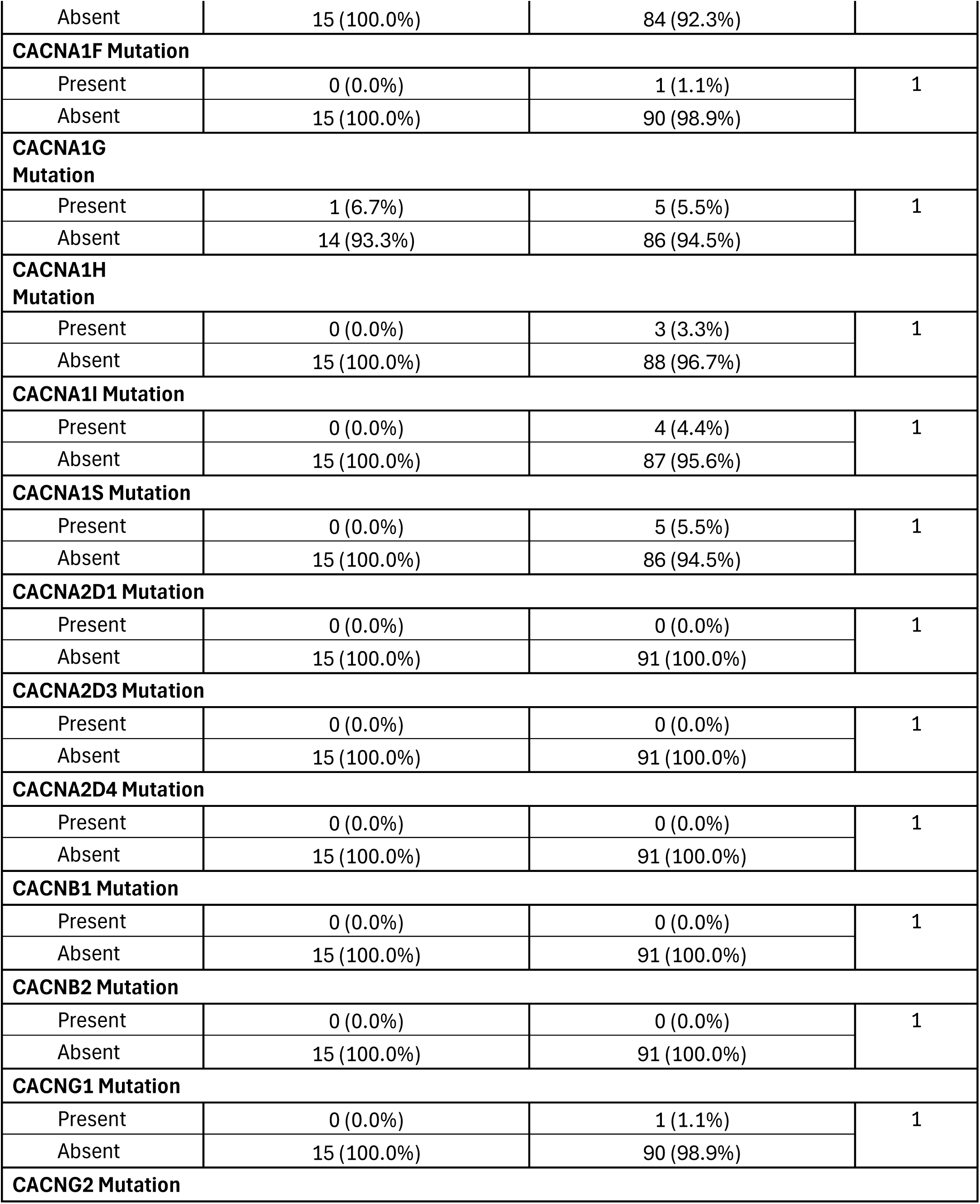
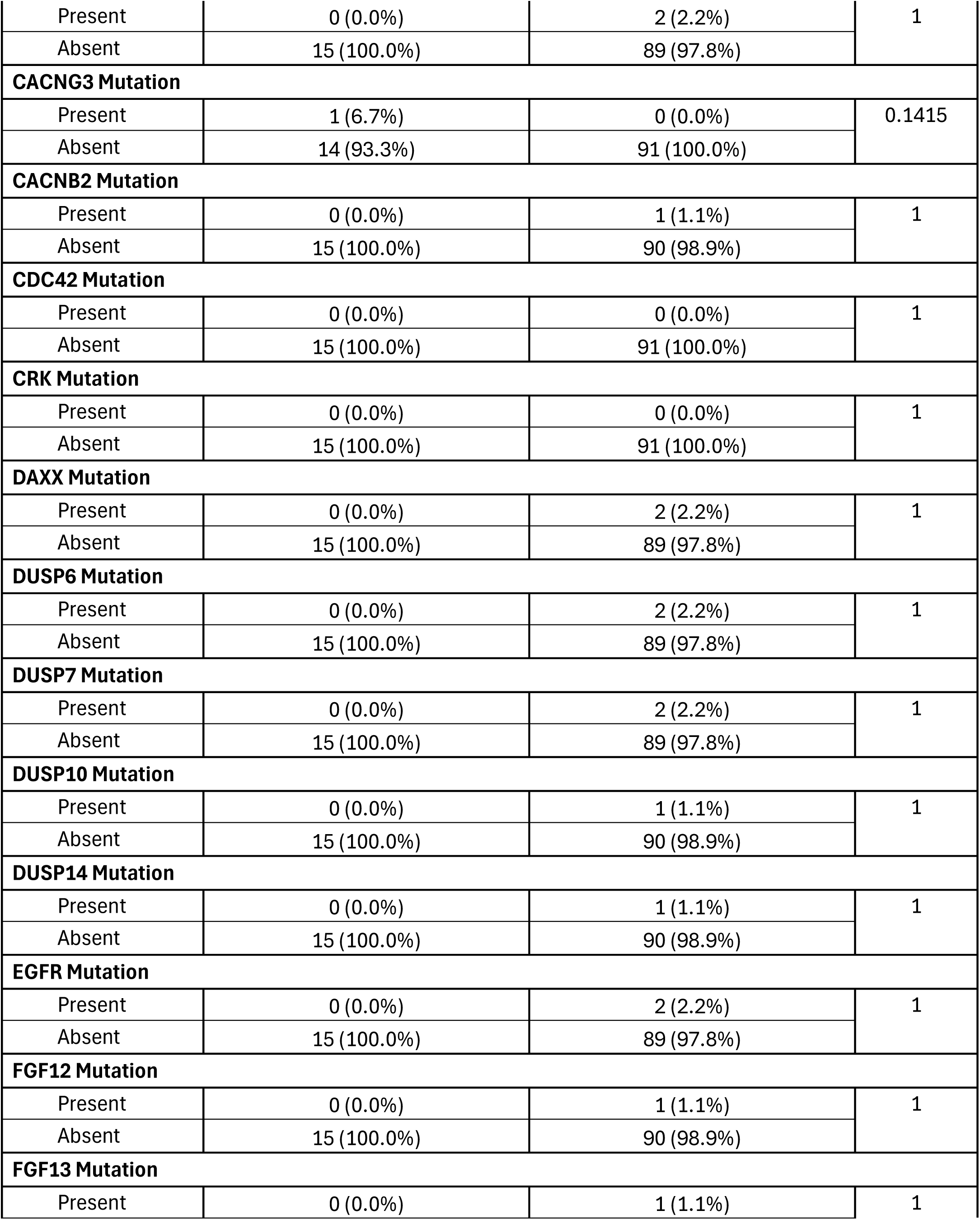
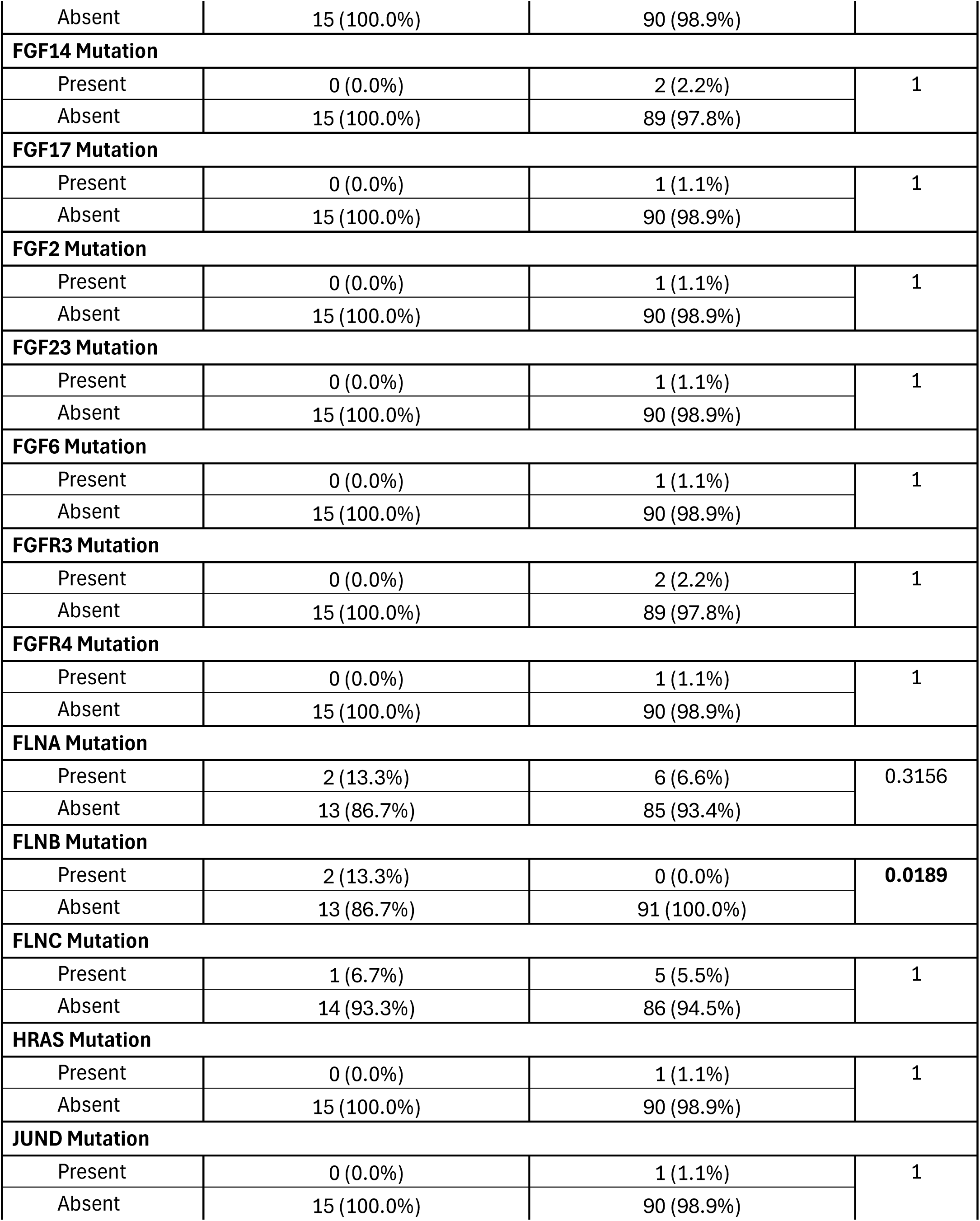
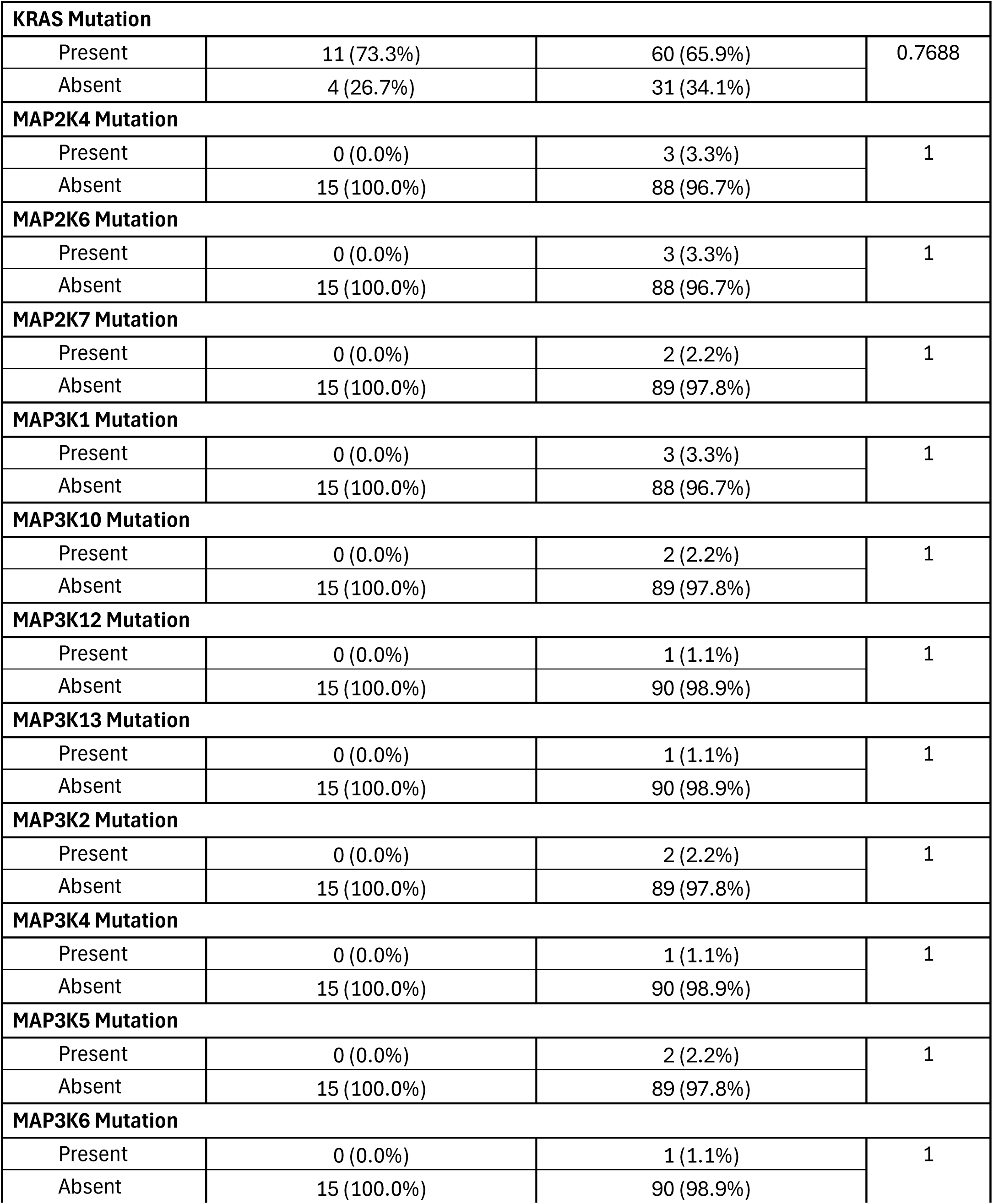
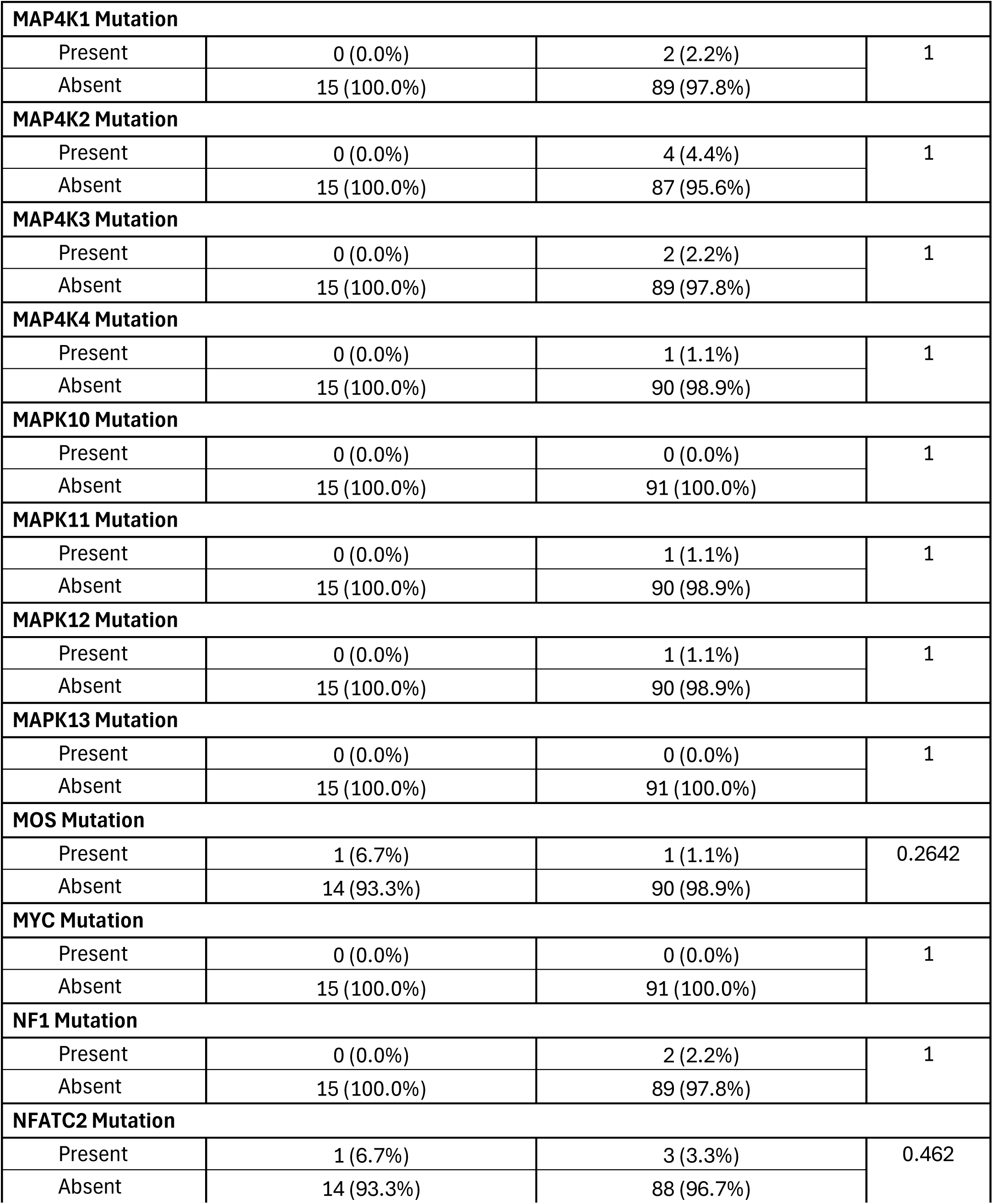
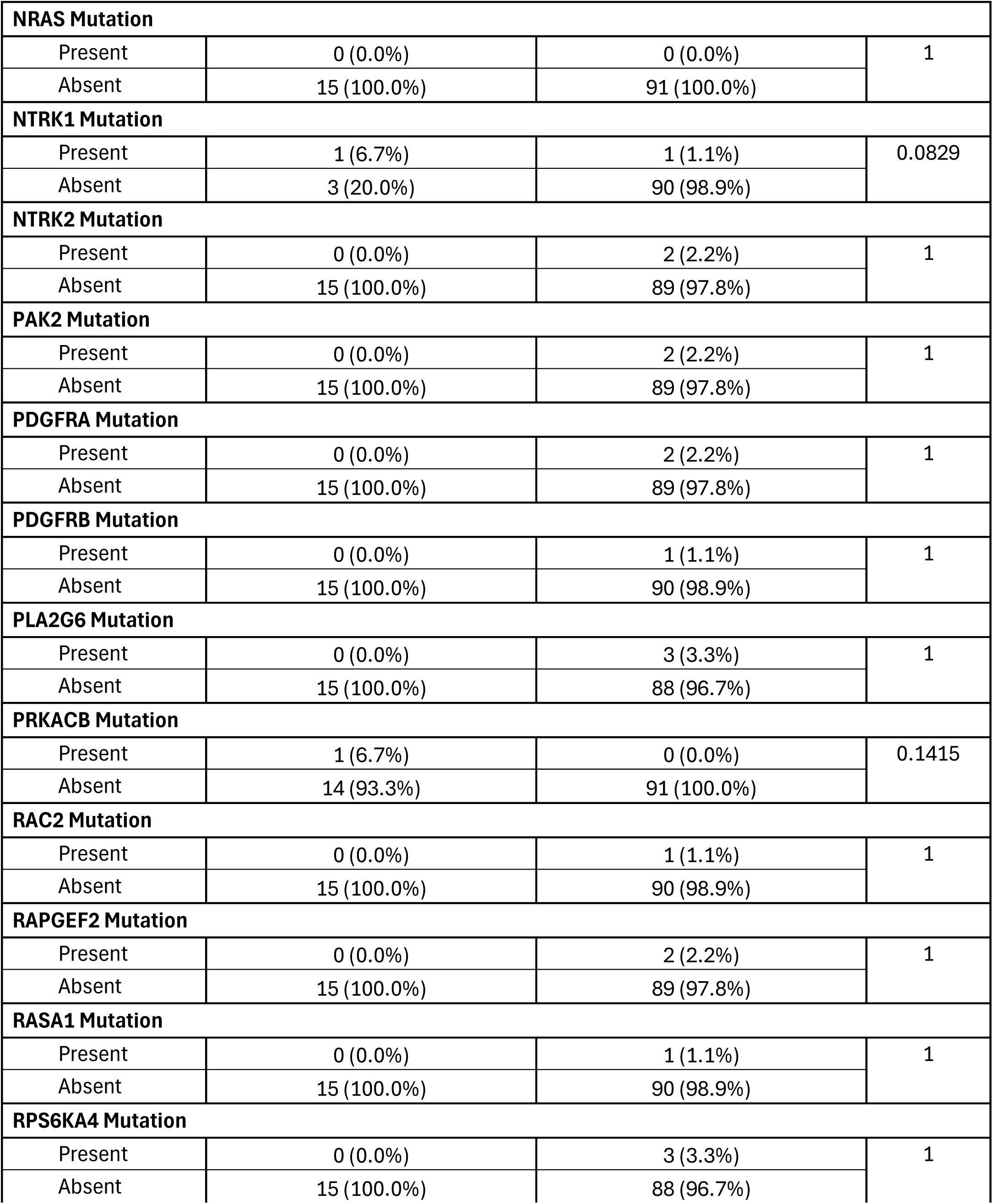
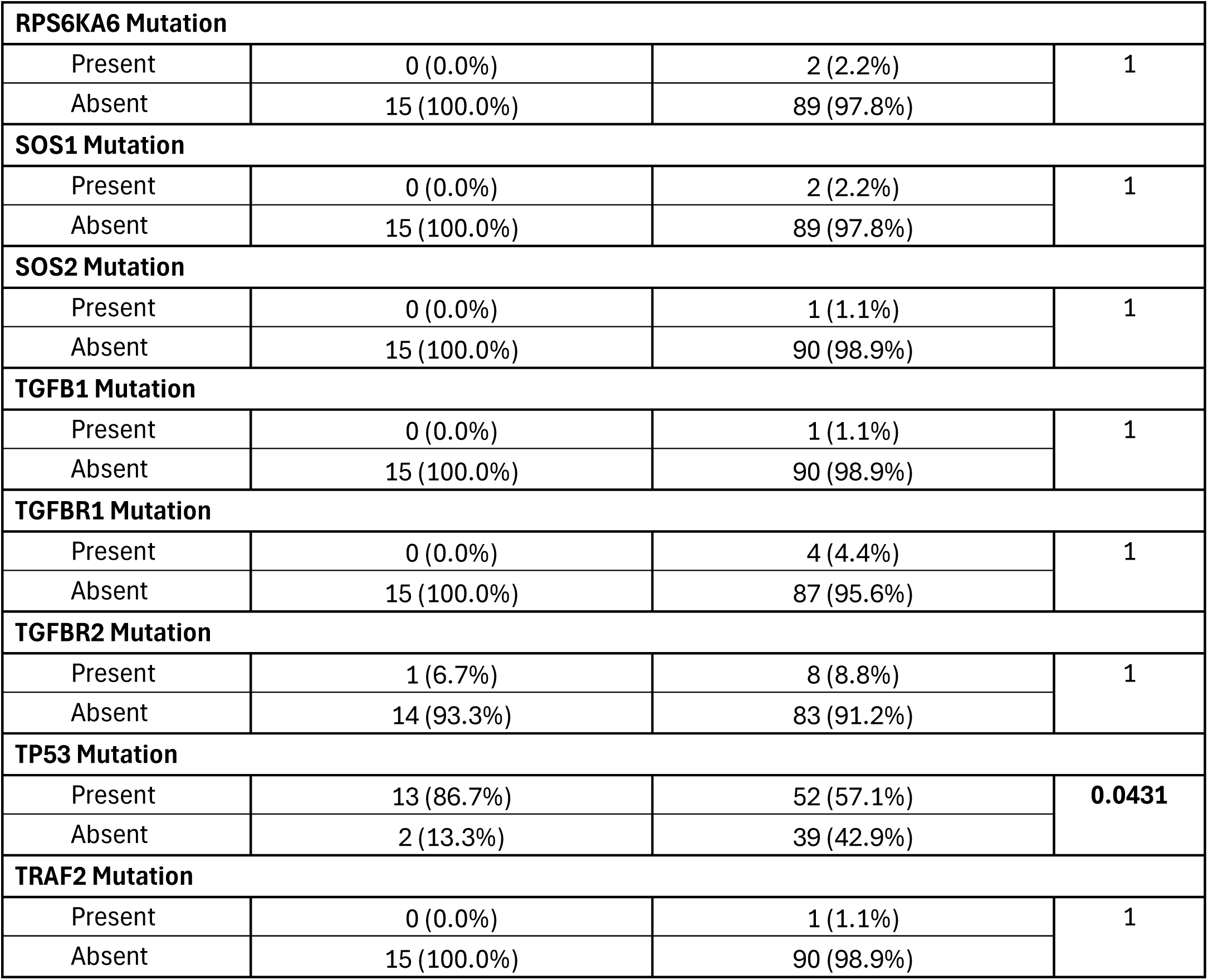
Comparison of Early-Onset PDAC Patients Versus Late-Onset PDAC Patients Treated with Gemcitabine.

**Table S8.** Comparison of Early-Onset PDAC Patients Versus Late-Onset PDAC Patients Not Treated with Gemcitabine.

| MAPK Pathway |  |  |  |
| --- | --- | --- | --- |
| Gene | Early-Onset<br>Not Treated with Gemcitabine<br>n (%) | Late-Onset<br>Not Treated with Gemcitabine<br>n (%) | p-value |
| ACVR1 Mutation |  |  |  |
| Present | 0 (0.0%) | 0 (0.0%) | 1 |
| Absent | 5 (100.0%) | 73 (100.0%) |  |
| AKT1 Mutation |  |  |  |
| Present | 0 (0.0%) | 0 (0.0%) | 1 |
| Absent | 5 (100.0%) | 73 (100.0%) |  |
| AKT2 Mutation |  |  |  |
| Present | 0 (0.0%) | 0 (0.0%) | 1 |
| Absent | 5 (100.0%) | 73 (100.0%) |  |
| AKT3 Mutation |  |  |  |
| Present | 0 (0.0%) | 0 (0.0%) | 1 |
| Absent | 5 (100.0%) | 73 (100.0%) |  |
| BRAF Mutation |  |  |  |
| Present | 0 (0.0%) | 2 (2.7%) | 1 |
| Absent | 5 (100.0%) | 71 (97.3%) |  |
| CACNA1A Mutation |  |  |  |
| Present | 0 (0.0%) | 1 (1.4%) | 1 |
| Absent | 5 (100.0%) | 72 (98.6%) |  |
| CACNA1B Mutation |  |  |  |
| Present | 0 (0.0%) | 4 (5.5%) | 1 |
| Absent | 5 (100.0%) | 69 (94.5%) |  |
| CACNA1C Mutation |  |  |  |
| Present | 0 (0.0%) | 0 (0.0%) | 1 |
| Absent | 5 (100.0%) | 73 (100.0%) |  |
| CACNA1D Mutation |  |  |  |
| Present | 0 (0.0%) | 1 (1.4%) | 1 |
| Absent | 5 (100.0%) | 72 (98.6%) |  |
| CACNA1E Mutation |  |  |  |
| Present | 0 (0.0%) | 0 (0.0%) | 1 |
| Absent | 5 (100.0%) | 73 (100.0%) |  |
| CACNA1F Mutation |  |  |  |
| Present | 0 (0.0%) | 1 (1.4%) | 1 |
| Absent | 5 (100.0%) | 72 (98.6%) |  |
| CACNA1G Mutation |  |  |  |
| Present | 0 (0.0%) | 1 (1.4%) | 1 |
| Absent | 5 (100.0%) | 72 (98.6%) |  |
| CACNA1H Mutation |  |  |  |
| Present | 0 (0.0%) | 1 (1.4%) | 1 |
| Absent | 5 (100.0%) | 72 (98.6%) |  |
| CACNA1I Mutation |  |  |  |
| Present | 0 (0.0%) | 2 (2.7%) | 1 |
| Absent | 5 (100.0%) | 71 (97.3%) |  |
| CACNA1S Mutation |  |  |  |
| Present | 0 (0.0%) | 1 (1.4%) | 1 |
| Absent | 5 (100.0%) | 72 (98.6%) |  |
| CACNA2D1 Mutation |  |  |  |
| Present | 2 (40.0%) | 1 (1.4%) | 0.0097 |
| Absent | 3 (60.0%) | 72 (98.6%) |  |
| CACNA2D3 Mutation |  |  |  |
| Present | 2 (40.0%) | 1 (1.4%) | 0.0097 |
| Absent | 3 (60.0%) | 72 (98.6%) |  |
| CACNA2D4 Mutation |  |  |  |
| Present | 2 (40.0%) | 1 (1.4%) | 0.0097 |
| Absent | 3 (60.0%) | 72 (98.6%) |  |
| CACNB1 Mutation |  |  |  |
| Present | 0 (0.0%) | 2 (2.7%) | 1 |
| Absent | 5 (100.0%) | 71 (97.3%) |  |
| CACNB2 Mutation |  |  |  |
| Present | 0 (0.0%) | 2 (2.7%) | 1 |
| Absent | 5 (100.0%) | 71 (97.3%) |  |
| CACNG1 Mutation |  |  |  |
| Present | 0 (0.0%) | 0 (0.0%) | 1 |
| Absent | 5 (100.0%) | 73 (100.0%) |  |
| CACNG2 Mutation |  |  |  |
| Present | 0 (0.0%) | 0 (0.0%) | 1 |
| Absent | 5 (100.0%) | 73 (100.0%) |  |
| CACNG3 Mutation |  |  |  |
| Present | 0 (0.0%) | 1 (1.4%) | 1 |
| Absent | 5 (100.0%) | 72 (98.6%) |  |
| CACNB2 Mutation |  |  |  |
| Present | 0 (0.0%) | 0 (0.0%) | 1 |
| Absent | 5 (100.0%) | 73 (100.0%) |  |
| CDC42 Mutation |  |  |  |
| Present | 0 (0.0%) | 1 (1.4%) | 1 |
| Absent | 5 (100.0%) | 72 (98.6%) |  |
| CRK Mutation |  |  |  |
| Present | 0 (0.0%) | 0 (0.0%) | 1 |
| Absent | 5 (100.0%) | 73 (100.0%) |  |
| DAXX Mutation |  |  |  |
| Present | 0 (0.0%) | 1 (1.4%) | 1 |
| Absent | 5 (100.0%) | 72 (98.6%) |  |
| DUSP6 Mutation |  |  |  |
| Present | 0 (0.0%) | 0 (0.0%) | 1 |
| Absent | 5 (100.0%) | 73 (100.0%) |  |
| DUSP7 Mutation |  |  |  |
| Present | 0 (0.0%) | 0 (0.0%) | 1 |
| Absent | 5 (100.0%) | 73 (100.0%) |  |
| DUSP10 Mutation |  |  |  |
| Present | 0 (0.0%) | 0 (0.0%) | 1 |
| Absent | 5 (100.0%) | 73 (100.0%) |  |
| DUSP14 Mutation |  |  |  |
| Present | 0 (0.0%) | 0 (0.0%) | 1 |
| Absent | 5 (100.0%) | 73 (100.0%) |  |
| EGFR Mutation |  |  |  |
| Present | 0 (0.0%) | 0 (0.0%) | 1 |
| Absent | 5 (100.0%) | 73 (100.0%) |  |
| FGF12 Mutation |  |  |  |
| Present | 0 (0.0%) | 0 (0.0%) | 1 |
| Absent | 5 (100.0%) | 73 (100.0%) |  |
| FGF13 Mutation |  |  |  |
| Present | 0 (0.0%) | 1 (1.4%) | 1 |
| Absent | 5 (100.0%) | 72 (98.6%) |  |
| FGF14 Mutation |  |  |  |
| Present | 0 (0.0%) | 0 (0.0%) | 1 |
| Absent | 5 (100.0%) | 73 (100.0%) |  |
| FGF17 Mutation |  |  |  |
| Present | 0 (0.0%) | 0 (0.0%) | 1 |
| Absent | 5 (100.0%) | 73 (100.0%) |  |
| FGF2 Mutation |  |  |  |
| Present | 0 (0.0%) | 0 (0.0%) | 1 |
| Absent | 5 (100.0%) | 73 (100.0%) |  |
| FGF23 Mutation |  |  |  |
| Present | 0 (0.0%) | 0 (0.0%) | 1 |
| Absent | 5 (100.0%) | 73 (100.0%) |  |
| FGF6 Mutation |  |  |  |
| Present | 1 (20.0%) | 0 (0.0%) | 0.0641 |
| Absent | 4 (80.0%) | 73 (100.0%) |  |
| FGFR3 Mutation |  |  |  |
| Present | 0 (0.0%) | 0 (0.0%) | 1 |
| Absent | 5 (100.0%) | 73 (100.0%) |  |
| FGFR4 Mutation |  |  |  |
| Present | 0 (0.0%) | 0 (0.0%) | 1 |
| Absent | 5 (100.0%) | 73 (100.0%) |  |
| FLNA Mutation |  |  |  |
| Present | 0 (0.0%) | 2 (2.7%) | 1 |
| Absent | 5 (100.0%) | 71 (97.3%) |  |
| FLNB Mutation |  |  |  |
| Present | 0 (0.0%) | 0 (0.0%) | 1 |
| Absent | 5 (100.0%) | 73 (100.0%) |  |
| FLNC Mutation |  |  |  |
| Present | 0 (0.0%) | 4 (5.5%) | 1 |
| Absent | 5 (100.0%) | 69 (94.5%) |  |
| HRAS Mutation |  |  |  |
| Present | 0 (0.0%) | 0 (0.0%) | 1 |
| Absent | 5 (100.0%) | 73 (100.0%) |  |
| JUND Mutation |  |  |  |
| Present | 0 (0.0%) | 0 (0.0%) | 1 |
| Absent | 5 (100.0%) | 73 (100.0%) |  |
| KRAS Mutation |  |  |  |
| Present | 4 (80.0%) | 46 (63.0%) | 0.6491 |
| Absent | 1 (20.0%) | 27 (37.0%) |  |
| MAP2K4 Mutation |  |  |  |
| Present | 0 (0.0%) | 1 (1.4%) | 1 |
| Absent | 5 (100.0%) | 72 (98.6%) |  |
| MAP2K6 Mutation |  |  |  |
| Present | 0 (0.0%) | 0 (0.0%) | 1 |
| Absent | 5 (100.0%) | 73 (100.0%) |  |
| MAP2K7 Mutation |  |  |  |
| Present | 0 (0.0%) | 1 (1.4%) | 1 |
| Absent | 5 (100.0%) | 72 (98.6%) |  |
| MAP3K1 Mutation |  |  |  |
| Present | 0 (0.0%) | 0 (0.0%) | 1 |
| Absent | 5 (100.0%) | 73 (100.0%) |  |
| MAP3K10 Mutation |  |  |  |
| Present | 0 (0.0%) | 0 (0.0%) | 1 |
| Absent | 5 (100.0%) | 73 (100.0%) |  |
| MAP3K12 Mutation |  |  |  |
| Present | 0 (0.0%) | 0 (0.0%) | 1 |
| Absent | 5 (100.0%) | 73 (100.0%) |  |
| MAP3K13 Mutation |  |  |  |
| Present | 0 (0.0%) | 0 (0.0%) | 1 |
| Absent | 5 (100.0%) | 73 (100.0%) |  |
| MAP3K2 Mutation |  |  |  |
| Present | 0 (0.0%) | 0 (0.0%) | 1 |
| Absent | 5 (100.0%) | 73 (100.0%) |  |
| MAP3K4 Mutation |  |  |  |
| Present | 0 (0.0%) | 0 (0.0%) | 1 |
| Absent | 5 (100.0%) | 73 (100.0%) |  |
| MAP3K5 Mutation |  |  |  |
| Present | 0 (0.0%) | 0 (0.0%) | 1 |
| Absent | 5 (100.0%) | 73 (100.0%) |  |
| MAP3K6 Mutation |  |  |  |
| Present | 0 (0.0%) | 0 (0.0%) | 1 |
| Absent | 5 (100.0%) | 73 (100.0%) |  |
| MAP4K1 Mutation |  |  |  |
| Present | 0 (0.0%) | 1 (1.4%) | 1 |
| Absent | 5 (100.0%) | 72 (98.6%) |  |
| MAP4K2 Mutation |  |  |  |
| Present | 0 (0.0%) | 0 (0.0%) | 1 |
| Absent | 5 (100.0%) | 73 (100.0%) |  |
| MAP4K3 Mutation |  |  |  |
| Present | 0 (0.0%) | 0 (0.0%) | 1 |
| Absent | 5 (100.0%) | 73 (100.0%) |  |
| MAP4K4 Mutation |  |  |  |
| Present | 0 (0.0%) | 0 (0.0%) | 1 |
| Absent | 5 (100.0%) | 73 (100.0%) |  |
| MAPK10 Mutation |  |  |  |
| Present | 0 (0.0%) | 1 (1.4%) | 1 |
| Absent | 5 (100.0%) | 72 (98.6%) |  |
| MAPK11 Mutation |  |  |  |
| Present | 0 (0.0%) | 0 (0.0%) | 1 |
| Absent | 5 (100.0%) | 73 (100.0%) |  |
| MAPK12 Mutation |  |  |  |
| Present | 0 (0.0%) | 0 (0.0%) | 1 |
| Absent | 5 (100.0%) | 73 (100.0%) |  |
| MAPK13 Mutation |  |  |  |
| Present | 0 (0.0%) | 1 (1.4%) | 1 |
| Absent | 5 (100.0%) | 72 (98.6%) |  |
| MOS Mutation |  |  |  |
| Present | 0 (0.0%) | 0 (0.0%) | 1 |
| Absent | 5 (100.0%) | 73 (100.0%) |  |
| MYC Mutation |  |  |  |
| Present | 0 (0.0%) | 0 (0.0%) | 1 |
| Absent | 5 (100.0%) | 73 (100.0%) |  |
| NF1 Mutation |  |  |  |
| Present | 0 (0.0%) | 2 (2.7%) | 1 |
| Absent | 5 (100.0%) | 71 (97.3%) |  |
| NFATC2 Mutation |  |  |  |
| Present | 0 (0.0%) | 0 (0.0%) | 1 |
| Absent | 5 (100.0%) | 73 (100.0%) |  |
| NRAS Mutation |  |  |  |
| Present | 0 (0.0%) | 0 (0.0%) | 1 |
| Absent | 5 (100.0%) | 73 (100.0%) |  |
| NTRK1 Mutation |  |  |  |
| Present | 0 (0.0%) | 1 (1.4%) | 1 |
| Absent | 5 (100.0%) | 72 (98.6%) |  |
| NTRK2 Mutation |  |  |  |
| Present | 0 (0.0%) | 0 (0.0%) | 1 |
| Absent | 5 (100.0%) | 73 (100.0%) |  |
| PAK2 Mutation |  |  |  |
| Present | 0 (0.0%) | 0 (0.0%) | 1 |
| Absent | 5 (100.0%) | 73 (100.0%) |  |
| PDGFRA Mutation |  |  |  |
| Present | 0 (0.0%) | 0 (0.0%) | 1 |
| Absent | 5 (100.0%) | 73 (100.0%) |  |
| PDGFRB Mutation |  |  |  |
| Present | 0 (0.0%) | 1 (1.4%) | 1 |
| Absent | 5 (100.0%) | 72 (98.6%) |  |
| PLA2G6 Mutation |  |  |  |
| Present | 0 (0.0%) | 0 (0.0%) | 1 |
| Absent | 5 (100.0%) | 73 (100.0%) |  |
| PRKACB Mutation |  |  |  |
| Present | 0 (0.0%) | 0 (0.0%) | 1 |
| Absent | 5 (100.0%) | 73 (100.0%) |  |
| RAC2 Mutation |  |  |  |
| Present | 0 (0.0%) | 0 (0.0%) | 1 |
| Absent | 5 (100.0%) | 73 (100.0%) |  |
| RAPGEF2 Mutation |  |  |  |
| Present | 0 (0.0%) | 2 (2.7%) | 1 |
| Absent | 5 (100.0%) | 71 (97.3%) |  |
| RASA1 Mutation |  |  |  |
| Present | 0 (0.0%) | 1 (1.4%) | 1 |
| Absent | 5 (100.0%) | 72 (98.6%) |  |
| RPS6KA4 Mutation |  |  |  |
| Present | 0 (0.0%) | 0 (0.0%) | 1 |
| Absent | 5 (100.0%) | 73 (100.0%) |  |
| RPS6KA6 Mutation |  |  |  |
| Present | 0 (0.0%) | 1 (1.4%) | 1 |
| Absent | 5 (100.0%) | 72 (98.6%) |  |
| SOS1 Mutation |  |  |  |
| Present | 0 (0.0%) | 0 (0.0%) | 1 |
| Absent | 5 (100.0%) | 73 (100.0%) |  |
| SOS2 Mutation |  |  |  |
| Present | 0 (0.0%) | 0 (0.0%) | 1 |
| Absent | 5 (100.0%) | 73 (100.0%) |  |
| TGFB1 Mutation |  |  |  |
| Present | 0 (0.0%) | 0 (0.0%) | 1 |
| Absent | 5 (100.0%) | 73 (100.0%) |  |
| TGFBR1 Mutation |  |  |  |
| Present | 0 (0.0%) | 0 (0.0%) | 1 |
| Absent | 5 (100.0%) | 73 (100.0%) |  |
| TGFBR2 Mutation |  |  |  |
| Present | 0 (0.0%) | 1 (1.4%) | 1 |
| Absent | 5 (100.0%) | 72 (98.6%) |  |
| TP53 Mutation |  |  |  |
| Present | 2 (40.0%) | 44 (60.3%) | 0.396 |
| Absent | 3 (60.0%) | 29 (39.7%) |  |
| TRAF2 Mutation |  |  |  |
| Present | 0 (0.0%) | 0 (0.0%) | 1 |
| Absent | 5 (100.0%) | 73 (100.0%) |  |

**Figure S1.**
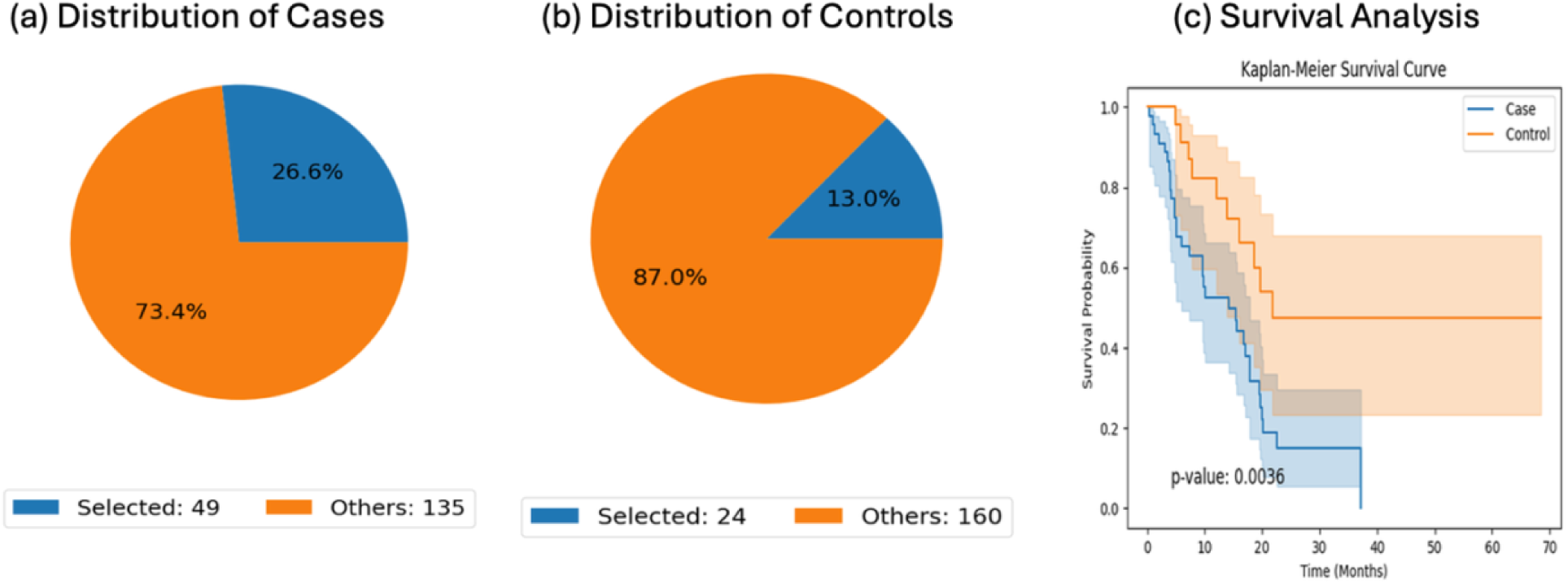
Conversational AI–enabled cohort definition and survival analysis in late-onset PDAC patients not treated with gemcitabine, stratified by RTK–RAS pathway status. This figure illustrates AI-guided cohort selection and outcome comparison within late-onset PDAC patients who did not receive gemcitabine. Using natural language criteria, the AI-HOPE-RTK-RAS agent identified (a) a case cohort comprising late-onset, non–gemcitabine-treated patients harboring RTK–RAS pathway alterations (n = 49; 26.6% of the dataset), and (b) a control cohort of similarly defined patients lacking RTK–RAS alterations (n = 24; 13.0%). Pie charts display the proportional representation of selected versus non-selected samples within the full cohort. (c) Kaplan–Meier overall survival analysis demonstrated a statistically significant difference between groups (log-rank p = 0.0036), with RTK–RAS–altered tumors associated with reduced survival compared with pathway–wild-type tumors. Shaded areas denote 95% confidence intervals. These results underscore the prognostic relevance of RTK–RAS pathway status in late-onset PDAC outside the context of gemcitabine exposure and highlight the capacity of conversational AI to reproducibly construct clinically meaningful cohorts.

**Figure S2.**
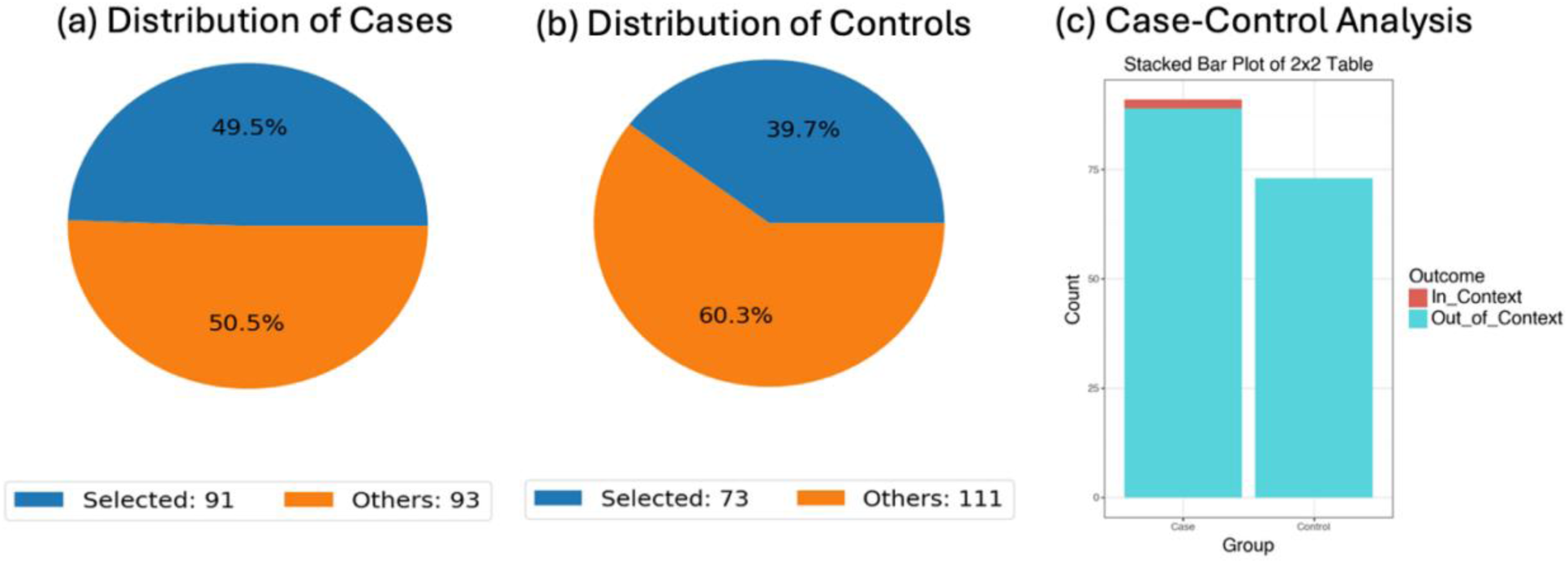
Conversational AI–guided comparison of ERBB2 mutation frequency in late-onset PDAC patients stratified by gemcitabine exposure. This figure illustrates an AI-enabled odds ratio analysis evaluating whether ERBB2 mutation prevalence differs between late-onset PDAC patients treated with gemcitabine (case cohort; n = 91) and those not treated with gemcitabine (control cohort; n = 73). Cohorts were constructed using structured natural language criteria within the AI-HOPE-RTK-RAS framework. Pie charts display the proportion of selected samples within each group relative to the total dataset. The stacked bar plot summarizes ERBB2-mutated (“In-Context”) versus non-mutated (“Out-of-Context”) samples across case and control cohorts. ERBB2 mutations were observed in 2.2% of gemcitabine-treated cases and 0.68% of non-treated controls. Fisher’s exact test demonstrated no statistically significant difference between groups (p = 0.576), and the estimated odds ratio indicated no meaningful enrichment of ERBB2 mutations associated with gemcitabine exposure in late-onset PDAC. These findings suggest that ERBB2 mutation frequency does not substantially differ by gemcitabine treatment status within this age-defined subgroup and demonstrate the utility of conversational AI for rapid genomic frequency comparisons.

**Figure S3.**
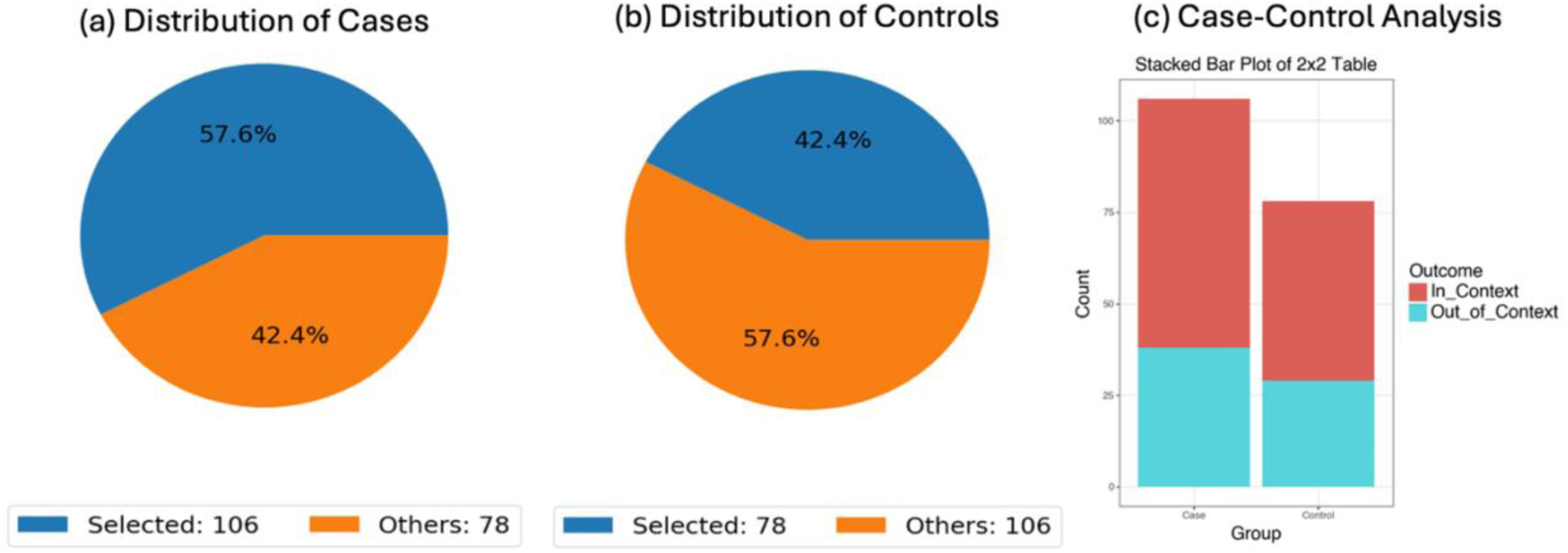
Conversational AI–enabled comparison of KRAS mutation prevalence by gemcitabine treatment status in PDAC. This figure depicts an AI-assisted odds ratio analysis evaluating whether KRAS mutation frequency differs between PDAC patients treated with gemcitabine (case cohort; n = 106) and those not treated with gemcitabine (control cohort; n = 78). Cohorts were defined using structured clinical criteria within the AI-HOPE-RTK-RAS framework. Pie charts illustrate the proportion of selected samples within each treatment group relative to the full dataset. The stacked bar plot summarizes KRAS-mutated (“In-Context”) and KRAS–wild-type (“Out-of-Context”) samples across treated and non-treated cohorts. KRAS mutations were highly prevalent in both groups (64.1% in treated vs. 62.8% in non-treated patients). Statistical testing demonstrated no significant difference in mutation frequency by gemcitabine exposure (Fisher’s exact p = 0.976; odds ratio 1.059, 95% CI 0.577–1.943). These findings indicate that KRAS mutation prevalence is comparable regardless of gemcitabine treatment status, reinforcing its near-ubiquitous role in PDAC biology and highlighting the utility of conversational AI for rapid treatment-stratified genomic comparisons.

**Figure S4.**
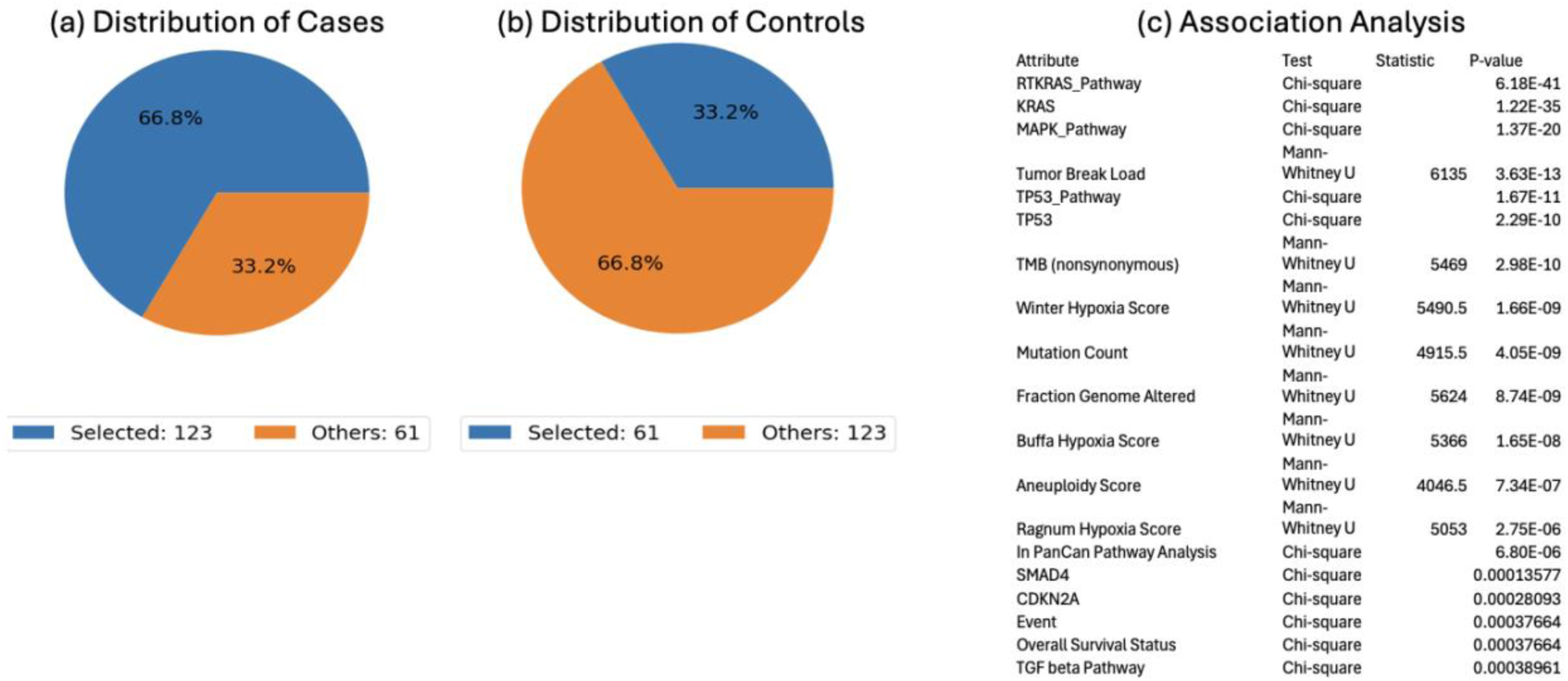
Conversational AI-driven association analysis of clinical and molecular attributes linked to RTK-RAS pathway status in PDAC. This figure summarizes an AI-guided exploratory analysis comparing PDAC samples harboring RTK-RAS pathway alterations (case cohort; n = 123) with pathway-non-altered tumors (control cohort; n = 61). Panels (a) and (b) illustrate the proportional distribution of selected (in-context) and unselected samples within each cohort, demonstrating that RTK-RAS-altered tumors comprised 66.8% of the dataset, whereas 33.2% were pathway-non-altered. Panel (c) presents the results of a comprehensive association analysis integrating categorical (Chi-square) and continuous (Mann-Whitney U) tests to identify attributes significantly linked to RTK-RAS pathway status. As expected, strong associations were observed with KRAS mutation status and MAPK pathway alterations. Additional significant associations included TP53 mutation status, TP53 pathway involvement, SMAD4 and CDKN2A alterations, and TGFβ pathway status. Quantitative genomic features such as tumor mutation burden (nonsynonymous), total mutation count, fraction of genome altered, aneuploidy score, and multiple hypoxia signatures (Winter, Buffa, Ragnum) were also significantly enriched in RTK-RAS-altered tumors. Clinical outcome variables, including overall survival status and event occurrence, were likewise associated with pathway status. These AI-derived associations highlight a coordinated genomic, pathway-level, and microenvironmental signature linked to RTK-RAS alteration status in PDAC, demonstrating the capacity of conversational AI to rapidly uncover multidimensional clinical-molecular relationships.

**Figure S5.**
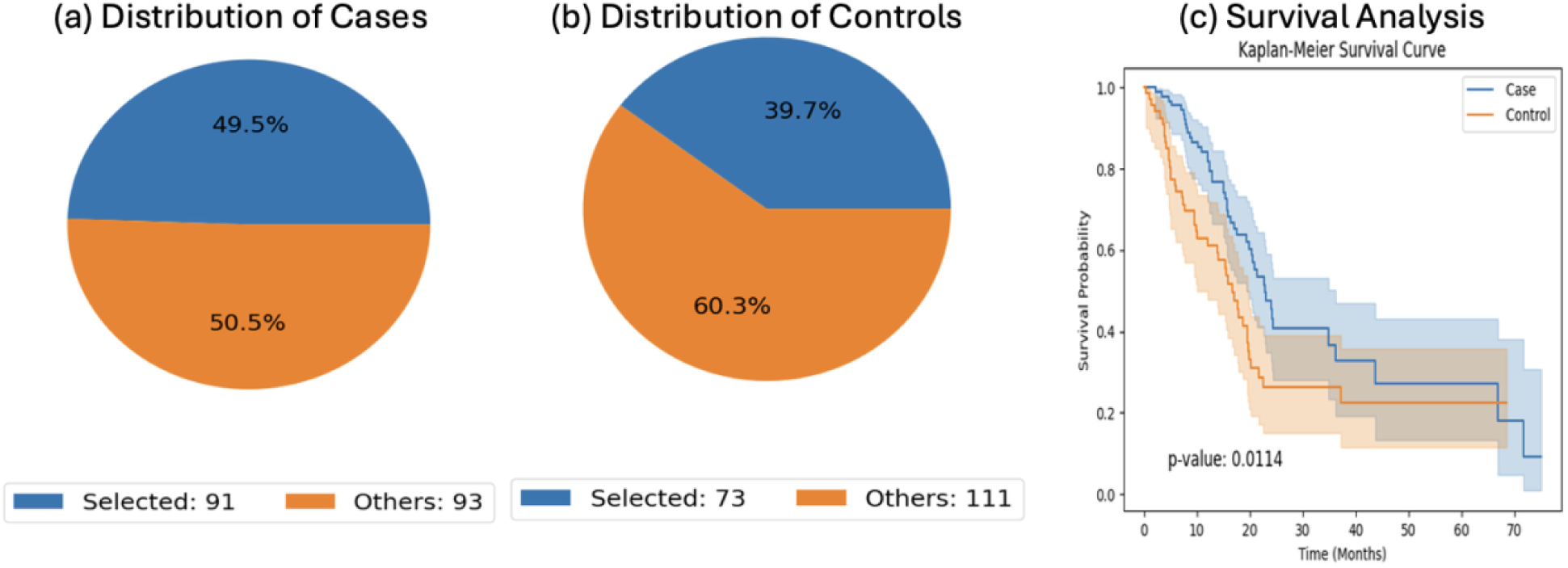
Conversational AI-driven cohort selection and survival analysis for late-onset PDAC stratified by MAPK pathway status and gemcitabine exposure. This figure demonstrates AI-enabled cohort construction and outcome comparison in late-onset PDAC. Using structured natural language queries, the AI-HOPE-MAPK module identified (a) a case cohort of gemcitabine-treated late-onset patients (n = 91; 49.5% of the dataset) and (b) a control cohort of non-gemcitabine-treated late-onset patients (n = 73; 39.7%). Pie charts depict the proportional distribution of selected versus non-selected samples within the full cohort. Panel (c) presents Kaplan-Meier overall survival curves comparing the two groups. A statistically significant difference in survival was observed (log-rank p = 0.0114), with gemcitabine-treated patients demonstrating distinct survival dynamics relative to untreated controls over time. Shaded regions indicate 95% confidence intervals. This figure highlights the capacity of conversational AI to rapidly define treatment-contextual cohorts and to generate reproducible survival analyses aligned with pathway-focused research questions in PDAC.

**Figure S6.**
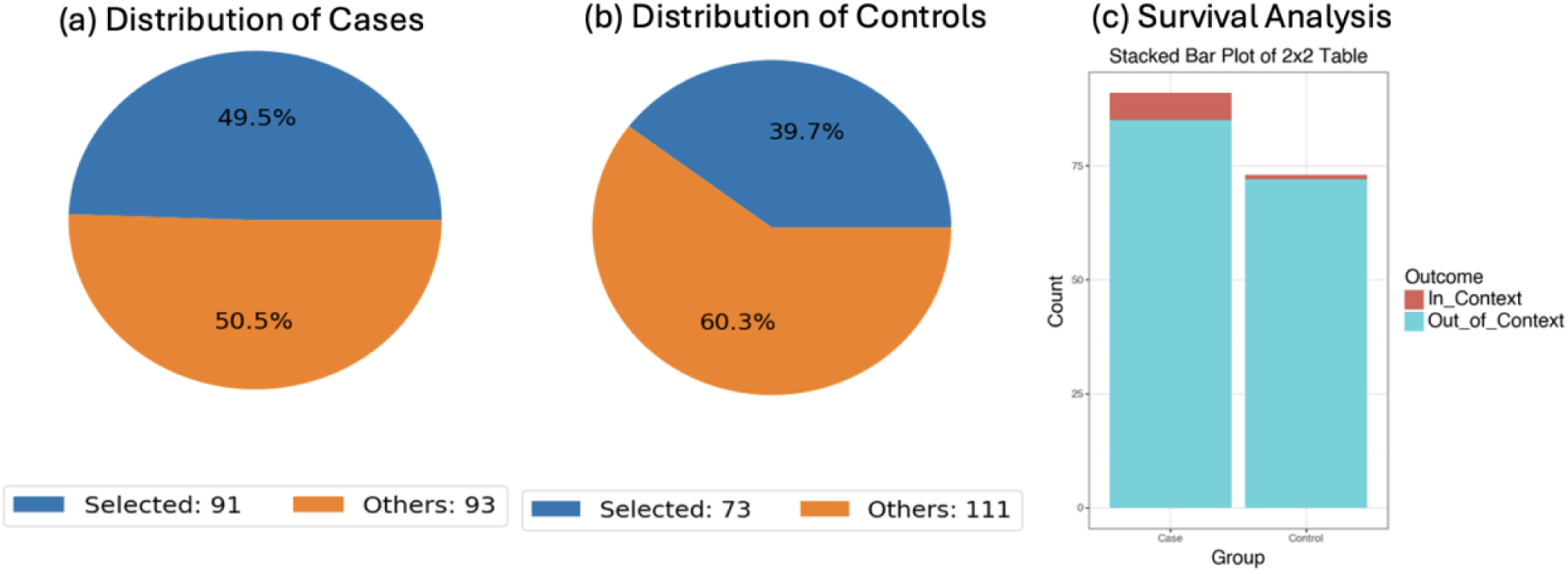
Conversational AI-based evaluation of TGFBR2 mutation frequency in late-onset PDAC stratified by gemcitabine exposure. This figure presents an AI-enabled odds ratio analysis assessing whether TGFBR2 mutation prevalence differs between late-onset PDAC patients treated with gemcitabine (case cohort; n = 91) and those not treated with gemcitabine (control cohort; n = 73). Cohorts were defined using structured clinical filters within the AI-HOPE-MAPK framework. Panels (a) and (b) display the distribution of selected (“in-context,” TGFBR2-mutated) and unselected (“out-of-context,” non-mutated) samples within each treatment group. The stacked bar plot summarizes mutation frequencies across case and control cohorts. TGFBR2 mutations were observed in 6.59% of gemcitabine-treated patients compared with 1.37% of non-treated patients. Statistical comparison using Fisher’s exact test did not demonstrate a significant association between gemcitabine exposure and TGFBR2 mutation status (p = 0.209; odds ratio 5.082, 95% CI 0.598-43.204). These findings indicate that, although numerically higher in the treated cohort, TGFBR2 mutation frequency does not differ significantly by gemcitabine exposure in late-onset PDAC. The analysis highlights the ability of conversational AI to rapidly generate treatment-stratified genomic comparisons within pathway-focused investigations.

**Figure S7.**
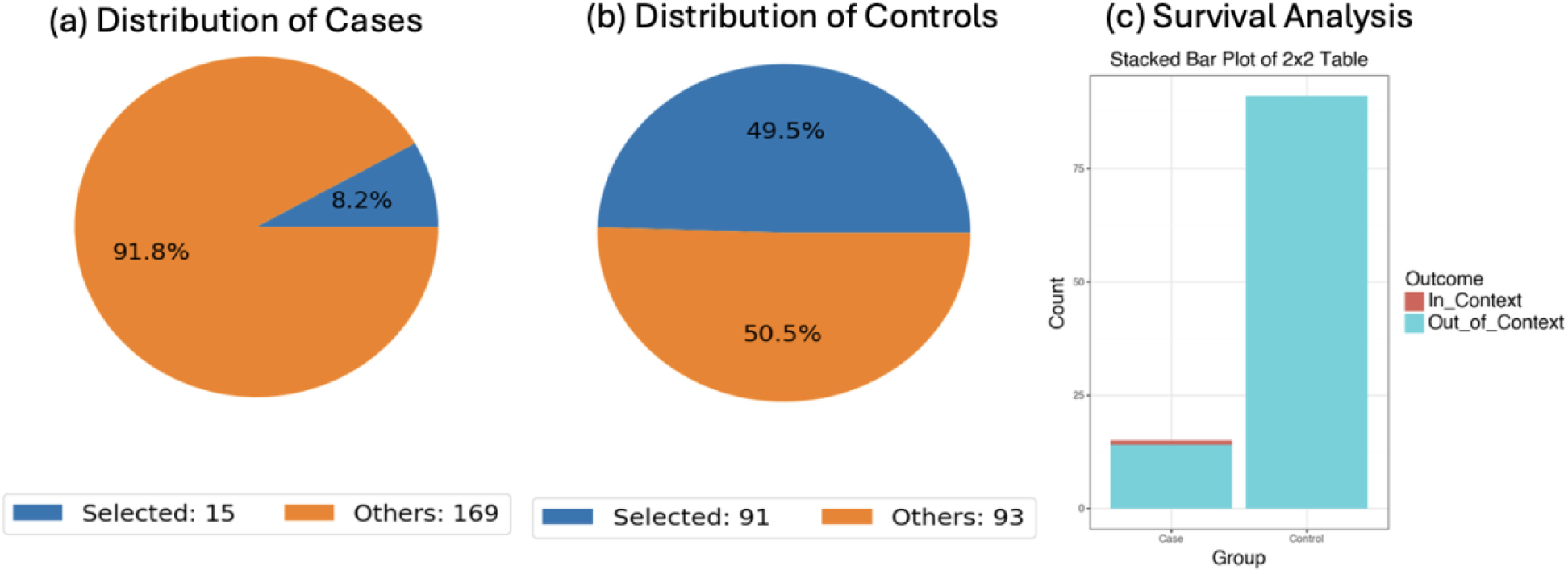
Conversational AI-enabled comparison of FLNB mutation frequency in gemcitabine-treated PDAC stratified by age at onset. This figure presents an AI-guided odds ratio analysis assessing whether FLNB mutation prevalence differs between early-onset (EO) and late-onset (LO) PDAC patients who received gemcitabine. The case cohort included EO gemcitabine-treated patients (n = 15), and the control cohort comprised LO gemcitabine-treated patients (n = 91), defined using structured clinical filters within the AI-HOPE-MAPK framework. Panels (a) and (b) illustrate the proportion of FLNB-mutated (“in-context”) and non-mutated (“out-of-context”) samples within each subgroup. FLNB mutations were observed in 6.67% of EO treated patients compared with 0.55% of LO treated patients. Panel (c) displays a stacked bar plot summarizing the 2×2 comparison. Statistical testing did not demonstrate a significant association between age group and FLNB mutation frequency in the gemcitabine-treated setting (Fisher’s exact p = 0.301; odds ratio 13.0, 95% CI 0.416-405.866). Although numerically enriched in early-onset treated tumors, FLNB mutation prevalence did not reach statistical significance, likely reflecting small sample size. This analysis highlights the utility of conversational AI for rapidly performing age-stratified, treatment-specific genomic comparisons within MAPK pathway investigations.

**Figure S8.**
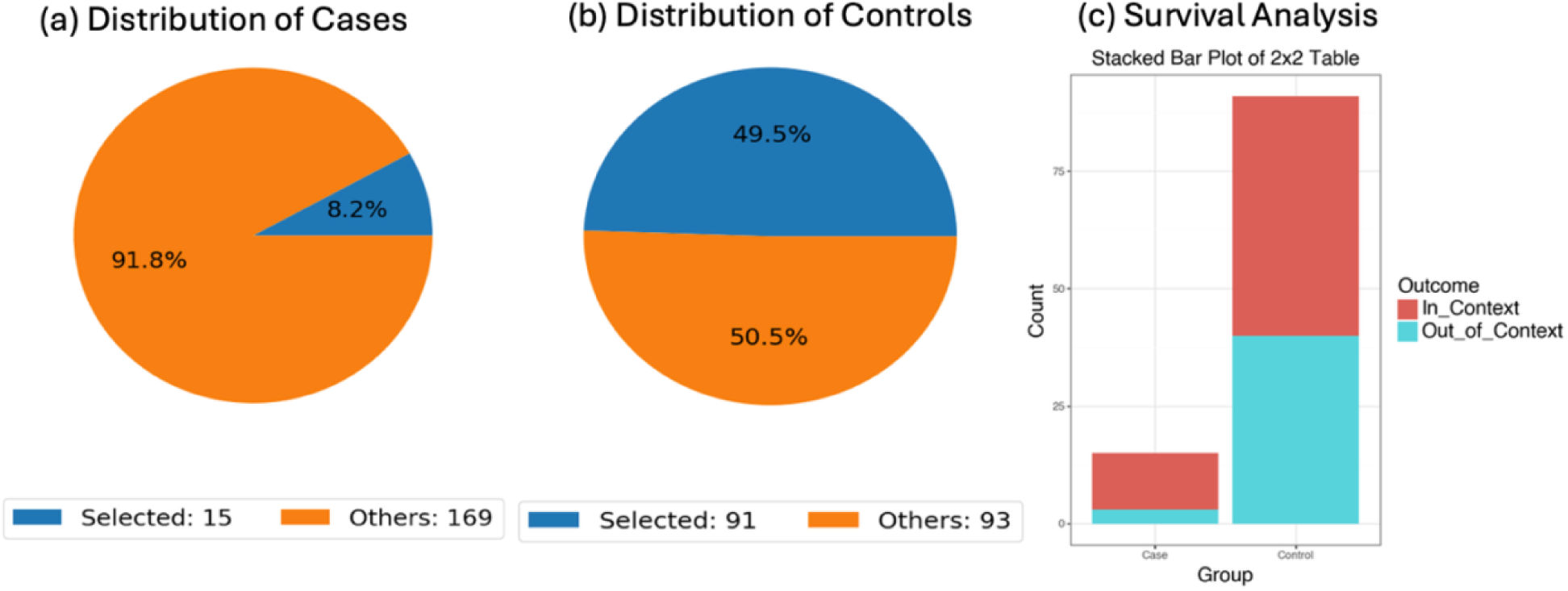
Conversational AI-driven evaluation of TP53 mutation frequency in gemcitabine-treated PDAC stratified by age at onset. This figure depicts an AI-enabled comparison of TP53 mutation prevalence between early-onset (EO) and late-onset (LO) PDAC patients who received gemcitabine. The case cohort included EO, gemcitabine-treated patients (n = 15), while the control cohort comprised LO, gemcitabine-treated patients (n = 91), identified through structured natural language queries within the AI-HOPE framework. Panels (a) and (b) present pie charts summarizing the proportion of TP53-mutated (“in-context”) and non-mutated (“out-of-context”) samples in each subgroup. TP53 alterations were observed in 80.0% of EO treated tumors compared with 56.0% of LO treated tumors. Panel (c) illustrates the corresponding 2×2 comparison using a stacked bar plot. Although TP53 mutations were numerically more frequent in early-onset treated patients, statistical testing did not demonstrate a significant association between age category and TP53 mutation status in the gemcitabine-treated setting (Chi-square p = 0.142; odds ratio 3.137, 95% CI 0.829-11.876). These findings suggest a potential enrichment of TP53 alterations in early-onset PDAC receiving gemcitabine; however, the lack of statistical significance and limited sample size warrant cautious interpretation. This example further highlights the capacity of conversational AI to rapidly perform age- and treatment-specific genomic comparisons within MAPK pathway-focused analyses.

**Figure S9.**
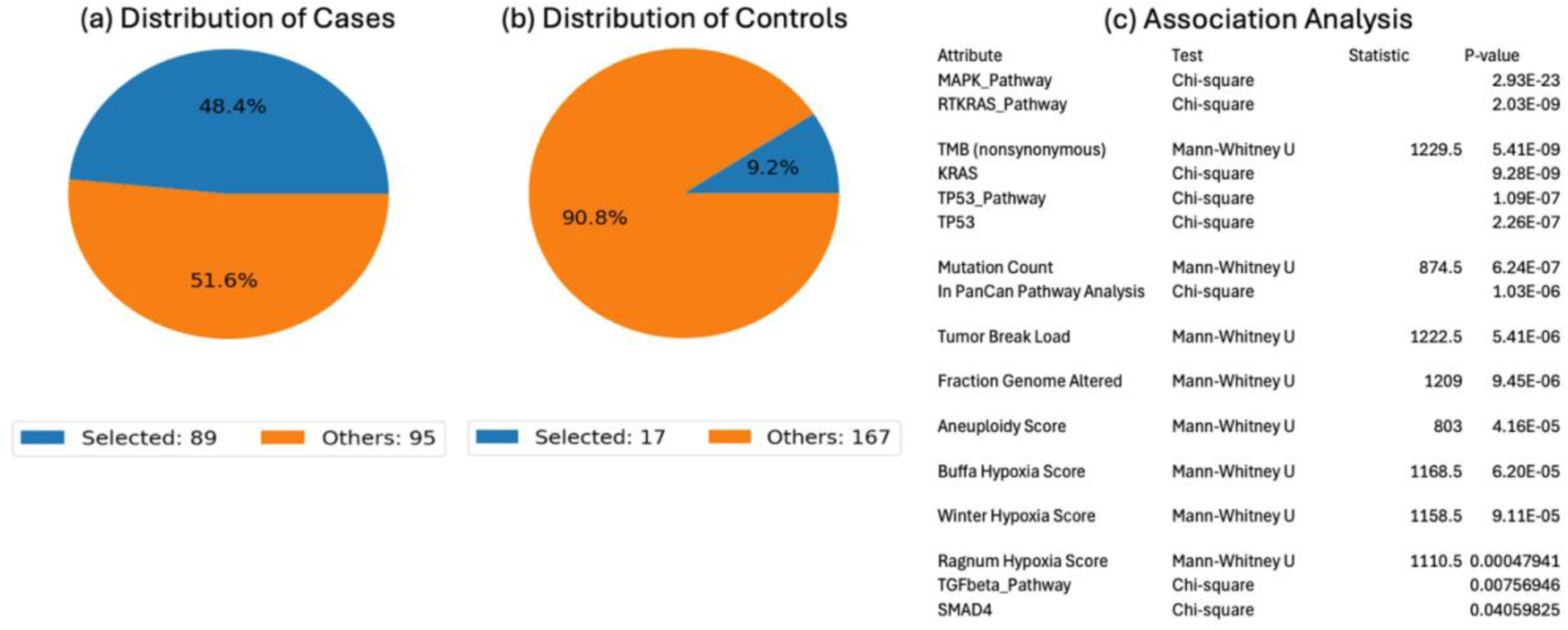
AI-enabled identification of clinical and genomic features associated with MAPK pathway alterations in gemcitabine-treated PDAC. This figure summarizes a conversational AI-guided comparative analysis of PDAC tumors treated with gemcitabine, stratified by MAPK pathway status. The case cohort consisted of gemcitabine-treated tumors harboring MAPK pathway alterations (n = 89), while the control cohort included gemcitabine-treated, MAPK-non-altered tumors (n = 17). Panels (a) and (b) display the distribution of selected (MAPK-altered) and unselected samples within each subgroup, illustrating the predominance of pathway-altered tumors among gemcitabine-treated cases. Panel (c) presents the results of a comprehensive association analysis integrating Chi-square tests for categorical variables and Mann-Whitney U tests for continuous measures. Strong associations were observed between MAPK pathway alteration status and RTK-RAS pathway involvement, KRAS mutation status, and TP53 pathway alterations. Quantitative genomic metrics, including nonsynonymous tumor mutation burden, total mutation count, tumor break load, fraction of genome altered, and aneuploidy score, were significantly enriched in MAPK-altered tumors. Hypoxia-related signatures (Winter, Buffa, and Ragnum scores) also demonstrated significant differences between groups. Additionally, TGFβ pathway and SMAD4 alterations were associated with MAPK pathway status in the gemcitabine-treated context. Collectively, these findings reveal that MAPK-altered PDAC treated with gemcitabine exhibits a distinct molecular and genomic instability profile, underscoring the value of conversational AI for rapidly uncovering multidimensional pathway-dependent associations in treatment-specific settings.

